# Physical interventions to interrupt or reduce the spread of respiratory viruses. Part 2 - Hand hygiene and other hygiene measures: systematic review and meta-analysis

**DOI:** 10.1101/2020.04.14.20065250

**Authors:** Lubna Al-Ansary, Ghada Bawazeer, Elaine Beller, Justin Clark, John Conly, Chris Del Mar, Elizabeth Dooley, Eliana Ferroni, Paul Glasziou, Tammy Hoffmann, Tom Jefferson, Sarah Thorning, Mieke van Driel, Mark Jones

**Affiliations:** Dept of Family and Community, College of Medicine, King Saud University; College of Pharmacy, King Saud University; Centre for Research in Evidence Based Practice (CREBP), Faculty of Health Sciences and Medicine, Bond University; Centre for Research in Evidence-Based Practice (CREBP), Faculty of Health Sciences and Medicine, Bond University; Department of Medicine, Microbiology, Immunology & Infectious Diseases, University of Calgary and Alberta Health Services; Faculty of Health Sciences and Medicine, Bond University; Institute of Evidence-Based Healthcare, Bond University; Regione Veneto, Azienda Zero; Cochrane Vaccines Field; Gold Coast Hospital and Health Service; Primary Care Clinical Unit, Faculty of Medicine, The University of Queensland

## Abstract

**OBJECTIVE:** To assess the effectiveness of hand hygiene, surface disinfecting, and other hygiene interventions in preventing or reducing the spread of illnesses from respiratory viruses.

**DESIGN:** Update of a systematic review and meta-analysis focussing on randomised controlled trials (RCTs) and cluster-RCTs (c-RCTs) evidence only.

**DATA SOURCES:** Eligible trials from the previous Cochrane review, search of the Cochrane Central Register of Controlled Trials, PubMed, Embase and CINAHL from 01 October 2010 to 01 April 2020, and forward and backward citation analysis of included studies.

**DATA SELECTION:** RCTs and c-RCTs involving people of any age, testing the use of hand hygiene methods, surface disinfection or cleaning, and other miscellaneous barrier interventions. Face masks, eye protection, and person distancing are covered in Part 1 of our systematic review. Outcomes included acute respiratory illness (ARI), influenza-like illness (ILI) or laboratory-confirmed influenza (influenza) and/or related consequences (e.g. death, absenteeism from school or work).

**DATA EXTRACTION AND ANALYSIS:** Six authors working in pairs independently assessed risk of bias using the Cochrane tool and extracted data. The generalised inverse variance method was used for pooling by using the random-effects model, and results reported with risk ratios (RR) and 95% confidence intervals (CIs).

**RESULTS:** We identified 51 eligible trials. We included 25 randomised trials comparing hand hygiene interventions with a control; 15 of these could be included in meta-analyses. We pooled 8 trials for the outcome of ARI. Hand hygiene showed a 16% relative reduction in the number of participants with ARI (RR 0.84, 95% CI 0.82 to 0.86) in the intervention group. When we considered the more strictly defined outcomes of ILI and influenza, the RR for ILI was 0.98 (95% CI 0.85 to 1.14), and for influenza the RR was 0.91 (95% CI 0.61 to 1.34). Three trials measured absenteeism. We found a 36% relative reduction in absentee numbers in the hand hygiene group (RR 0.64, 95% CI 0.58 to 0.71). Comparison of different hand hygiene interventions did not favour one intervention type over another. We found no incremental effects of combining hand hygiene with using face masks or disinfecting surfaces or objects.

**CONCLUSIONS:** Despite the lack of evidence for the impact of hand hygiene in reducing ILI and influenza, the modest evidence for reducing the burden of ARIs, and related absenteeism, justifies reinforcing the standard recommendation for hand hygiene measures to reduce the spread of respiratory viruses. Funding for relevant trials with an emphasis on adherence and compliance with such a measure is crucial to inform policy and global pandemic preparedness with confidence and precision.

## INTRODUCTION

Viral acute respiratory infections (ARIs) represent a huge burden on global health, whether during an epidemic, pandemic, or in non-epidemic situations [1]. Preventing the virus from spreading amongst people via a combination of social and physical interventions may be the only option to reduce the spread of outbreaks.

This systematic review is the second part of a review of physical interventions to interrupt or reduce the spread of respiratory viruses. Part 1 of the review examined the effectiveness of masks, eye protection, with or without person distancing [2]. This part examines the effectiveness of other physical interventions (such as hand hygiene, surface disinfecting, and other multi-component hygiene interventions) that may reduce spread by limiting the transfer of viral particles on to and from surfaces.

The last update of the Cochrane review in 2011 [3] included 23 randomised trials on hand hygiene and other hygiene measures. It was not possible to perform trial meta-analysis due to poor reporting and heterogeneity. Case-control trials were sufficiently homogenous to enable meta-analysis which provided evidence that handwashing for a minimum of 11 times a day prevented cases of SARS during the 2003 epidemic (odds ratio (OR) 0.54, 95% CI 0.44 to 0.67). Many randomised trials have been published in the past decade, and this has prompted us to include this higher-level evidence in this review.

## METHODS

### Inclusion criteria

We included randomised controlled trials (RCTs) and cluster-RCTs (C-RCTs) involving participants of all ages that tested interventions including hand hygiene (alone or with other physical interventions), surface or object disinfection, and any other physical barrier interventions with no language restriction. Face masks, eye protection, and person distancing were excluded as these were covered in Part 1 of our systematic review. We included only trials that reported an outcome measure of acute respiratory illness (ARI). Measures including influenza-like illness (ILI), influenza, or respiratory infections – with or without related consequences (e.g. days off work, complications, hospitalisation and death, if clearly reported as consequences of the respiratory illness) were eligible. Relevant RCTs from the previous versions of this Cochrane review were also included [3-5]. We excluded observational trials because of the number of available randomized datasets which we hoped would provide stronger evidence. We plan a follow up of the Cochrane review with all studies included.

### Search strategy

We identified relevant RCTs and C-RCTs from our 2011 Cochrane Review [3]. These earlier trials were analysed using word frequency to create a new search string that was used to search PubMed [6]. This search string was converted using the Polyglot Search Translator [7] and was also used to search the Cochrane Central Register of Controlled Trials, Embase and CINAHL. The searches were conducted from 01 October 2010 to 01 April 2020. Search strings for all databases are presented Appendix 1. We used Scopus to perform backwards and forward citation analysis for all new studies retrieved. We screened search and citation analysis results using the RobotSearch tool to remove all obvious non-RCTs [8]. Three authors (JC, MJ, ST) independently reviewed the titles and abstracts of the identified studies to assess eligibility for inclusion. We resolved discrepancies by consensus.

### Risk of bias assessment

Three pairs of authors worked independently (TJ/EB, LA/GB, MJ/EF) to assess risk of bias using the Cochrane risk of bias tool for randomised trials (RoB 1.0). We resolved disagreements by discussion. See Part 1 of our systematic review for further details on the risk of bias methodology [2].

### Data extraction and analysis

Three pairs of authors independently (TJ/EB, LA/GB, MJ/EF) to study data using a standard template that was developed and applied for previous versions of this Cochrane review, but revised to reflect our focus on RCTs and cRCTs only for this update. We resolved any discrepancies in the data extractions by discussion. We extracted and reported descriptions of interventions using the Template for Intervention Description and Replication (TIDieR) template [9]. We entered data on outcomes into RevMan software [10] and meta-analysed using the generalised inverse variance random-effects model. The random-effects model was chosen because we expected clinical heterogeneity due to differences in pooled interventions and outcome definitions, and methodological heterogeneity due to pooling of RCTs and C-RCTs. Where possible, we pooled estimates from C-RCTs accounting for clustering. Treatment effects were reported as risk ratios (RR) with 95% confidence interval (CI) and P values. We used the I2 statistic and chi-square test to assess statistical heterogeneity [11]. Relevant results from study types that could not be pooled were reported descriptively. Because all the authors consider themselves as past patients or potential future patients, formal opinion of patient or public representation was not sought in the design, or conduct, or reporting, or dissemination plans of our research.

## RESULTS

### Results of the search

We searched four databases (see appendix 1) and retrieved 1486 records. Backwards (screening of the reference lists) and forwards citation analysis, undertaken in Scopus, on our initial list of included trials, retrieved 1694 records (n = 3180 records). We removed 706 duplicate records for a total of 2474 records that were screened by title and abstract. We excluded 2351 records following title and abstract screening. We obtained full text publications for 123 records. During full text screening and data extraction, we excluded 92 studies as not meeting inclusion criteria. We included 31 trials reported in 31 references, 20 trials from the previous review were also added, for a total of 51 trials. For a detailed description of our screening process, see the PRISMA flow diagram in Figure 1. We also searched two trials registers [12, 13] and identified 42 additional trials, of these, we identified three ongoing trials.

**Figure 1:**
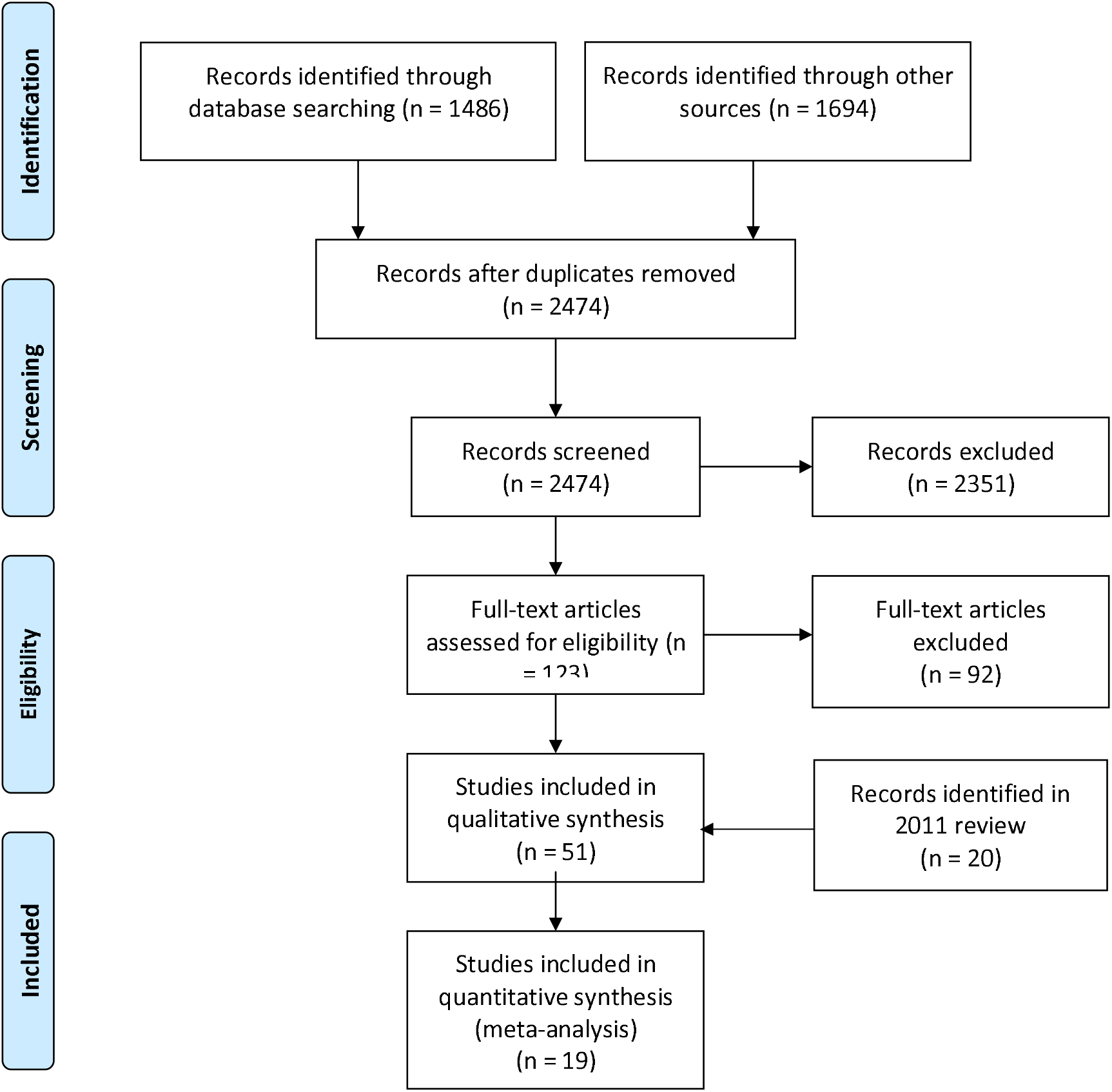
PRISMA Flow Diagram.

### Risk of bias

Reporting of sequence generation and allocation concealment was poor in 30% to 50% of studies across the categories of intervention comparisons. Due to the nature of the interventions being compared, blinding of treatment allocation after randomisation was rarely achieved. Most outcomes were assessed by study participants. This meant that outcome assessment was not blinded and therefore at high risk of bias. Some studies had laboratory-confirmed outcomes which we considered were likely to be at low risk of bias. We found no evidence of selective reporting of outcomes within the included studies. We believe publication bias is unlikely, as the included studies demonstrated a range of effects, both positive and negative, over all sizes of study. Risk of bias assessment for individual studies are shown on forest plots (Figures 2, 3, S1, S2 and S3). Figure 4 presents an overall summary of risk of bias for included studies.

**Figure 2:**
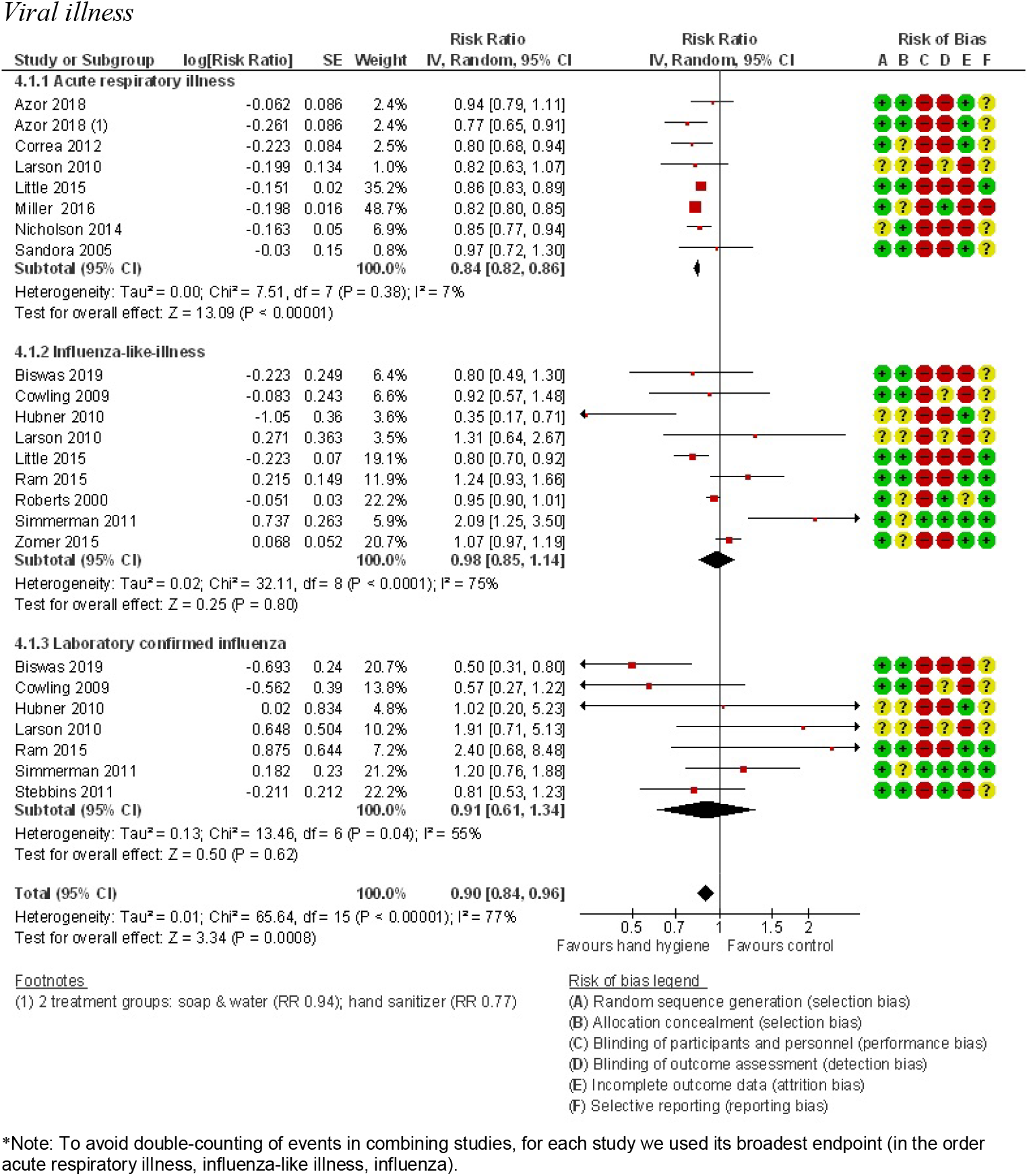
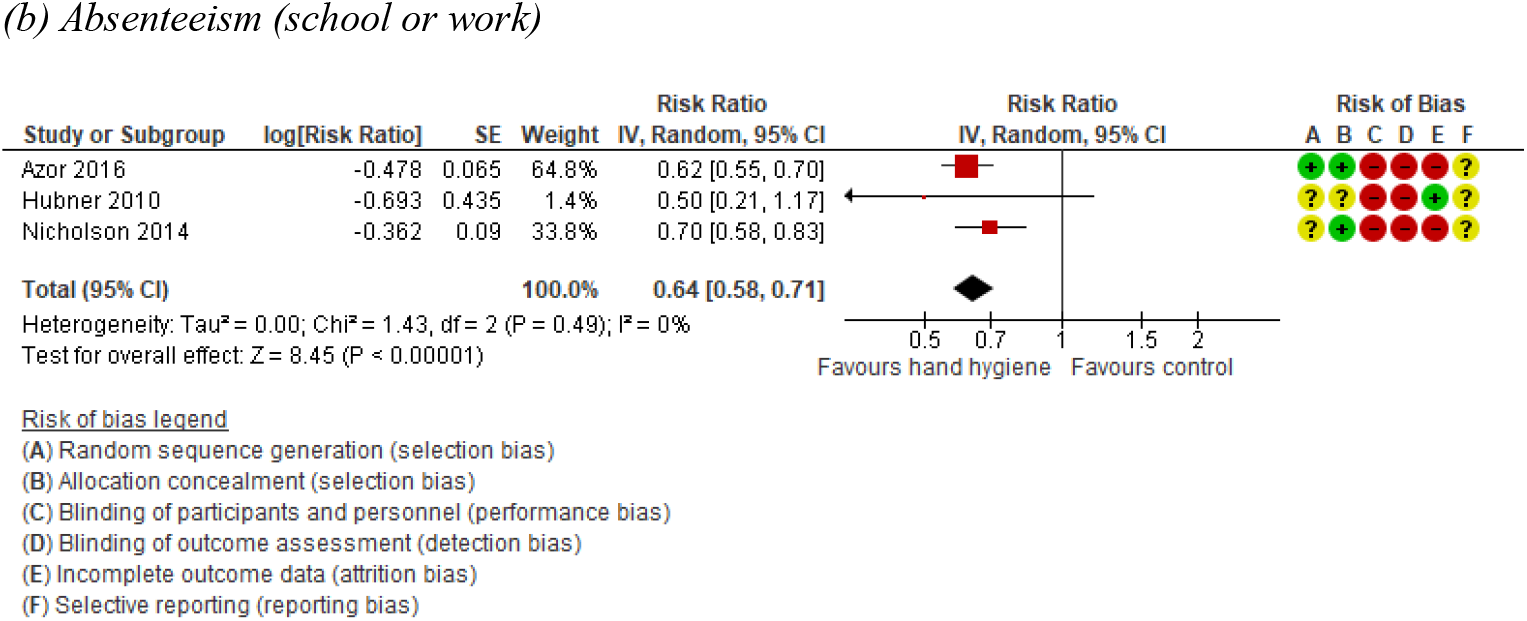
Meta-analysis of all trials comparing Hand hygiene vs control: effect on: (a) acute respiratory illness, influenza-like illness and laboratory-confirmed influenza and (b) absenteeism. *Note: To avoid double-counting of events in combining studies, for each study we used its broadest endpoint (in the order acute respiratory illness, influenza-like illness, influenza).

**Figure 3:**
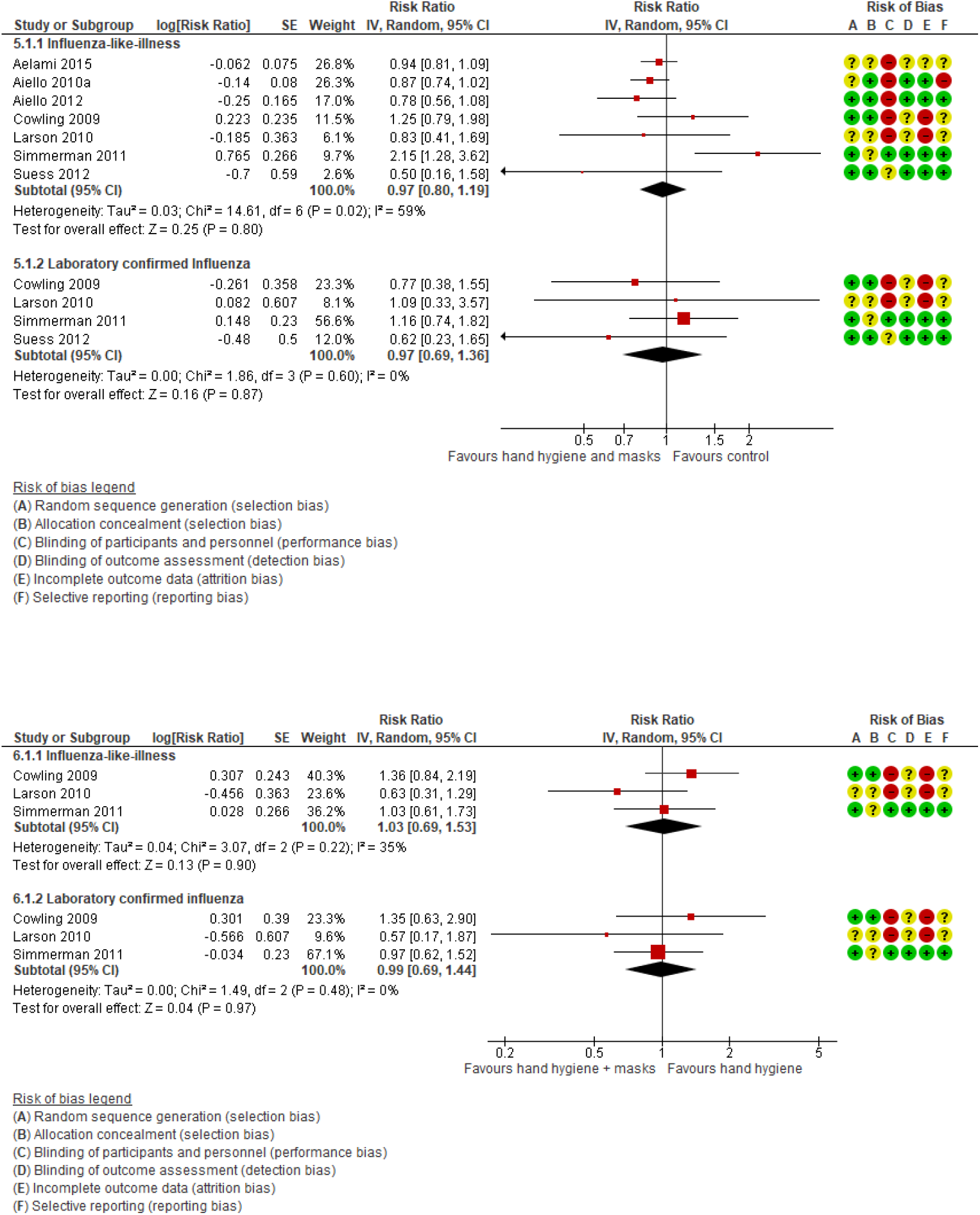
(a): Meta-analysis of trials comparing hand hygiene + masks vs control for influenza-like illness and laboratory-confirmed Influenza. (b): Meta-analysis of trials comparing hand hygiene + masks vs hand hygiene for influenza-like illness and laboratory-confirmed Influenza.

**Figure 4:**
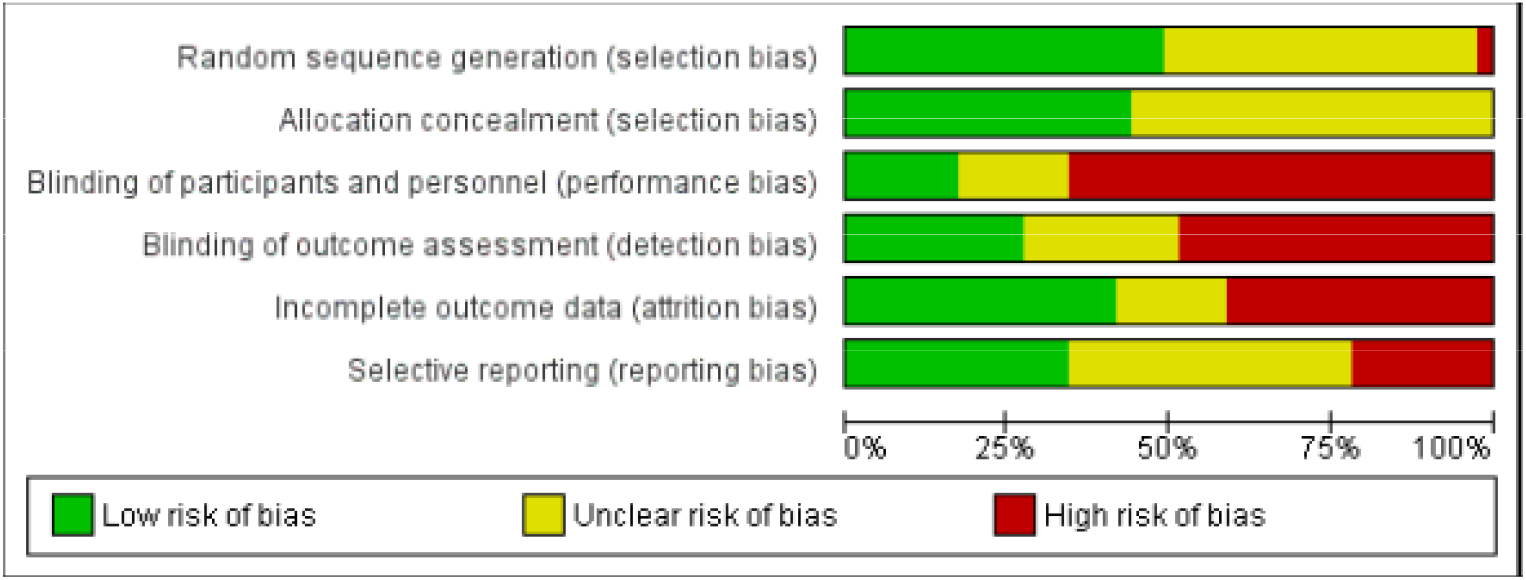
Summary of Risk of Bias (RoB) in the Included Studies.

### Hand hygiene versus control

Fifteen trials compared hand hygiene interventions with control [14-29] and provided sufficient data for meta-analysis. Populations included adults, children and families, in settings such as schools, childcare centres, homes, and offices. None were conducted during pandemics, although a few studies were conducted during peak influenza seasons. Table 1 presents characteristics of the included trials. Table 2 presents interventions investigated in the trials. Viral illness outcome definitions as reported by authors are in Table 3.

**Table 1:** Overview of characteristics of included studies.

| Study | Study period | Population | Comparison (see Table 2 for details of interventions) | Baseline use of intervention | Reported outcomes (see Table S1 for details of definitions) | Results | Adherence |
| --- | --- | --- | --- | --- | --- | --- | --- |
| **HAND HYGIENE** |  |  |  |  |  |  |  |
| Alzaher 2018; c-RCT Saudi Arabia | January to March 2018 | 496 Girls aged 6-12 yo in 4 primary schools in Riyadh | HW workshop and posters vs usual practice | Not reported | % Absence days due to URI | 0.39% and 0.72% in intervention group schools; 0.86% and 1.39% in control schools | Not reported |
| Arbogast 2016; c-RCT USA | February 2014 to March 2015 | 1386 Employees ≥18yo in 3 facilities of a health insurance company in Ohio | Hand sanitiser + wipes + hand foam vs none; both groups received education + signage about HW | > 50% of control carry HS, use HS at work and public places, 30% use HS at home, 40% carry HS throughout the day (Figure 4) | 1: Health insurance claims for preventable illnesses per employee<br>2: Absences per employee | 1: 0.30 claims in intervention; 0.37 in control (27% relative reduction, p=0.03)<br>2: 1.45 in intervention; 1.53 in control (5.0% relative reduction in intervention, p=0.30) | Estimated: per employee per day: HS 1.8 to 3.0 times; soap 2.1 to 4.4 times. Wipes at desk 1.4 to 1.5 times per week. |
| Azor-Martinez 2016;* RCT Spain | October 2009 to May 2010 | 1341 children 4-12 yo in 5 Primary schools in Almeria Province | Handwashing with soap & water plus hand sanitiser vs usual handwashing practices | Both groups used hand sanitizer before study started (C=10.7%, I=11.8%) | % absence days due to URI | 1.15% in intervention; 1.68% in control. Significantly lower in intervention (p<0.001) | Not reported, but mentioned children who washed hands correctly showed 11% decrease in URIs |
| Azor-Martinez 2018*; c-RCT Spain | November 2013 to June 2014 | 911 Children 0-3 yo in day care centres; excluded children with chronic illness or immune-suppressant medication in metropolitan Almeria | Educational and Hand hygiene (HH) with soap & water or HH with sanitiser vs usual hand-washing procedures. | All groups attended 1-hr workshop on hand-washing practices | 1: RI incidence rate ratio (primary);<br>2: Percentage difference in absenteeism days | 1: HH soap vs control 0.94 (95%CI 0.82 to 1.08); HH sanitiser vs control 0.77 (95%CI 0.68 to 0.88); HH soap vs HH sanitiser 1.21 (95%CI 1.06 to 1.39);<br>2: HH soap 3.25% vs control 4.2% (p<0.001); HH sanitiser 3.9% vs control 4.2% (p=0.026); HH soap 3.25% vs HH sanitiser 3.9% (p<0.001) | Compliance not measured. Estimated that each child used hand sanitiser 6-8 times per day |
| Biswas 2019*; c-RCT Bangladesh | June to September 2015 (influenza season) | 10,855 students aged 5 to 10 yo in 24 Primary schools (number not reported) in Dhaka | Hand sanitiser and respiratory hygiene education and cough/sneeze hygiene vs no intervention | Baseline HW events were similar between groups, both groups have equal% of schools implementing hand hygiene curriculum, less in control group have curriculum on respiratory hygiene | 1.ILI incidence rate (at least 1 episode);<br>2.Laboratory-confirmed influenza | 1: 22 per 1000 student-weeks in intervention; 27 per 1000 student-weeks in control, not statistically significantly different.<br>2: 3 per 1000 student-weeks in intervention; 6.2 per 1000 student-weeks in control, p=0.01 | HW observed; sanitiser used in 91% of events, 4.3mL per child per day; cough/sneeze etiquette observed in 33% of intervention and 2% of control group |
| Correa 2012*; c-RCT Colombia | 16 April to 18 December 2008 (three school terms) | Children aged up to 5y in 42 child-care facilities in six towns in Bogotá and five neighbouring towns | Alcohol-based handwash in addition to handwashing vs usual handwashing practice | Hand hygiene (HH) infrastructure is similar between groups | 1: ARIs in 3<sup>rd</sup> trimester of follow-up ($\geq 2$ of the following symptoms $\geq 24$ hours, for $\geq 2$ days: runny, stuffy, or blocked nose or noisy breathing; cough; fever, hot sensation, or chills; and/or sore throat. Ear pain was considered an ARI. | 1: Hazard ratio for intervention to control 0.93 (95%CI 0.57 to 0.83) | Median hand soap-water (HSW) frequency at trial end in control centers was 3 times/day. From start to end of trial, median number of applications per child rose from 3.5 to 4.5 in preschools and from 3.5 to 5.5 in community centers |
| Cowling 2009*; c-RCT Hong Kong | January to September 2008 | 407 adults with influenza A/B and 794 household members in 259 households In Hong Kong | HH (136 households); facemask + hand hygiene (137 households); education (134 control households) | Not reported | 2: Secondary attack ratio<br>3: Laboratory-confirmed influenza ILI definition 1<br>4: ILI definition 2 | 2: HH 5; HH+masks 7; control 10<br>3: HH 16; HH+masks 21; control 19<br>4: HH 4; HH+masks 7; control 5 | At the final home visit, the intervention groups reported higher adherence to the interventions than the control group |
| DiVita 2011 (conference abstract); RCT Bangladesh | 2009-2010 | 274 index case patients and their household members in rural setting | Handwashing stations with soap and motivation vs none | Not reported | 1: Secondary attack rate (SAR) for laboratory-confirmed influenza<br>2: SAR for ILI | 1: SAR higher in intervention group (11.0% vs 7.5%)<br>2: SAR higher in intervention group (14.2% vs 11.9%) | Not reported |
| Feldman 2016; c-RCT Israel | May to September 2014 | Ships in a naval base (697 sailors) | Hand disinfection + soap and water installed vs none | Not reported | 1: Number of respiratory infections;<br>2: Number of off-duty days | 1: 11 in each group<br>2: 112 in intervention; 104 in control | Disinfectant mean 8.2mL per sailor per day |
| Gwaltney 1980; RCT USA | November 1978 and January 1979 | Healthy volunteers experimentally exposed to rhinovirus at University of Virginia, Charlottesville | Virucidal hand wash vs placebo | Not reported | 1: Number with illness after immediate exposure;<br>2: Number with illness after 2-hour delay in exposure (Illness defined as symptom score $\geq 5$ and nasal discharge) | 1: 0 of 8 in intervention; 7 of 7 in control<br>2: 1 of 10 in intervention; 6 of 10 in control | Not reported |
| Hubner 2010*; RCT Germany | March 2005 to April 2006 | 1230 Public administration employees, at Ernst-Moritz-Arndt University Greifswald | Hand disinfection provided vs none | Study recruited those who do not already apply hand disinfection at work | Odds ratios (95% CI) (intervention:control)<br>1: Influenza<br>2: Common cold<br>3: Sinusitis<br>4: Sore throat<br>5: Fever<br>6: Cough | 1: 1.02 (0.20 to 5.23)<br>2: 0.35 (0.17 to 0.71)<br>3: 1.87 (0.52 to 6.74)<br>4: 0.62 (0.31 to 1.25)<br>5: 0.38 (0.14 to 0.99)<br>6: 0.45 (0.22 to 0.91) | Mean hand disinfection frequency >5 times/d in 19%, 3-5 time/d in 59.8%, 1-2 times/d in 20.5% per person month |
| Ladegaard 1999; RCT Denmark | Not reported? | 475 children 0-6 years in day care centers | Hand hygiene and education | Not reported | Sick days during the 'effect period' | 2.22 days/child in the intervention group vs 3.36 days/ child in the control group | Not reported |
| Larson 2010*; c-RCT USA | November 2006 to July 2008 | 509 primarily Hispanic households with at least 3 people and a preschool or elementary school child in New York city | 1 education; 2 education with alcohol-based hand sanitizer; 3 education with hand sanitizer and face masks | Not reported | Incidence rate ratios (episodes per 1000 person weeks) for:<br>1: URI<br>2: ILI<br>3: Influenza<br>4: Secondary attack rates<br>URI/ILI/influenza<br>5: ILI/influenza | 1; HS 29; HS+masks 39; control 35<br>2: HS 1.9; HS+masks 1.6; control 2.3<br>3: HS 0.6; HS+masks 0.5; control 2.3<br>4: HS 0.14; HS+masks 0.12; control 0.14<br>5: HS 0.02; HS+masks 0.02; control 0.02 | Compliance was poor: Gp-2 used HS occasionally (44.2%), gp-3 used HS 1-2 times within the previous 24 hr (56.9). Gp-2 used a mean of 11.6 ounces HS/month, while gp-3 used a mean of 12.1 ounces/month of HS. Compliance with mask was poor (two masks/day/ILI episode (range: 0-9)) |
| Little 2015*; RCT England | Jan 17, 2011, to March 31, 2013 | 20,066 Adults $\geq 18$ yo in the community | Bespoke automated web-based motivational | Not clearly reported | Number of episodes of RTIs in index participants (risk ratio) | Risk ratio for intervention to control | Questionnaire response rate was 84% |
|  |  | recruited through GP practices | intervention, with tailored feedback, vs none |  |  | 0.86 (95%CI 0.83 to 0.89, p<0.001) |  |
| Luby 2005; RCT Pakistan | April 2002 to April 2003 | Households in 36 Squatter settlements in Karachi | Antibacterial soap or plain soap and education about HW provided weekly vs none | At baseline all households purchased an average of 1 bar hand soap per week. | 1: Cough or difficulty breathing in children <15yrs (episodes/100 person-weeks);<br>2: Congestion or coryza in children <15yrs (episodes/100 person-weeks);<br>3: Pneumonia in children <5yrs (episodes/100 person-weeks) | All comparisons significantly lower than control<br>1: 4.21 in antibacterial soap group; 4.16 in plain soap group; 8.50 in control<br>2: 7.32 in antibacterial soap group; 6.87 in plain soap group; 14.78 in control<br>3: 2.42 in antibacterial soap group; 2.20 in plain soap group; 4.40 in control | Not reported, but soap bar consumption increased (3.3 soap bar/week) |
| Millar 2016*: c-RCT USA | May 2010 to Jan 2012 | Around 30,000 healthy males, army trainees (18-42yo) in Georgia | Standard promotion of handwashing, enhanced promotion, and promotion plus a once-weekly application of chlorhexidine-based body wash | Not reported | Incidence rates of ARI over 20 months | 37.7 HH; 29.3 enhanced; 35.3 standard; RR for HH to standard 1.07 (95%CI 1.03 to 1.11); RR for enhanced to HH 0.78 (95%CI 0.75 to 0.81) | Not reported |
| Morton 2004; RCT-Crossover study USA | Not reported | 253 elementary school children from 17 classrooms in Northern New England | Alcohol gel plus education vs regular hand washing | Not reported | Absence due to infectious illness | Results not stated numerically | Not reported |
| Nicholson 2014*: c-RCT India | October 22 2007 to August 2 2008 | Children 5 yo in low-income urban communities in Mumbai | Combination hand washing promotion with provision of free soap | Not reported | Target children:<br>1: Episodes of ARI (per 100 person weeks);<br>2: School absence | 1: 16 in intervention; 19 in control<br>2: 1.2 in intervention; 1.7 in control<br>3: 10 in intervention; 11 in control | Soap consumption to be 45g per household/week in control compared with 235g in intervention households |
|  |  |  |  |  | Families:<br>3: Episodes of ARI |  |  |
| Pandejpong 2012;<br>c-RCT<br>Thailand | December 2009 to February 2010 | 1437 preschool-age children in a kindergarten in Bangkok | Alcohol handgel every hour or every 2 hours or once before lunch (3 groups) | Not reported | Absent days due to confirmed ILI/present days | 0.017 in the every hour group; 0.025 in every 2 hours group; 0.026 in before lunch group. Statistically significant difference between every hour group and before lunch group, and between every hour and every two hours groups | Not reported |
| Priest 2014;<br>c-RCT<br>New Zealand | April to September 2009 | In 68 primary schools in cities of Christchurch, Dunedin, Invercargill | Hand hygiene education for all and hand sanitiser in intervention group | Baseline self-reported overall family hand hygiene was 87.1% in control, 84.7% in HS group. | 1: % Absence days due to respiratory illness;<br>2: % Absence days due to any illness | 1: 0.84% in intervention group; 0.80% in control (p=0.44)<br>2: 1.21% in intervention group; 1.16% in control (p=0.35) | Average hand sanitiser solution dispensed per child in the intervention schools was 94 ml (SD19). |
| Ram 2015*;<br>RCT<br>Bangladesh | June 2009 to December 2010 | Index case-patients with ILI (fever with cough or sore throat) who were only symptomatic person in their household (rural) | Promoting intensive handwashing in households to prevent transmission of ILI | Not reported | 1: Secondary attack ratio for intervention to control for ILI;<br>2: Laboratory-confirmed influenza | 1: 1.24 (95% CI 0.93 to 1.65)<br>2: 2.40 (95%CI 0.68 to 8.47) | Median per capita soap consumption of 2.3g (interquartile range: 1.7 to 3.2g) in the first 12 days of enrolment |
| Roberts 2000*;<br>c-RCT<br>Australia | March and November 1996 | Children aged ≤3yo attending 23 child-care centres at least 3 days a week in Australian Capital Territory | Handwashing programme with training for staff and children | Not reported | Incidence rate ratio for ARI (runny nose, cough and blocked nose) | IRR 0.92 for intervention to control (95%CI 0.86 to 0.99) | HW group 1 compliance rate 53%–69% HW group 2 compliance rate 70%–79%; HW group 3 compliance rate over 80% |
| Sandora 2005*;<br>c-RCT<br>USA | November 2002 to April 2003 | 292 families with 1 or more children 6 months to 5 yo who were in childcare for 10 or more hours a week in | Using a hand sanitiser (HS) and a programme of instruction on the transmissions of GI infections and ARIs in families. | Baseline use of HS: 36% in the control, 41% in intervention group. Control families were asked not to use hand | Incidence rates for ARI (episodes per person month) (2 of the following symptoms for 1 day or 1 of the following | 0.43 in intervention;<br>0.42 in control | HS median use 5.2 times/day IRR in family consuming >2 oz HS per 2weeks vs. ≤2oz, IRR 0.81 (95% CI: 0.65–1.09; P .06) |
|  |  | Massachusetts neighbourhoods |  | sanitizer during the study period | symptoms for 2 days: runny nose, cough, sneezing, stuffy or blocked nose, fever, sore throat). An illness episode had to be separated by 2 symptom-free days from a previous episode |  |  |
| Savalainen 2012; c-RCT Finland | November 2008 to May 2010 | 683 people in 21 office work units in 6 corporations in Helsinki | Hand hygiene with soap and water (IR1 group), or with alcohol-based hand rub (IR2 group) or control (none); intervention groups also received education | Not clearly reported | 1: Number of respiratory infection episodes/week;<br>2: Number of reported infection episodes/week;<br>3: Number of reported sick leave episodes/week | 1: 0.076 in IR1; 0.085 in IR2; 0.080 in control, NS<br>2: 0.097 in IR1; 0.107 in IR2; 0.104 in control, NS<br>3: 0.042 in IR1; 0.035 in IR2; 0.035 in control. Significantly higher in IR1 compared with control | Soap or disinfectant usage per participant was: 6.1 in IR1; 6.9 in IR2; control: not reported |
| Simmerman 2011*; c-RCT Thailand | April 2008 to August 2009 | 442 index children with 1147 household members recruited in a Bangkok Paediatric outpatient department | Hand washing (HW), or hand washing plus paper surgical face masks (HW + FM) or control (none) | Not clearly reported | Odds ratios for secondary attack rates for influenza | OR for HW:control 1.20 (95%CI 0.76 to 1.88)<br>OR for HW+masks:control 1.16 (95%CI 0.74 to 1.82) | Hand washing episodes/day on day 7: control: 3.9; HW: 4.7; HW+FM: 4.9 |
| Stebbins 2011*; c-RCT USA | Jan 2007 to April 2008 influenza season | 3360 children in 10 Pittsburgh elementary schools | Training in hand and respiratory (cough) hygiene. Hand sanitizer was provided and encouraged to be used regularly | Not reported | Incidence rate ratios for intervention to control for<br>1: Laboratory-confirmed Influenza (RT-PCR) ;<br>2: Influenza-A;<br>3: Absence | 1: IRR 0.81 (95%CI 0.54 to 1.23)<br>2: 0.48 (95%CI 0.26 to 0.87)<br>3: 0.74 (95%CI 0.56 to 0.97) | Not reported |
| Talaat 2011;<br>c-RCT<br>Egypt | February 16 to<br>May 12 2008 | 44,451 children in<br>60 elementary<br>schools in Cairo | Children in the<br>intervention schools<br>were required to wash<br>hands twice daily, and<br>health messages were<br>provided through<br>entertainment activities | At baseline, schools<br>no soap or hand-drying material.<br>Hand washing, if<br>done at all, was only<br>performed by<br>rinsing hands in<br>water. | 1: Number of absence<br>days due to ILI;<br>2: Number of absence<br>days | 1: 917 in intervention;<br>1,671 in control<br>($p<0.001$)<br>2: 13,247 in<br>intervention; 19,094 in<br>control ($p<0.001$) | Not reported |
| Temime<br>2018;<br>c-RCT<br>France | April 2014 to<br>April 2015 | 26 nursing homes in<br>Paris | Hand hygiene with<br>alcohol-based handrub,<br>promotion, staff<br>education, and local<br>work groups vs none | Baseline quantity of<br>consumed handrub<br>solution in NHs was<br>4.5 mL per resident<br>per day and did not<br>differ between<br>control and<br>intervention groups | Incidence rate of acute<br>respiratory infection<br>clusters (5 or more<br>people in same nursing<br>home) | 2 ARI clusters in<br>intervention; 1 in control | Not reported |
| Turner 2004;<br>RCT<br>Canada | Not reported | Healthy volunteers' response to<br>application of<br>rhinovirus on hands;<br>85 in study 1 and<br>122 in study 2 in<br>Winnipeg, Manitoba | Study 1: ethanol vs<br>salicylic acid 3.5% or<br>salicylic acid 1%;<br>Study 2: skin cleanser<br>wipe vs ethanol |  | % of volunteers<br>Infected with<br>rhinovirus | 7% in each intervention<br>group; 32% in control<br>(study 1)<br>22% in intervention,<br>30% in control (study 2) | Not reported |
| Turner 2012;<br>RCT<br>USA | 9 weeks in Fall<br>2009 | 212 Healthy<br>volunteers from<br>University of<br>Virginia >18 years | Antiviral hand<br>treatment vs no<br>treatment | Not reported | 1: Number of<br>rhinovirus infections;<br>2: Common cold<br>infections<br>3: Rhinovirus-<br>associated illnesses | 1: 49 in intervention; 49<br>in control, NS<br>2: 56 in intervention; 72<br>in control, NS<br>3: 26 in intervention; 24<br>in control, NS | All subjects (100%)<br>applied at least 90% of<br>the expected amount of<br>hand treatment |
| Zomer<br>2015*;<br>c-RCT<br>Netherlands | September<br>2011 to April<br>2012 | 545 Children aged 6<br>months and 3.5yrs at<br>start of the trial in<br>71 day care centres<br>in Rotterdam,<br>Gouda and Leiden | Four components:<br>1. HH products, paper<br>towel dispensers, soap,<br>alcohol-based hand<br>sanitizer & hand cream<br>provided for 6 months<br>2. Training & booklet | At baseline,<br>compliance in<br>intervention DCCs<br>was 53% vs. 63% in<br>control DCCs (OR<br>0.59, 95% CI 0.37–<br>0.94). | 1: Incidence rate ratio<br>for intervention to<br>control for common<br>cold ;<br>2: The common cold<br>was defined as a<br>blocked or runny nose<br>with at least 1 of the<br>following symptoms: | 1: IRR 1.07 (95%CI<br>0.97 to 1.19)<br>2: 8.2 episodes per child<br>year in intervention; 7.4<br>episodes per child year<br>in control | At 6 months follow-up,<br>compliance was 59%<br>vs. 44%, respectively<br>(baseline-corrected OR<br>4.13, 95% CI 2.33–<br>7.32) |
|  |  |  | 3. 2 team training sessions aimed at HH improvement<br>4. Posters and stickers for caregivers and children as reminders |  | coughing, sneezing, fever, sore throat, or earache |  |  |
| Yeung 2011; c-RCT Hong Kong | April to November 2007 | Staff in aged care facilities in Hong Kong | Alcohol-based hand gel + materials + education vs control (basic life support workshop) | Soap and water hand hygiene used at baseline | Difference between pre-study period and post-study in pneumonia infections recorded in residents | 0.63/1000 reduction in intervention group; 0.16/1000 increase in control | Overall HH adherence was 33.3% |
| **HAND HYGIENE AND MASKS** |  |  |  |  |  |  |  |
| Aelami 2015 (conference abstract); RCT Saudi Arabia | 2012 Hajj season | 664 Iranian pilgrims during Annual Hajj to Makkah | Education on personal hygiene including a hygienic package with alcohol-based handrub (gel or spray), surgical masks, soap, paper handkerchiefs, and user instructions versus none | Not reported | Proportion with ILI (defined as presence of ≥ two of the following during their stay: fever, cough, and sore throat) | 52% in intervention; 55.3% in control (p<0.001) | Not reported |
| Aiello 2010*; C-RCT USA | Nov 2006 to Mar 2007 | 1297 university students living in residence halls at University of Michigan | Residence halls were randomly assigned to 1 of 3 groups—face mask use, face masks with hand hygiene, or control— for 6 weeks. | 55% of study participants own HS. All arms received HH education | 1: ILI (defined as cough and ≥1 constitutional symptom (fever, chills, or body aches)); 2: Laboratory-confirmed influenza A or B. Tested with cell cultures and RT-PCR | Significant reduction in ILI cases in both intervention groups compared with control over weeks 3 – 6. No significant differences between FM and FM+HH | Not reported |
| Aiello 2012*; C-RCT USA | 2007–2008 Influenza season | 1,111 students residing in university residence halls (N = 37) at University of Michigan | Residence halls were randomly assigned to 1 of 3 groups—face mask use (FM), face masks with hand hygiene (FM+HH), or control— for 6 weeks. | Baseline hand hygiene and hand sanitisers use was similar across all groups | 1: Clinical ILI (presence of cough and ≥1 of fever/feverishness, chills, or body aches); 2: Laboratory-confirmed influenza A or B. Throat swab | 1: Non-significant reductions in FM group compared with control over all weeks. Significant reduction in FM+HH group compared with control in weeks 3 – 6. | Mask use: FM+HH wore masks 5.08 hours per day (SD, 2.23); mask group 5.04 hours per day [SD, 2.20] HS use: FM+HH 4.49 times per day (SD, 4.10) |
|  |  |  |  |  | specimens were tested for influenza A or B using real-time polymerase chain reaction (Rt-PCR) | 2: Non-significant reductions in both intervention groups compared with control. | M only:1.29 times per day (SD, 1.77); Control: 1.51 times per day (SD, 2.25) |
| Cowling 2009*; c-RCT Hong Kong | January 2 to September 30, 2008 | 407 patients with influenza A/B and 794 household members in 259 households | 3 groups: 1 Control = lifestyle measures; 2 control plus enhanced hand hygiene; 3 control plus enhanced hand hygiene (HH) plus facemasks (HH+mask) | Not reported | 1: Secondary attack ratio (SAR);<br>2: Laboratory-confirmed influenza;<br>3: ILI definition 1;<br>4: ILI definition 2 | 1: HH 5; HH+masks 7; control 10<br>2: HH 16; HH+masks 21; control 19<br>3: HH 4; HH+masks 7; control 5 | Adherence to the interventions was low |
| Larson 2010*; c-RCT USA | November 2006 to July 2008 | 509 primarily Hispanic households with at least 3 people and a preschool or elementary school child in upper Manhattan | 1: education (control); 2: education with alcohol-based hand sanitizer (HS); 3: education with hand sanitizer and face masks(HS+mask) | Not reported | Incidence rate ratios (episodes per 1000 person weeks) for:<br>1: URI;<br>2: ILI;<br>3: Influenza<br>Secondary attack rates (SAR for:<br>4: URI/ILI/influenza;<br>5: ILI/influenza | 1: HS 29; HS+masks 39; control 35<br>2: HS 1.9; HS+masks 1.6; control 2.3<br>3: HS 0.6; HS+masks 0.5; control 2.3<br>4: HS 0.14; HS+masks 0.12; control 0.14<br>5: HS 0.02; HS+masks 0.02; control 0.02 | Compliance was poor: Gp-2 used HS occasionally (44.2%), Gp-3 used HS 1-2 times within the previous 24 hr (56.9%). Gp-2 used a mean of 11.6 ounces HS/month,Gp-3 used a mean of 12.1 ounces/month of HS. Compliance with mask was poor (two masks/day/ILI episode (range: 0–9) |
| Simmerman 2011*; c-RCT Thailand | April 2008 to April 2009 | Households with a febrile influenza-positive child; 442 index cases and 1147 household members recruited in a Bangkok Paediatric outpatient department | Control, handwashing or handwashing plus paper surgical facemasks | Not clearly reported | Odds ratios for secondary attack rates for influenza | OR for HW:control 1.20 (95%CI 0.76 to 1.88)<br>OR for HW+masks:control 1.16 (95%CI 0.74 to 1.82) | Hand washing episodes/day on day 7: control: 3.9; HW: 4.7; HW+FM: 4.9. Mask use: average of 12 masks/person /week with a mean 211 min/mask/day |
| Suess 2012*; c-RCT Germany | Two influenza seasons 2009-2010 2010-2011 | 84 Households with influenza index case In Berlin | Facemask + Hand hygiene vs facemasks only vs none | Not clearly reported. But mentioned hand washing had been generally recommended already before the pandemic | 1: Secondary attack rates in household contact;<br>2: Laboratory-confirmed influenza;<br>3: ILI | 1: Mask 9; Mask+HH 15; control 23<br>2: Mask 9; Mask+HH 9; control 17 | Adherence was described as good for adults and children, contacts and index cases |
| **HAND HYGIENE AND SURFACE / OBJECT DISINFECTION** |  |  |  |  |  |  |  |
| Ban 2015; c-RCT China | October 2010 to September 2011 | 408 children <5yo In two Kindergartens In Xiantao city | Antibacterial products for hand hygiene, surface cleaning and disinfection provided to families and kindergartens | Not clearly reported | 1: Respiratory illness;<br>2: Cough and expectoration | 1: OR 0.47 for intervention to control (95%CI 0.38 to 0.59)<br>2: OR 0.56 (95%CI 0.48 to 0.65) | Adherence was described as high |
| Carabin 1999; c-RCT Canada | September 1 1996 to November 30 1997 | 1729 children in 47 day care centers in south central Quebec area | One off hygiene education and materials vs none | At baseline most of the kindergartens in both groups had some level of the intervention such as washing toys, use of diluted bleach to clean toys. | Difference in incidence rate for URTI (cluster-level result) | 0.28 episodes per 100 child-days lower in intervention group (95%CI 1.65 lower to 1.08 higher) | Not reported |
| Chard 2019; c-RCT Laos | 2014 to 2017 | 3993 children in 100 primary schools in Saravane Province | Complex sanitation intervention and education vs none | No schools had handwashing facilities as baseline (Chard 2018) | Pupil-reported symptoms of respiratory infection over 1 week | NS difference between groups. 29% of intervention group, 32% control, adjusted risk ratio 1.08 (95%CI 0.95 to 1.23) | Reported adherence as ratio between groups (RR 0.97 (95% CI 0.84, 1.11), fulfilling ≥75% of intervention outcomes was considered adherence. |
| Ibfelt 2015; c-RCT Denmark | December 2012 to April 2013 | 587 children aged 6 months to 3 years in 12 day-care nurseries in Copenhagen | Disinfectant washing of linen and toys by commercial company every 2 weeks vs usual care | Not reported | Presence of respiratory viruses on surfaces | Statistically significant reduction in intervention group in adenovirus, rhinovirus, RSV, metapneumovirus, but not other viruses including coronavirus | Not reported |
| Kotch 1994;<br>RCT<br>USA |  | 371 families with children in day care centers in Northern Carolina | Training in hand washing and diapering for care givers and all other staff | Washing hands after diapering (I: 79.2%, C: 87.5%). Washing toys daily (I: 45.4%, C: 56.5%) | Respiratory illness incidence rate in<br>1: children <24 mos;<br>2: children ≥24 mos | 1: 14.78 episodes per child-year in intervention; 15.66 in control<br>2: 12.87 in intervention; 11.77 in control | Not reported |
| McConeghy 2017;<br>RCT<br>USA | October 1 2015 to May 31 2016 | 5 Nursing Homes in Colorado | Staff education, cleaning products and audit of compliance and feedback vs none | Not reported | Infection rates | Upper respiratory infections not reliably recorded or reported | The number of hand-washing occasions per NH resident was steady over time but differed by treatment facility (P=.03) |
| Sandora 2008*;<br>c-RCT<br>USA | November 2002 to April 2003 | 292 families with children attending 26 child-care centers in Avon-Ohio | Hand sanitiser and hand hygiene education materials supplied biweekly vs materials about good nutrition (control) | HS in home was 47% in control, 51% in intervention | Absence due to respiratory illness (multivariable analysis) | Rate ratio 1.10 for intervention to control (95%CI 0.97 to 1.24) | Not reported but HS use per classroom per week was 1.25 bottles |
| White 2001<br>DB-RCT<br>USA | March to April 1999 | 769 children age 5-12 years from one private and two public elementary schools in California. | Participants were blinded to either hand rub with benzalkonium chloride or placebo, batches of 4 colour-coded bottles containing both. | All children attended a 22-minute on proper hand washing technique and cough/sneezing behaviour (4-min video) | ARI symptoms, laboratory testing of<br>Laboratory: testing of virucidal and bactericidal activity of the product | 30% to 38% decrease of illness and absenteeism (RR for illness absence incidence 0.69, RR for absence duration 0.71) | Reported that Large portion of the original study participants were lost due to a lack of compliance |
| **OTHER (MISCELLANEOUS) INTERVENTIONS** |  |  |  |  |  |  |  |
| Hartinger 2016;<br>c-RCT<br>Peru | September 2008 to January 2010 | 534 children 51 rural communities in San Marcos province | Cooking and sanitation provision and education vs none | Not reported | Number of ARI episodes per child-year | NS difference between groups. Risk ratio for intervention to control 0.95 (95% CI 0.82, 1.10) | Compliance with SODIS was 60% with a steady decline throughout follow-up reaching 10% at end of study. |
| Huda 2012;<br>c-RCT<br>Bangladesh | 2007 to 2009 (18 months) | 1692 children in (unknown number of) household clusters | Sanitation provision and education vs none | Baseline handwashing 28% | Respiratory illness | 12.6% in intervention group, 13.0% in control group. Not adjusted for multiple outcome measurements. No CIs reported | Improved frequency of handwashing in all groups |
| Najnin 2019; c-RCT Bangladesh | September 24 2011 to August 31 2013 | 5877 households with 39,089 people in 60 geographic areas in Dhaka | Sanitation and behaviour change intervention (plus cholera vaccine) vs none | Presence of water, soap, or ash in handwashing location (47%). Handwashing at baseline was low | Respiratory illness in past 2 days | 2.8% in intervention group, 2.9% in control. Adjusted risk ratio for intervention to control 0.82 (95%CI 0.69 to 0.98) | Uptake of the intervention increased during the study |
| Satomura 2005; RCT Japan | December 2002 to March 2003 | 387 subjects at 18 sites across Japan | Water gargling (WG) vs. povidone-iodine gargling (IG) vs. control. Frequency of gargling was 3 times/day for 60 days | All subject kept usual handwashing routine (measured at end as WG 6.5 times/d, IG 6.2/d control 6.2/d | Incidence of first upper respiratory tract infection (URTI)<br>Severity of URTI symptoms | Incidence rate of first URTI as episodes/30 person-days was 0.17 WG, 0.24 in IG and 0.26 in control subjects. IR ratios against controls were 0.64 (95%CI 0.41– 0.99) and 0.89 (95%CI 0.60 –1.33) | None of the two gargling groups skipped gargling |
| **VIRUCIDAL TISSUES** |  |  |  |  |  |  |  |
| Farr 1988; c-RCT USA Trial 1 and Trial 2 | 1983-1986 | 186 Charlottesville, Virginia families in trial 1 and 98 in trial 2 | Trial 1: Virucidal nasal tissues vs placebo vs none; Trial 2: Virucidal nasal tissues vs placebo | Not reported | Respiratory illnesses per person over 24 weeks<br>Trial 1;<br>Trial 2 | Trial 1: 3.4 in tissues group; 3.9 in placebo; 3.6 in no tissues; Trial 2: 3.4 in tissues group; 3.6 in placebo. NS differences | Families not using tissues regularly were excluded from analysis |
| Longini 1988; DB-PC RCT USA | August 1984 to March 1985 | 296 households in Tecumseh, Michigan | Virucidal nasal tissues vs placebo | Not reported | Secondary attack rate of viral infections (number of infections in household members of index case) | 10.0 in intervention, 14.3 in placebo, NS. | Virucidal tissue group 82% using 'all the time' vs 71% in placebo group |

\*: Studies marked with \* have been pooled in meta-analysis
NS: Not significantly different; yo: years old; mos: months; SAR: Secondary attack rate; ILI: Influenza-like illness; HH: hand hygiene; HS: hand sanitiser; HW: handwashing
**Colour codes:** Hand hygiene only (yellow), Both hand and masks (orange), Hand hygiene + surface / materials disinfection (green), Other / Complex (blue) and Virucidal tissues (grey)

**Table 2.**
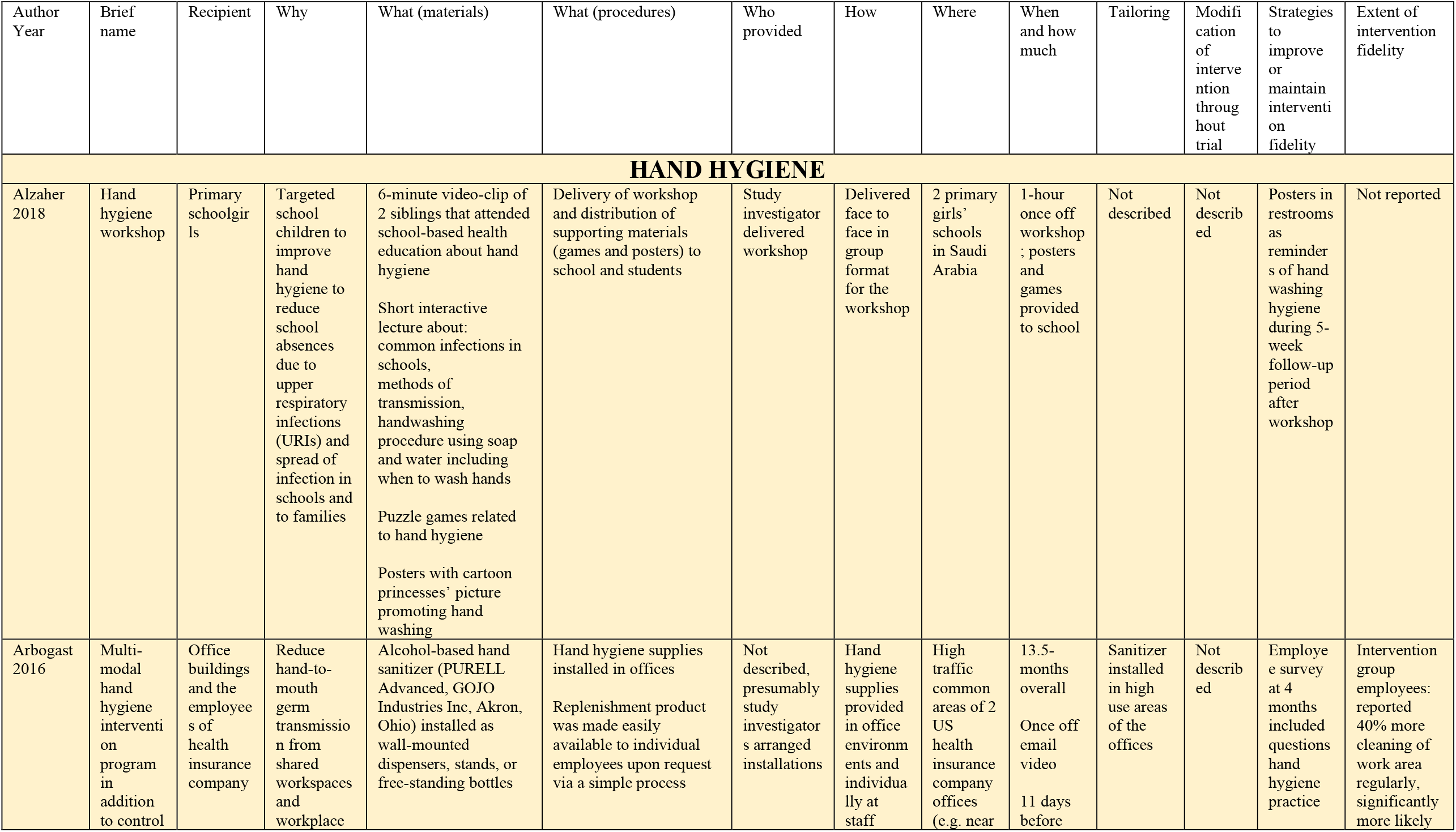
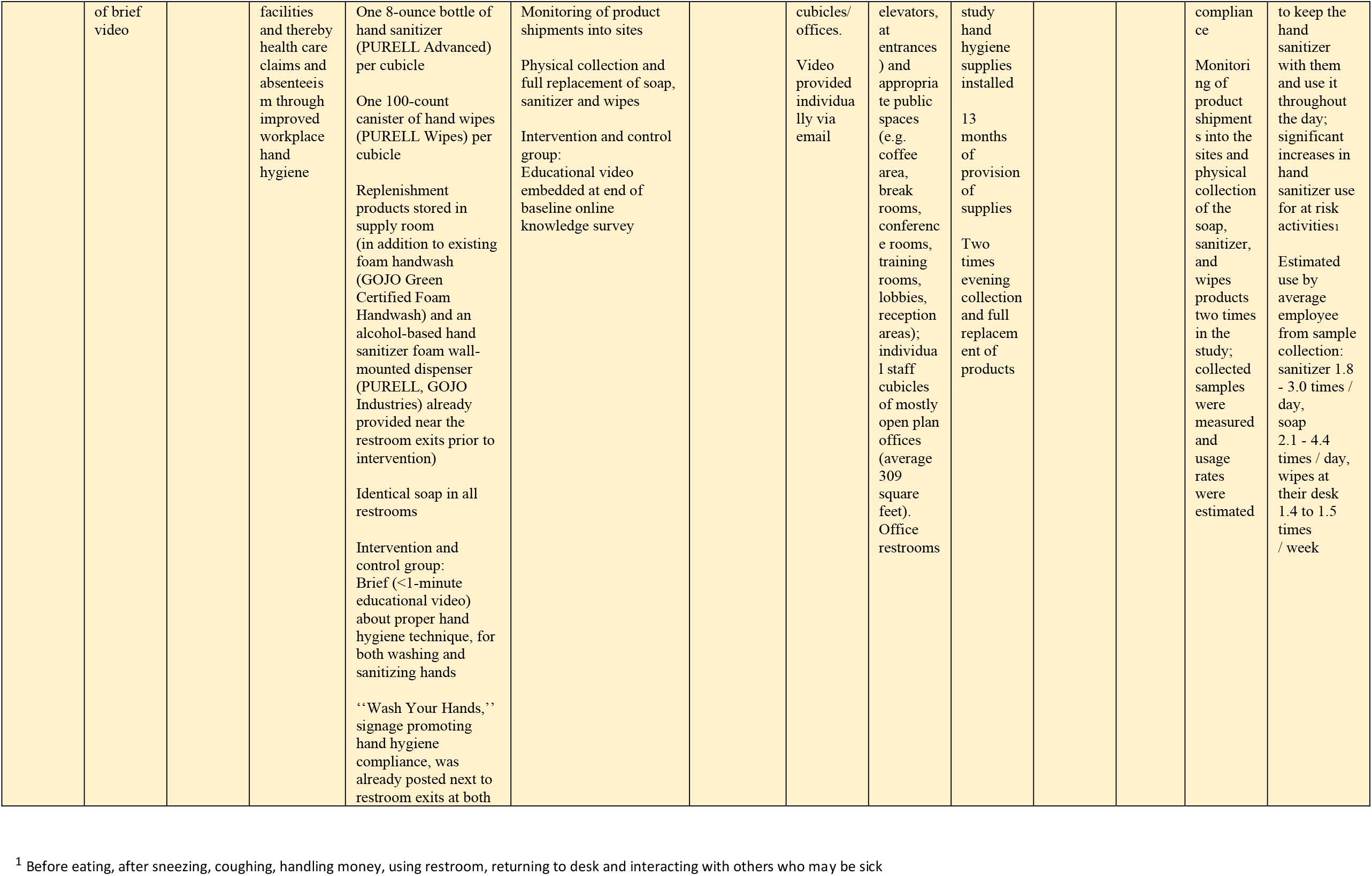
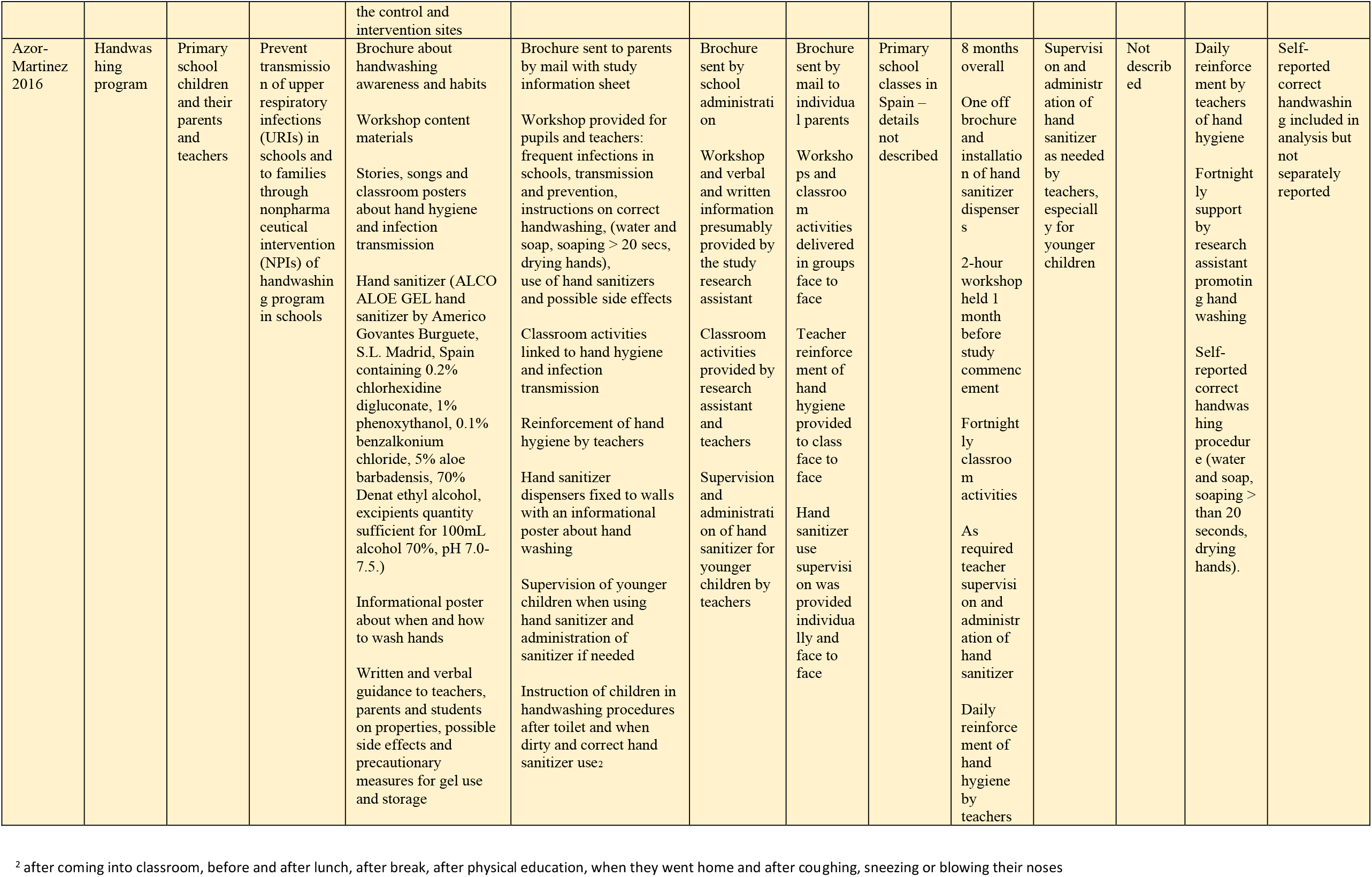
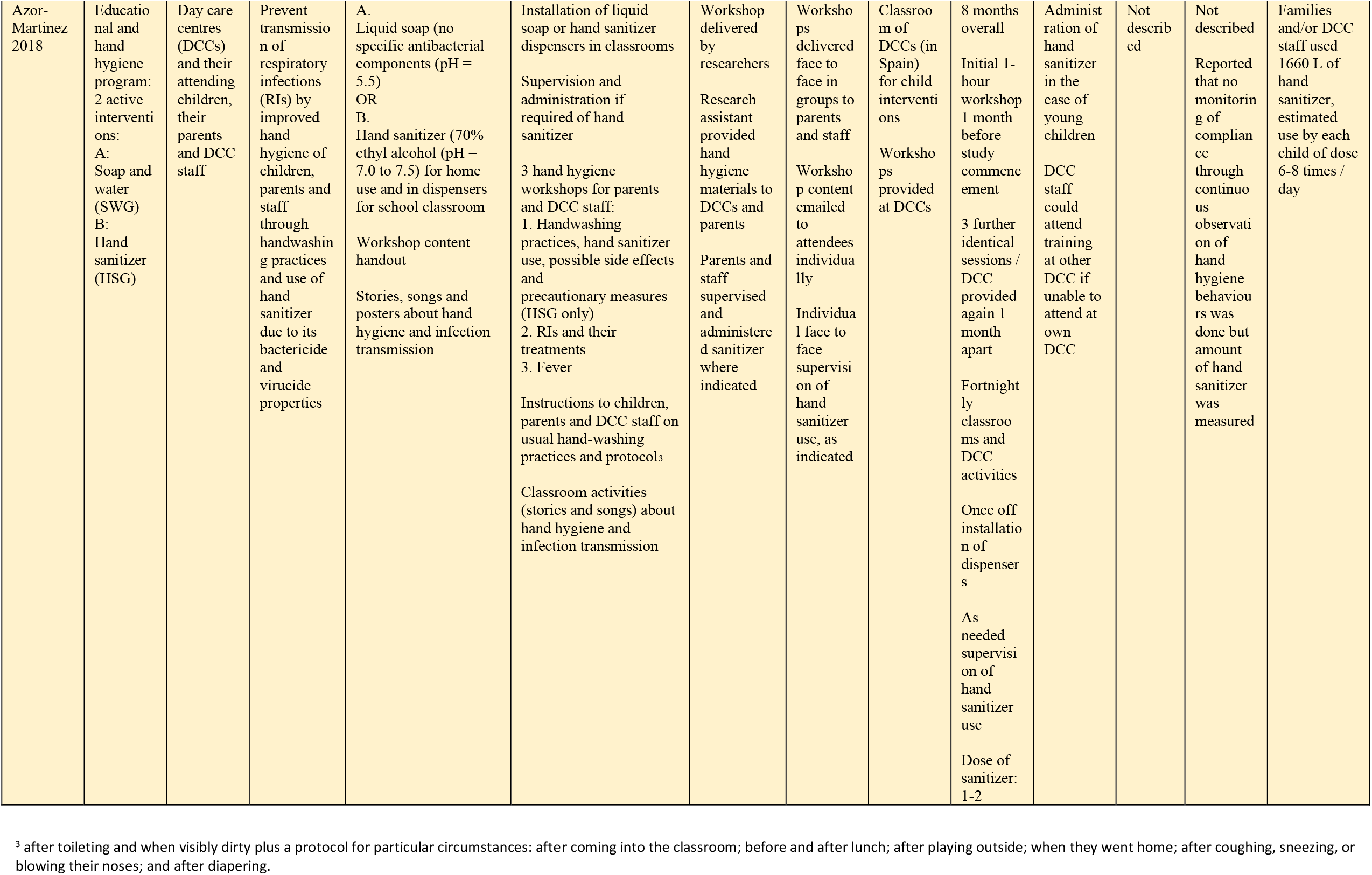
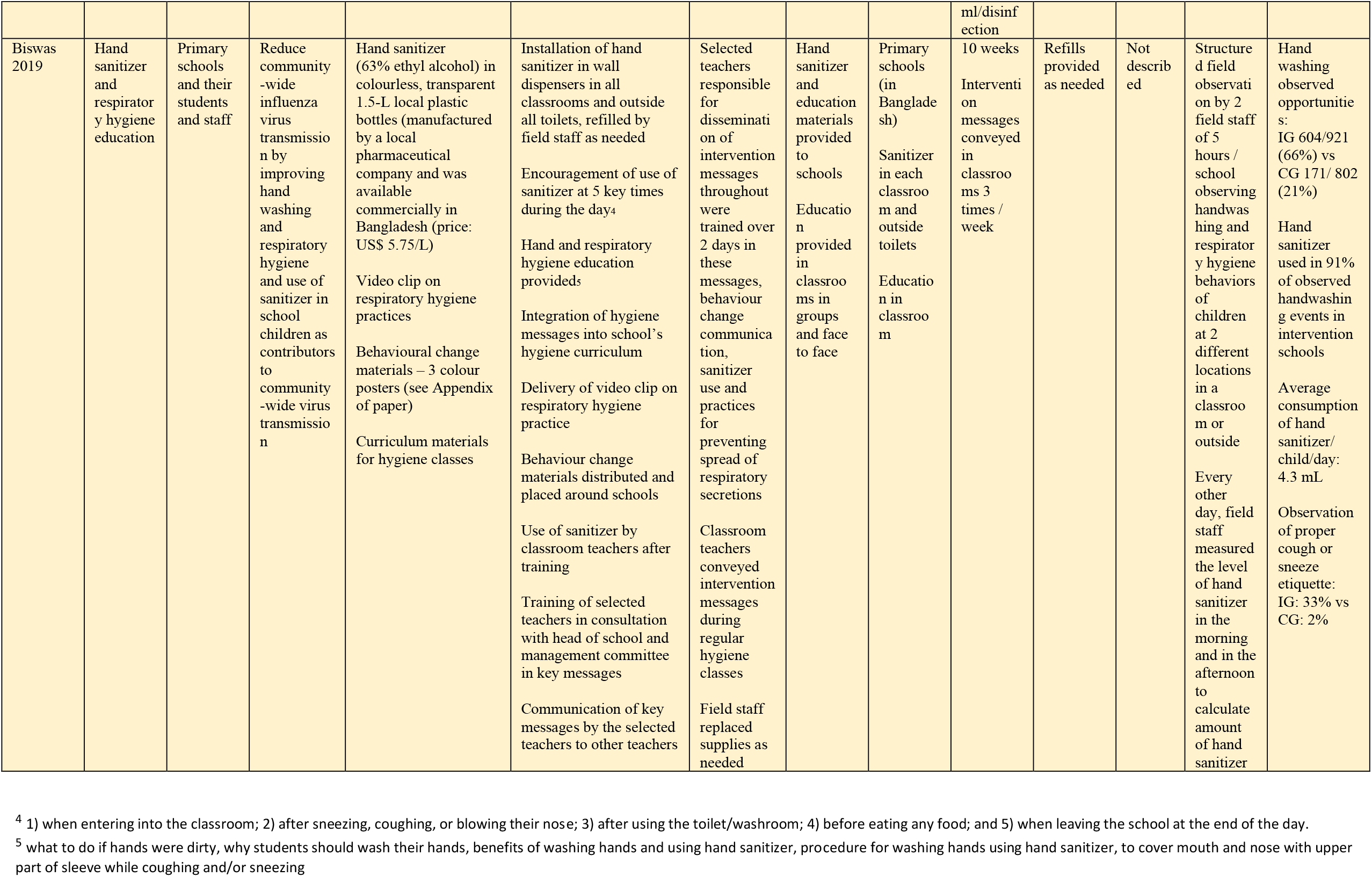
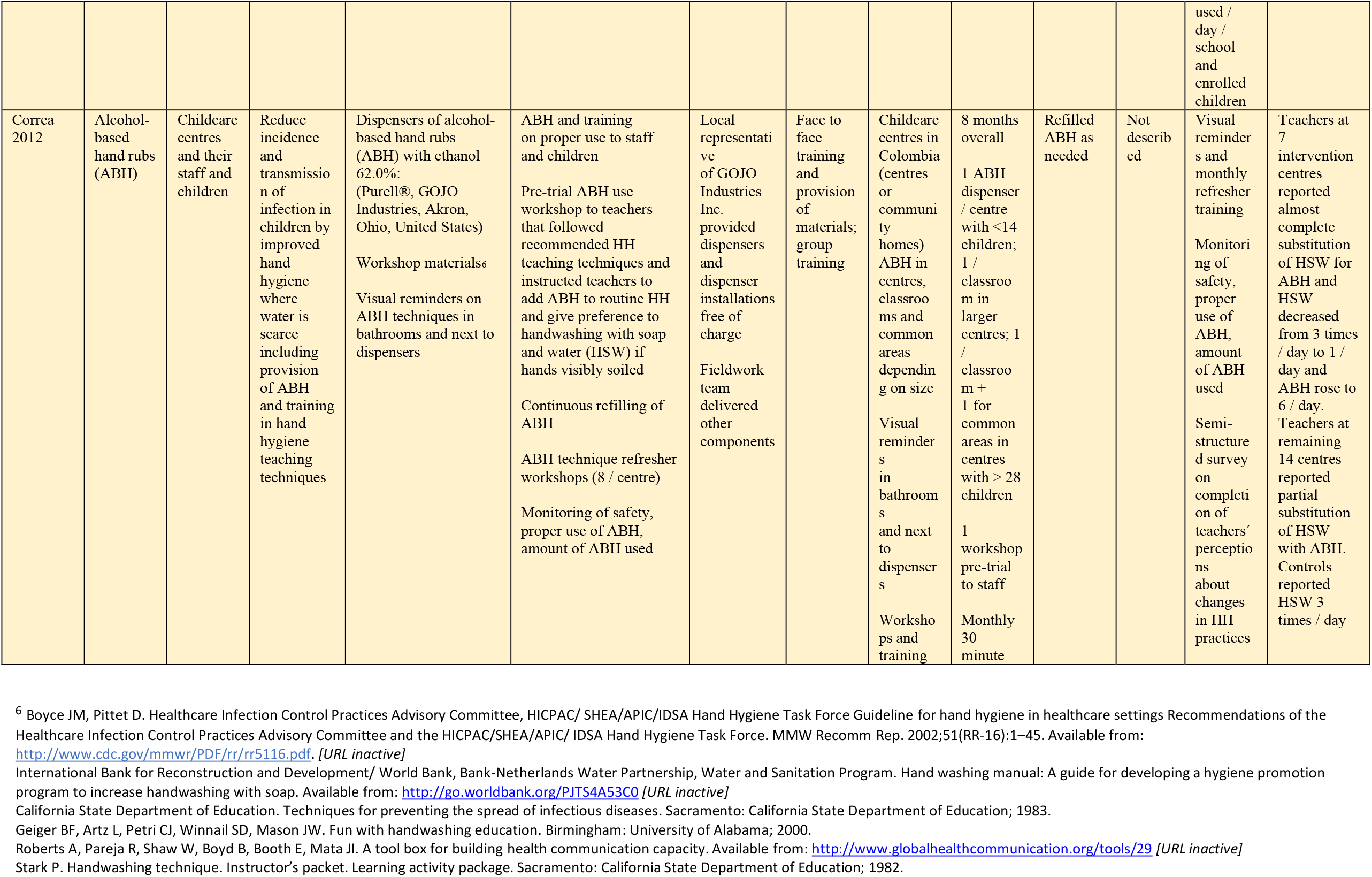
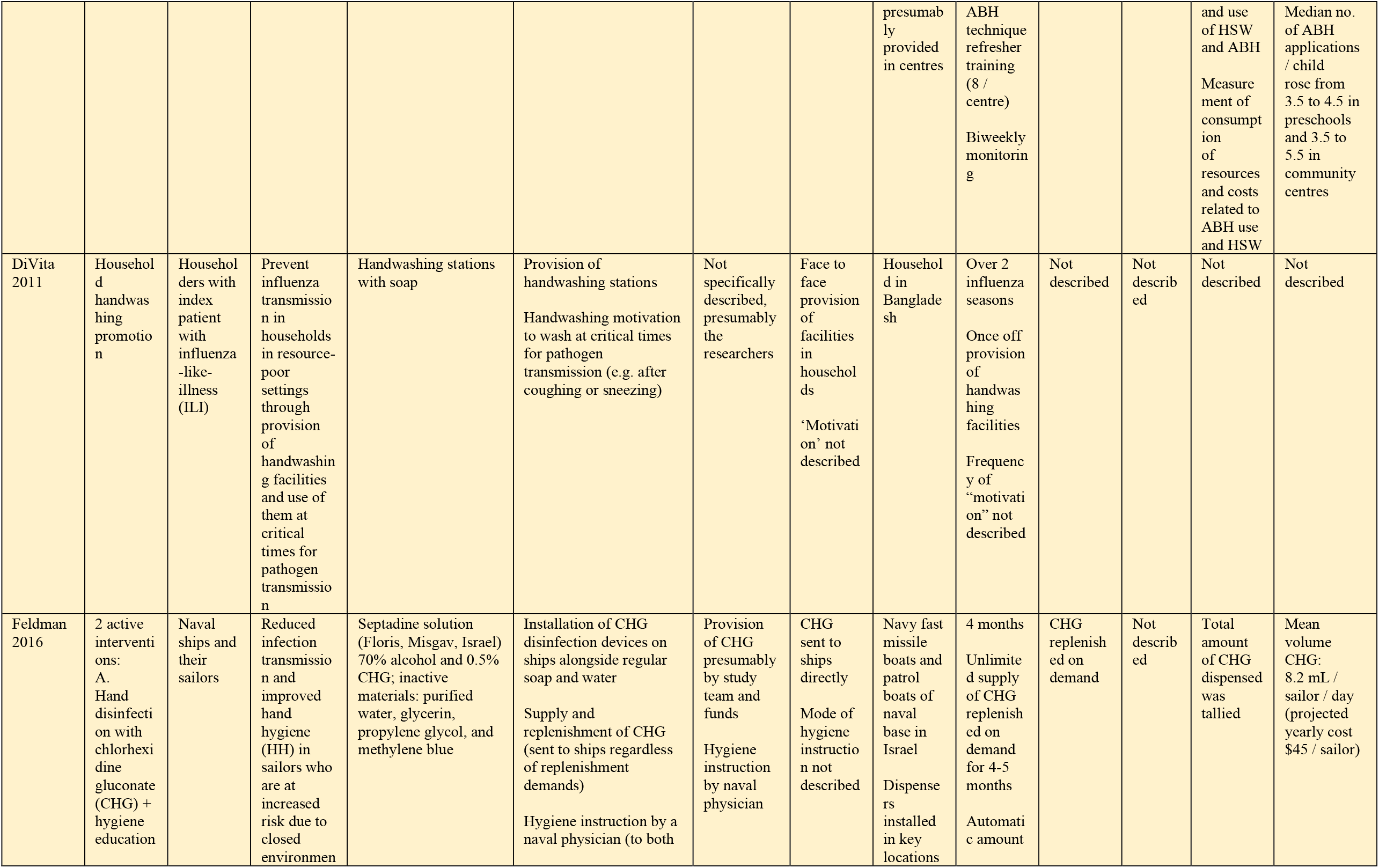
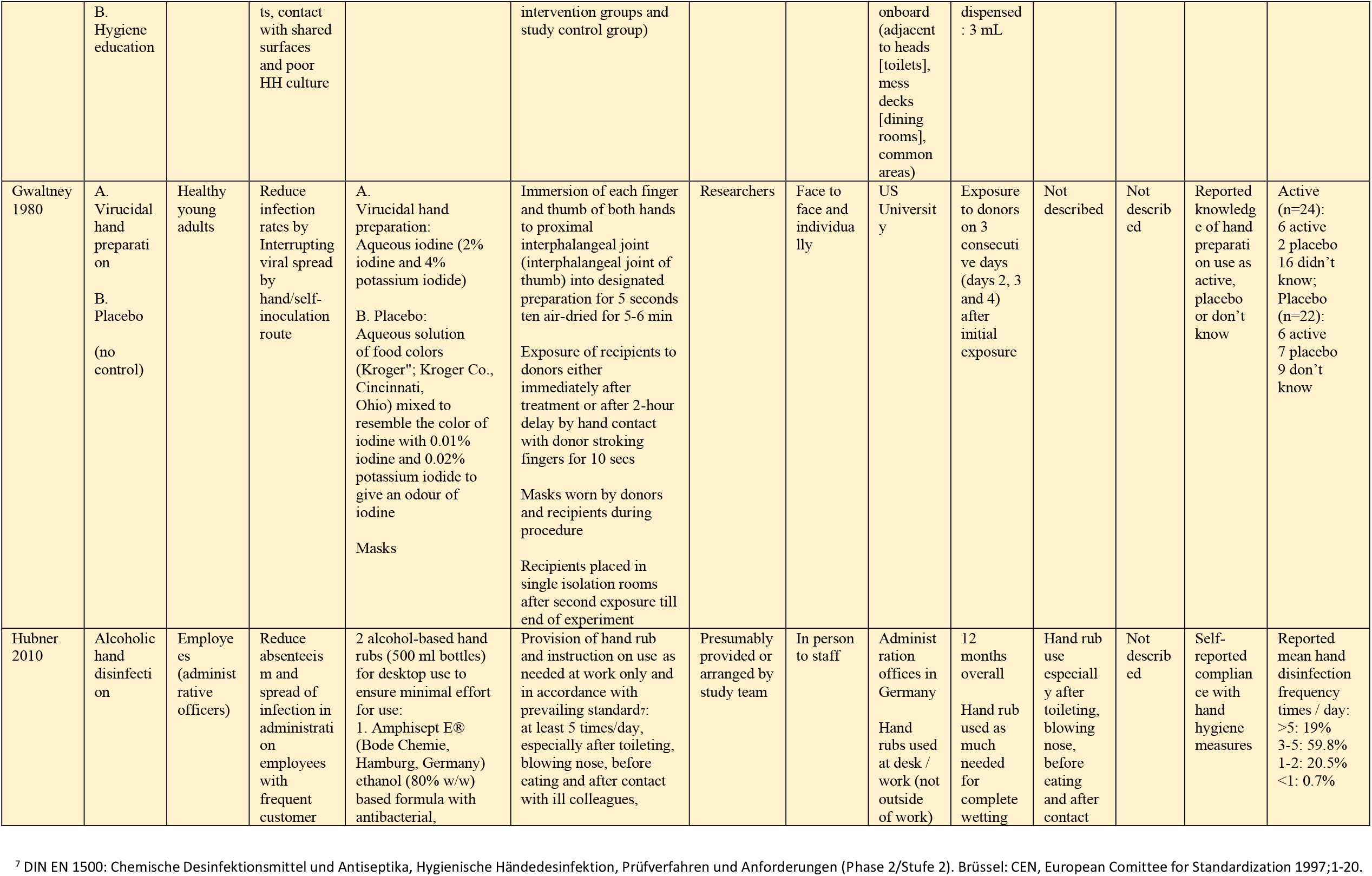
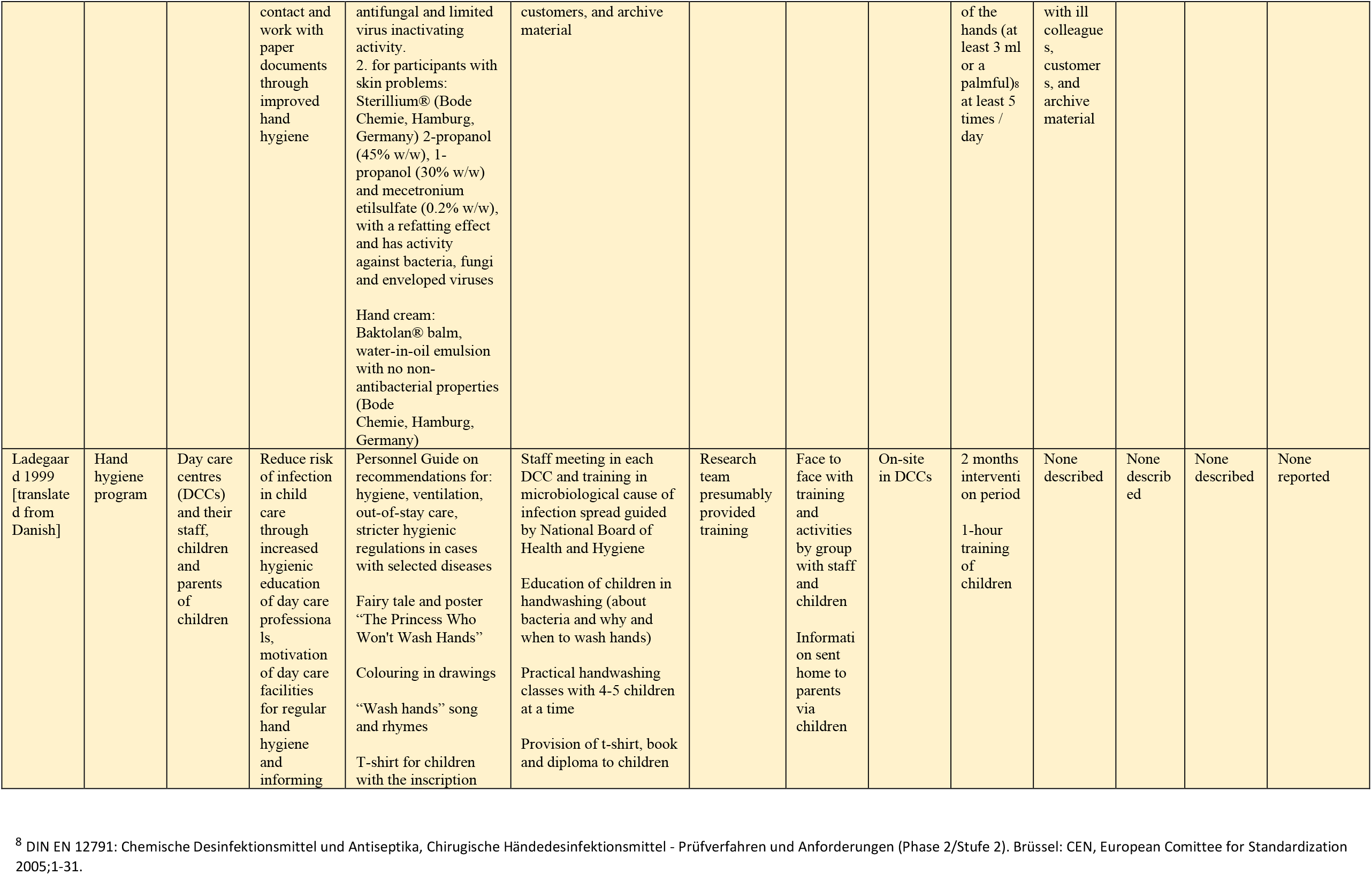
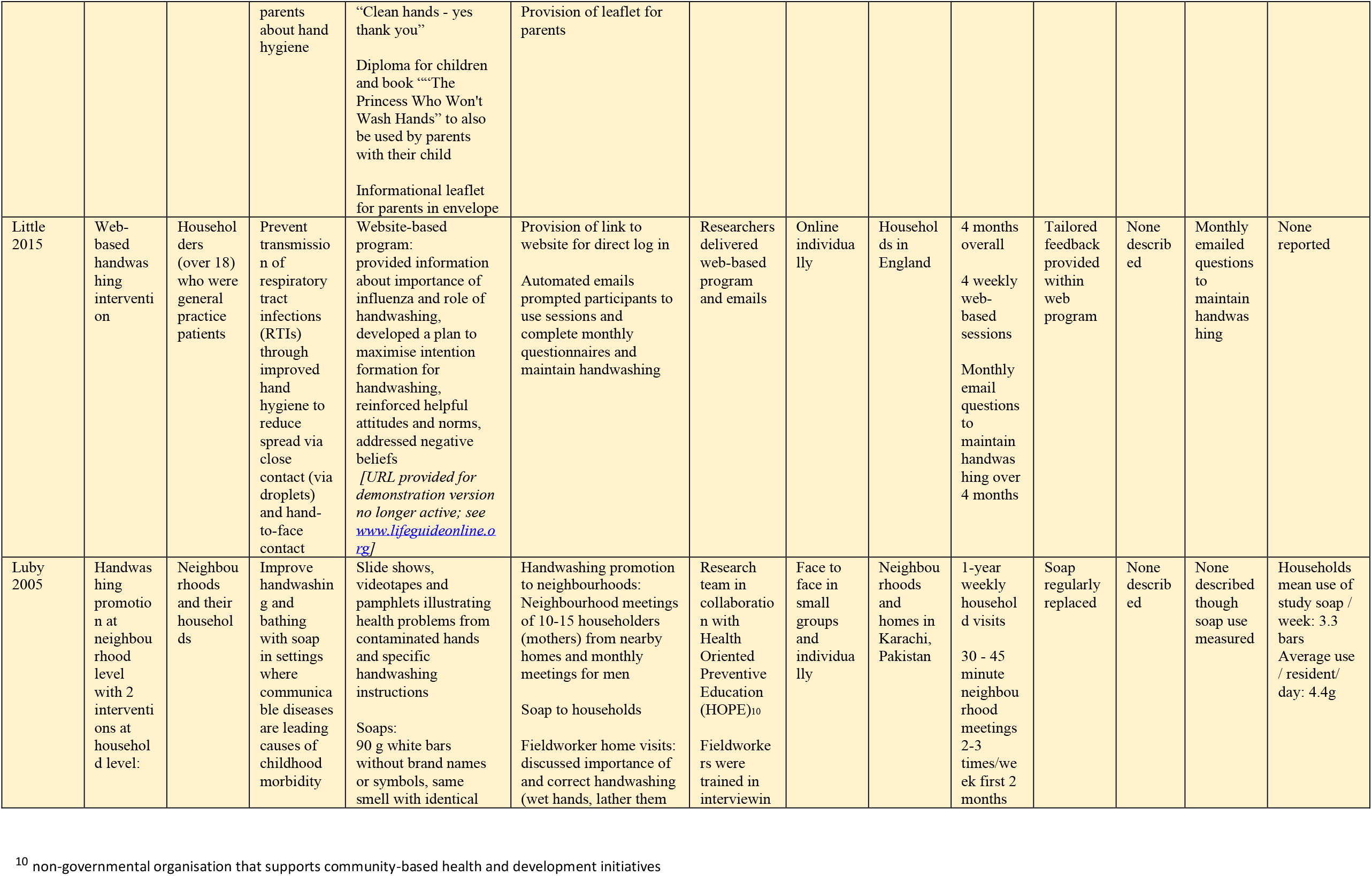
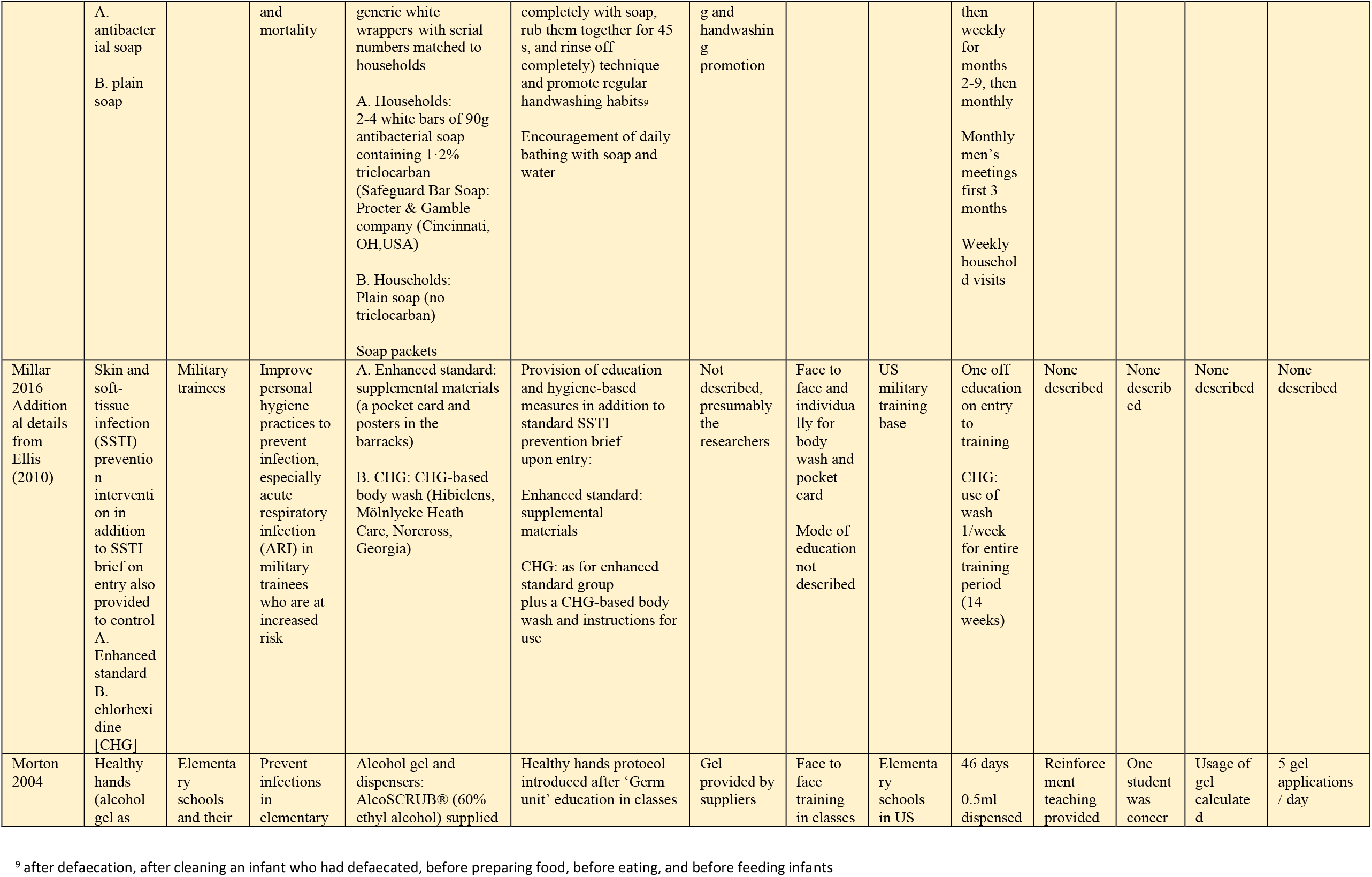
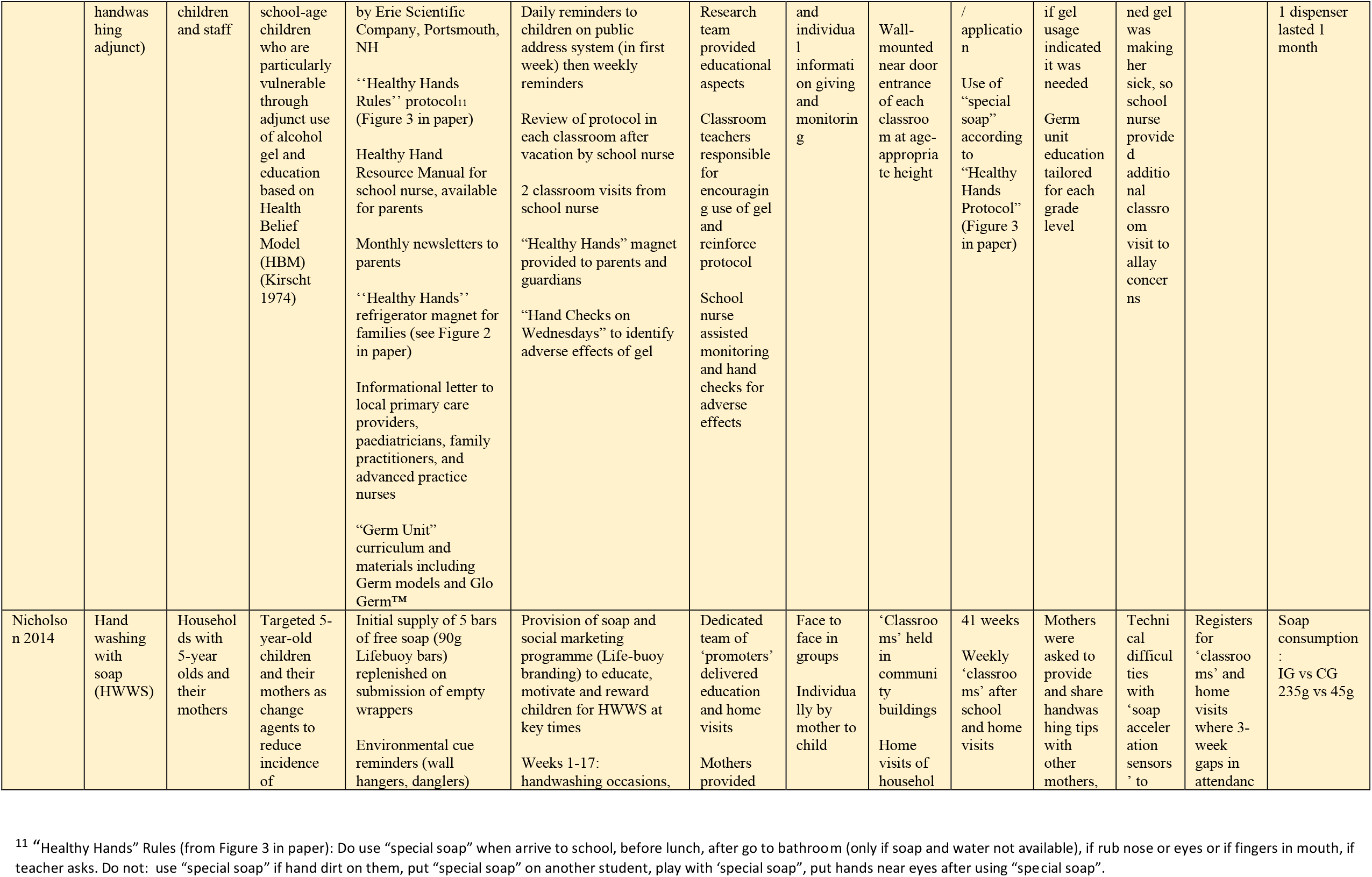
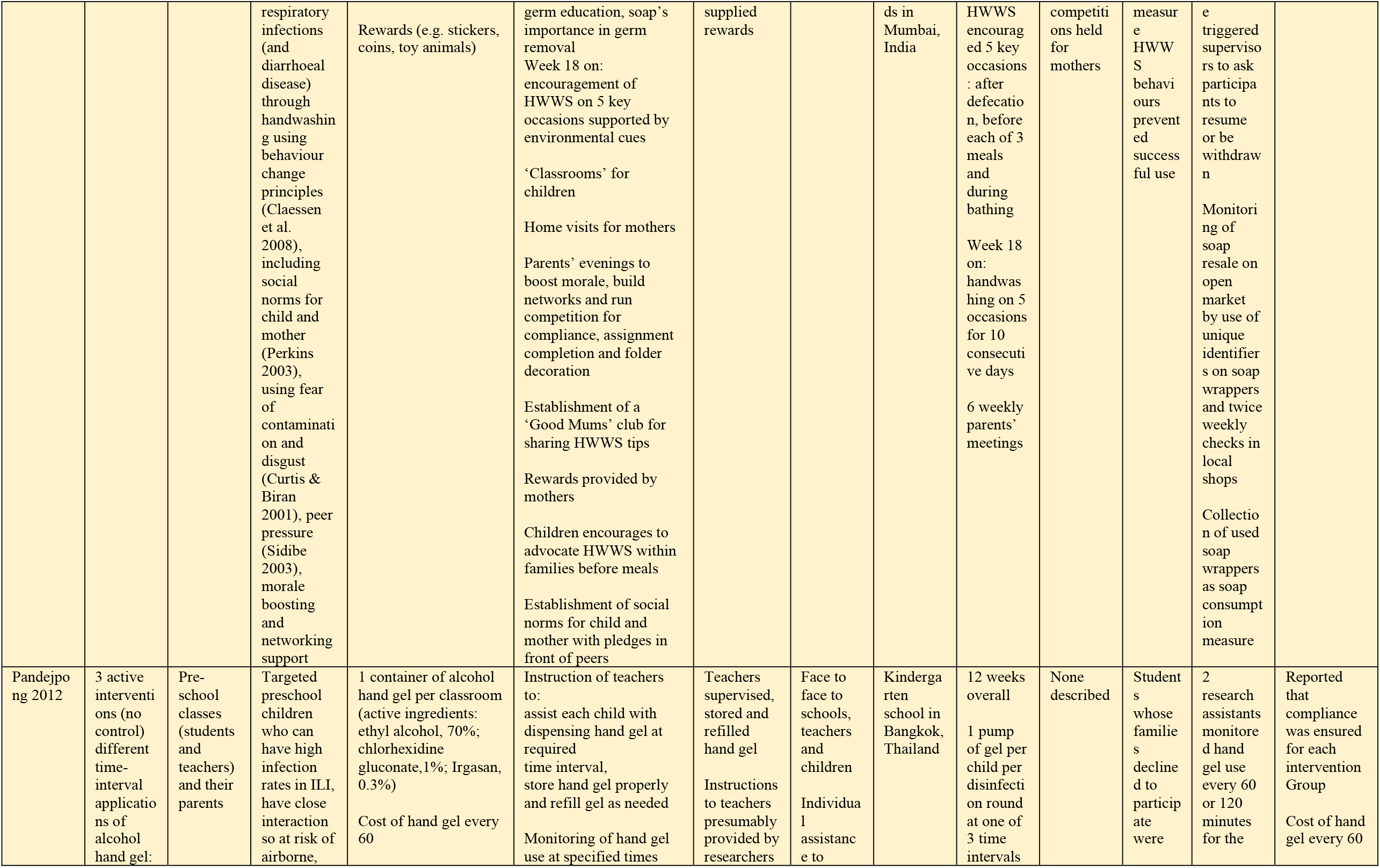
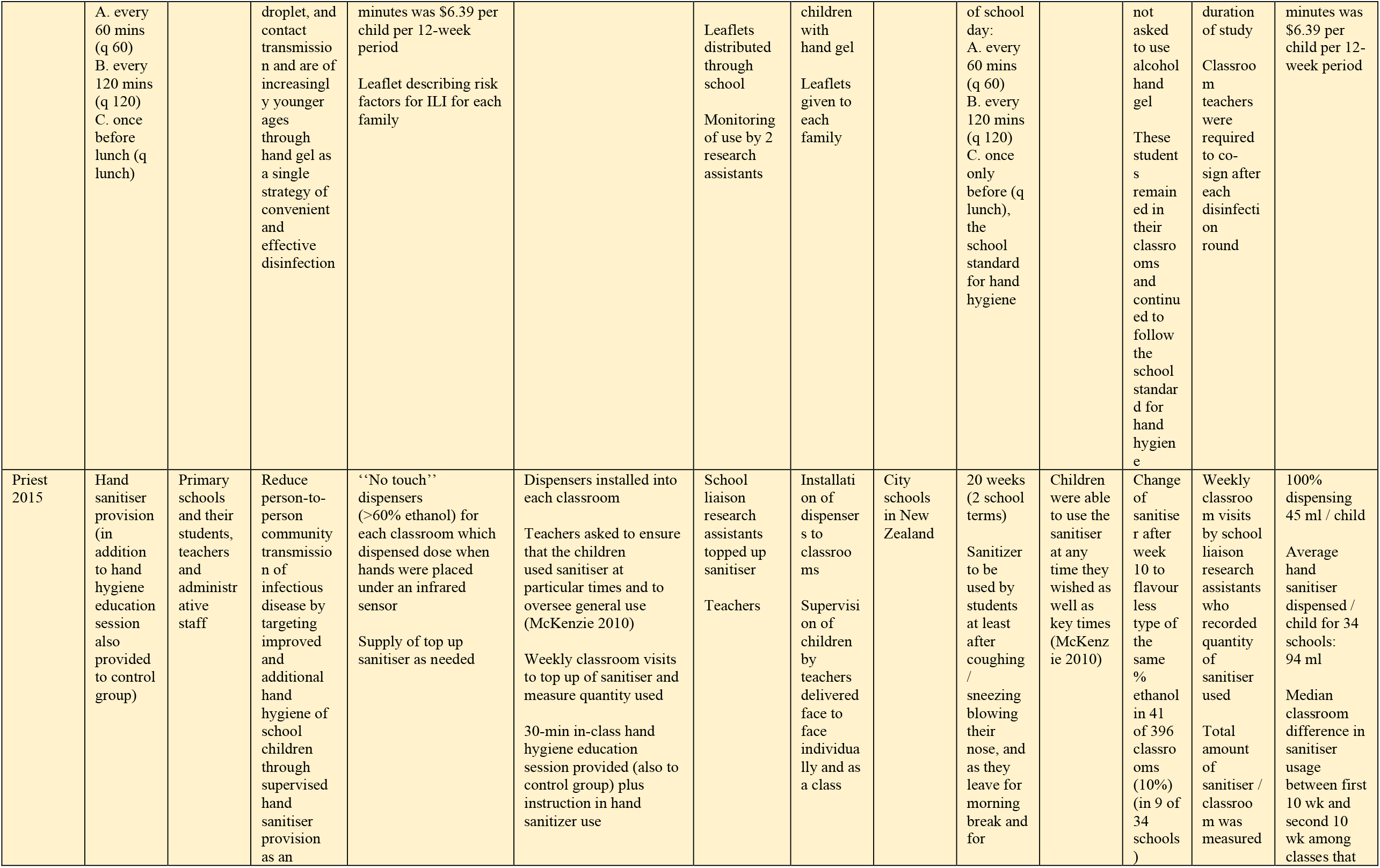
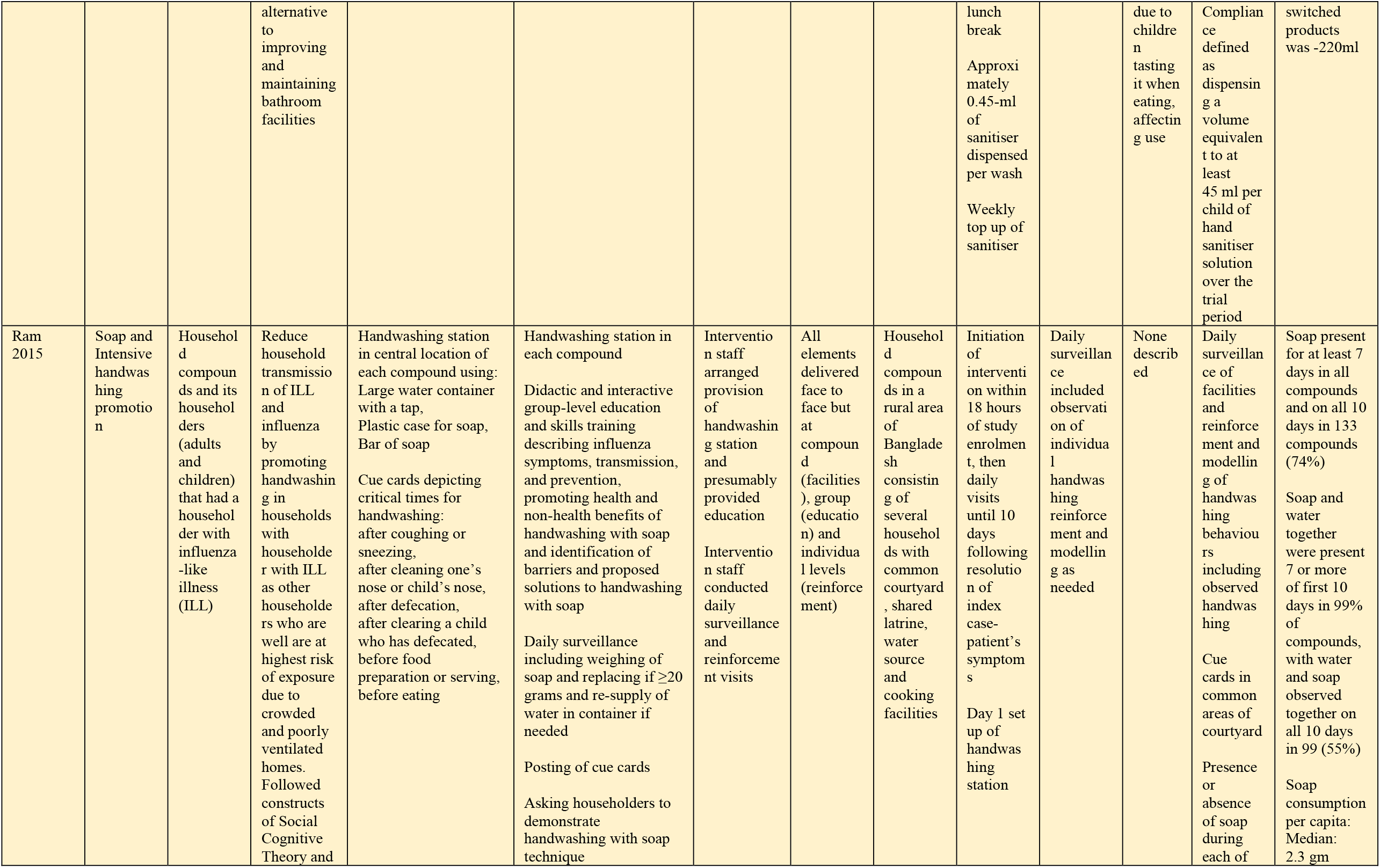
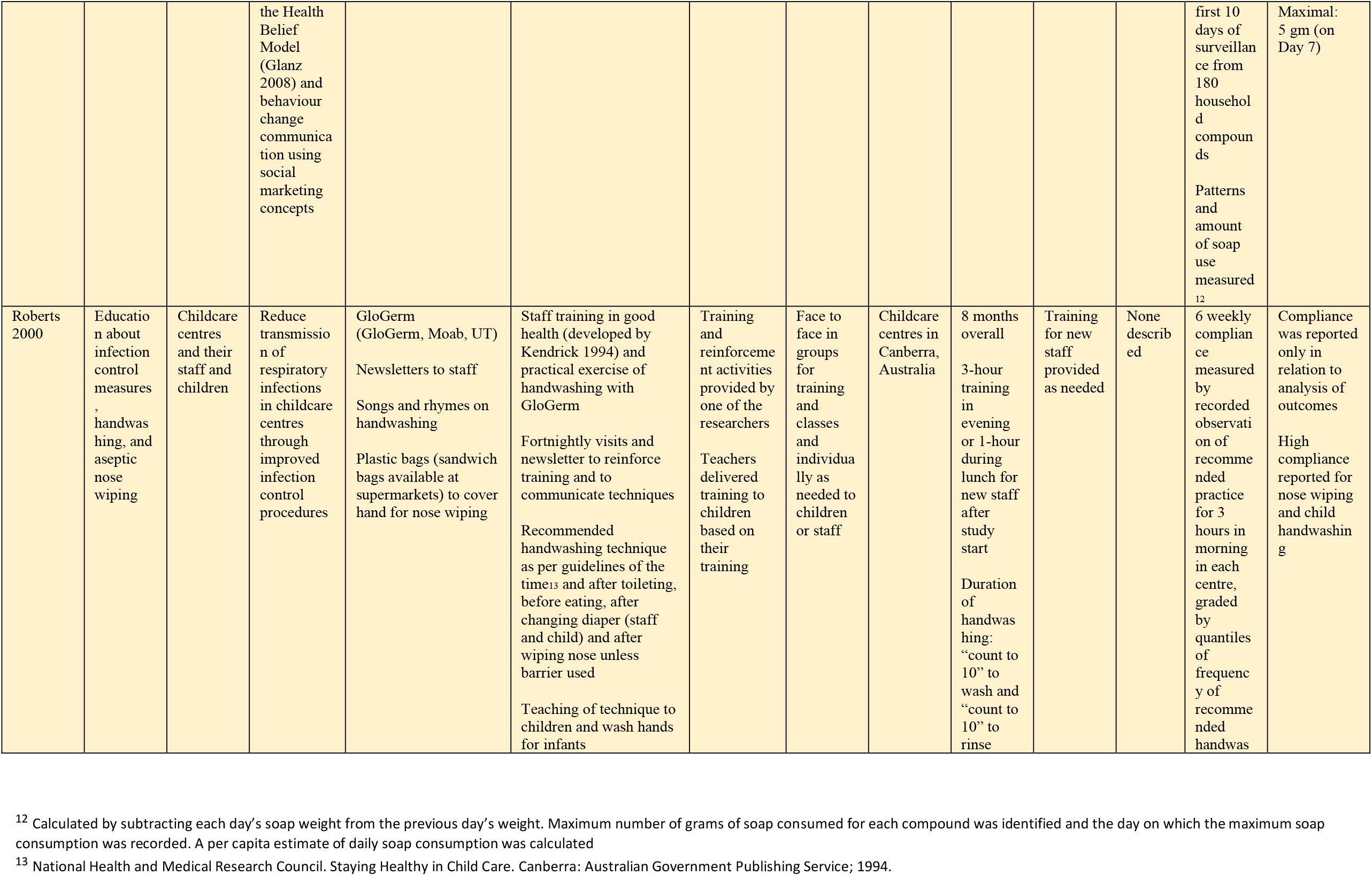
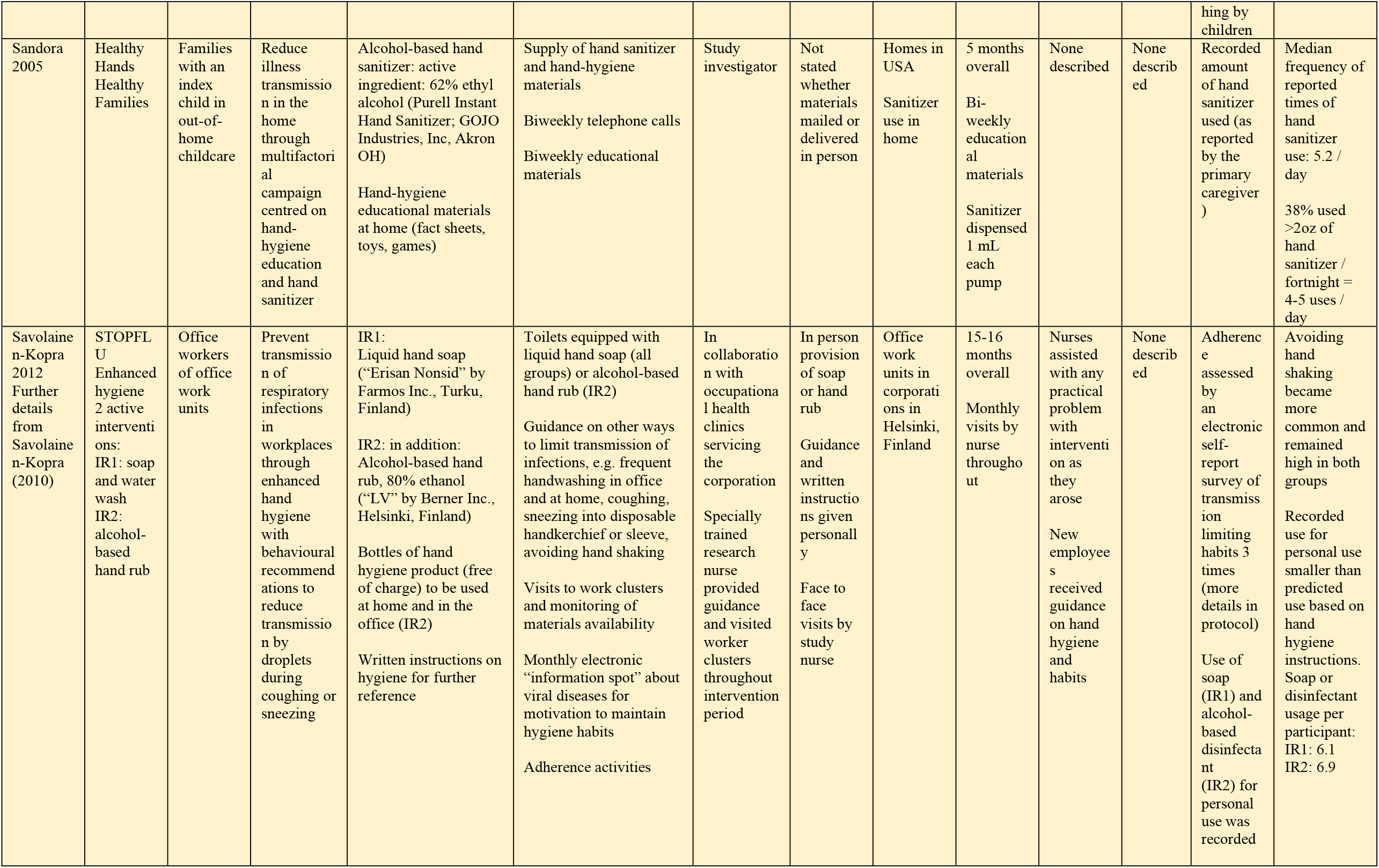
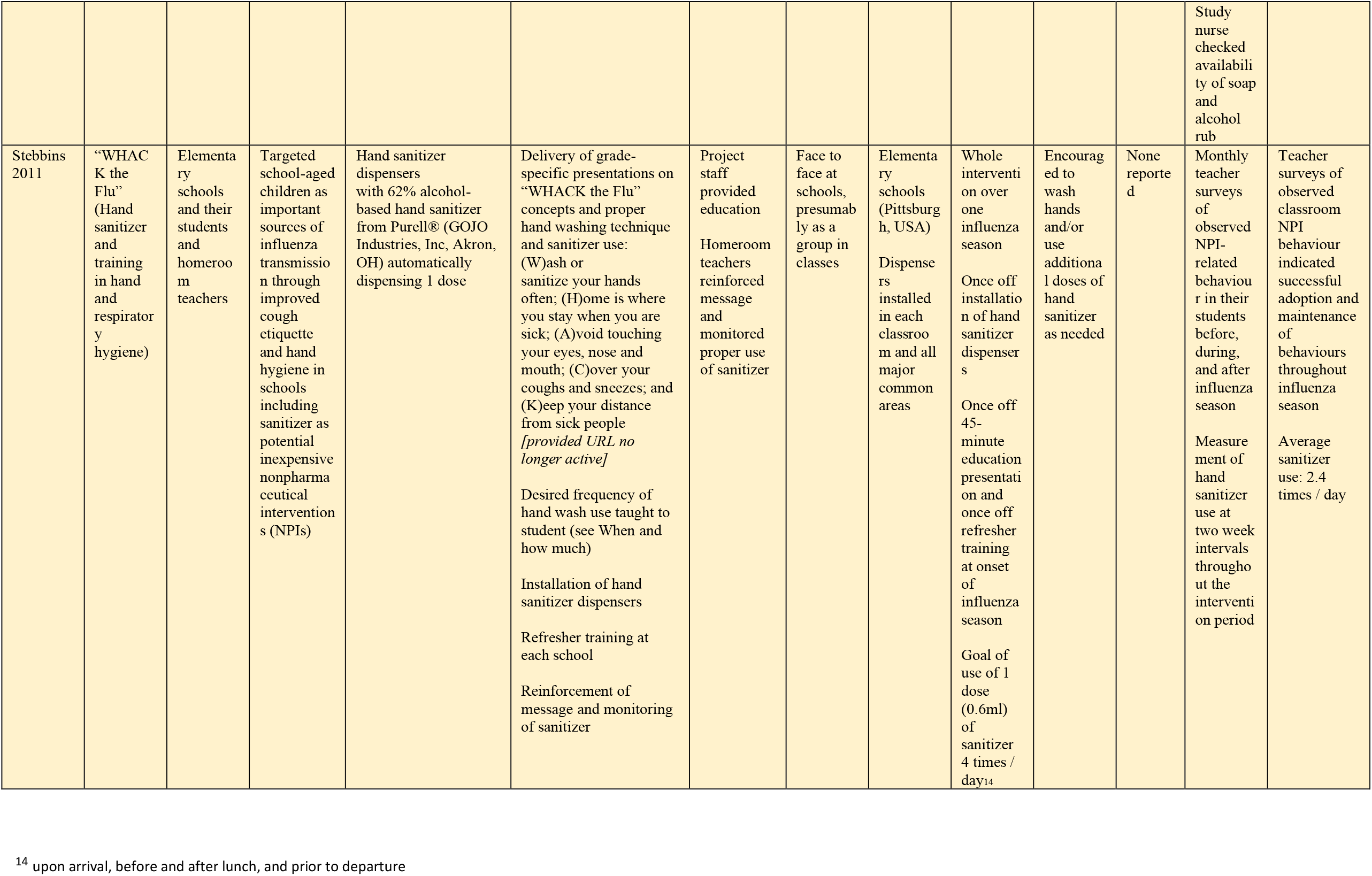
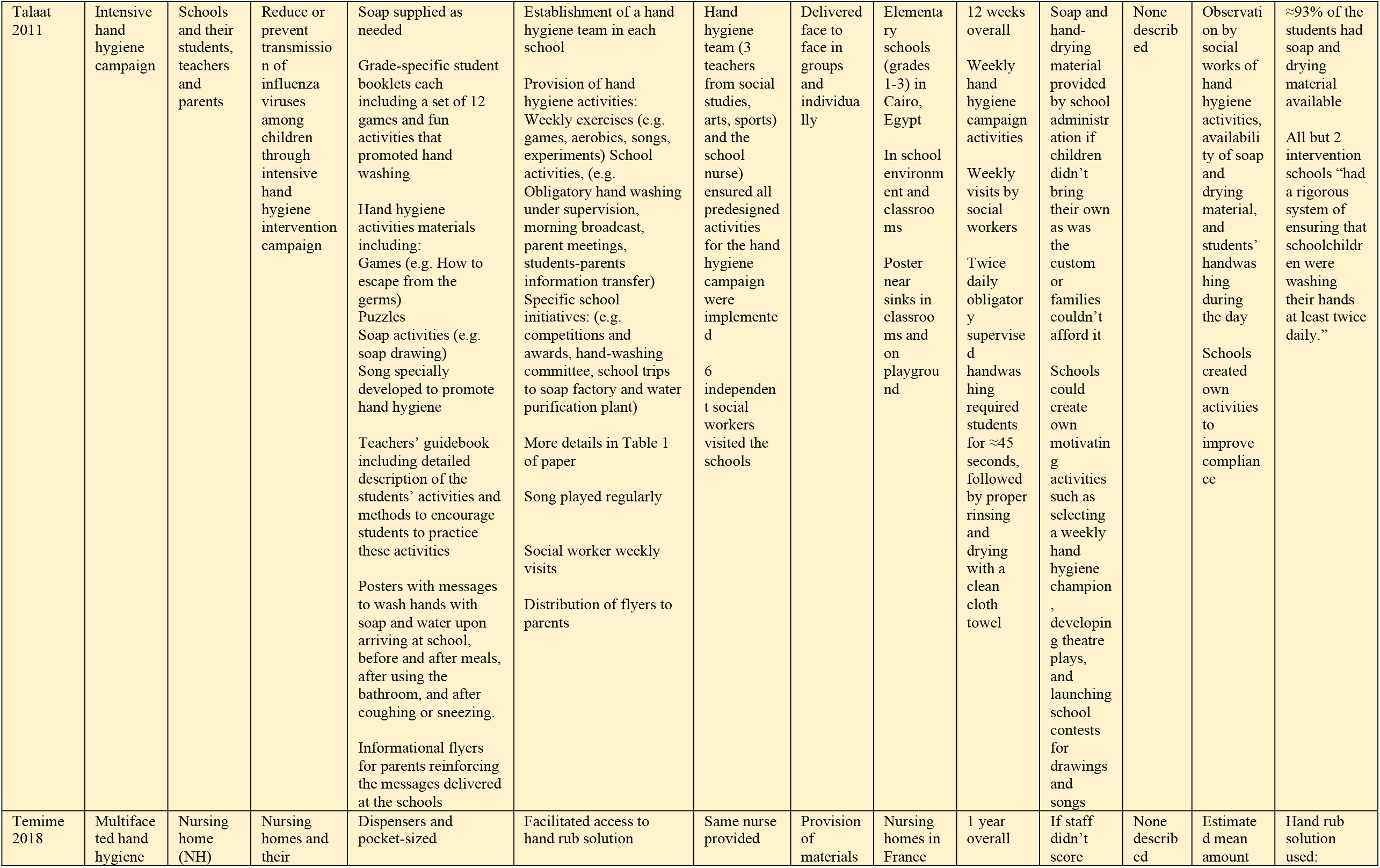
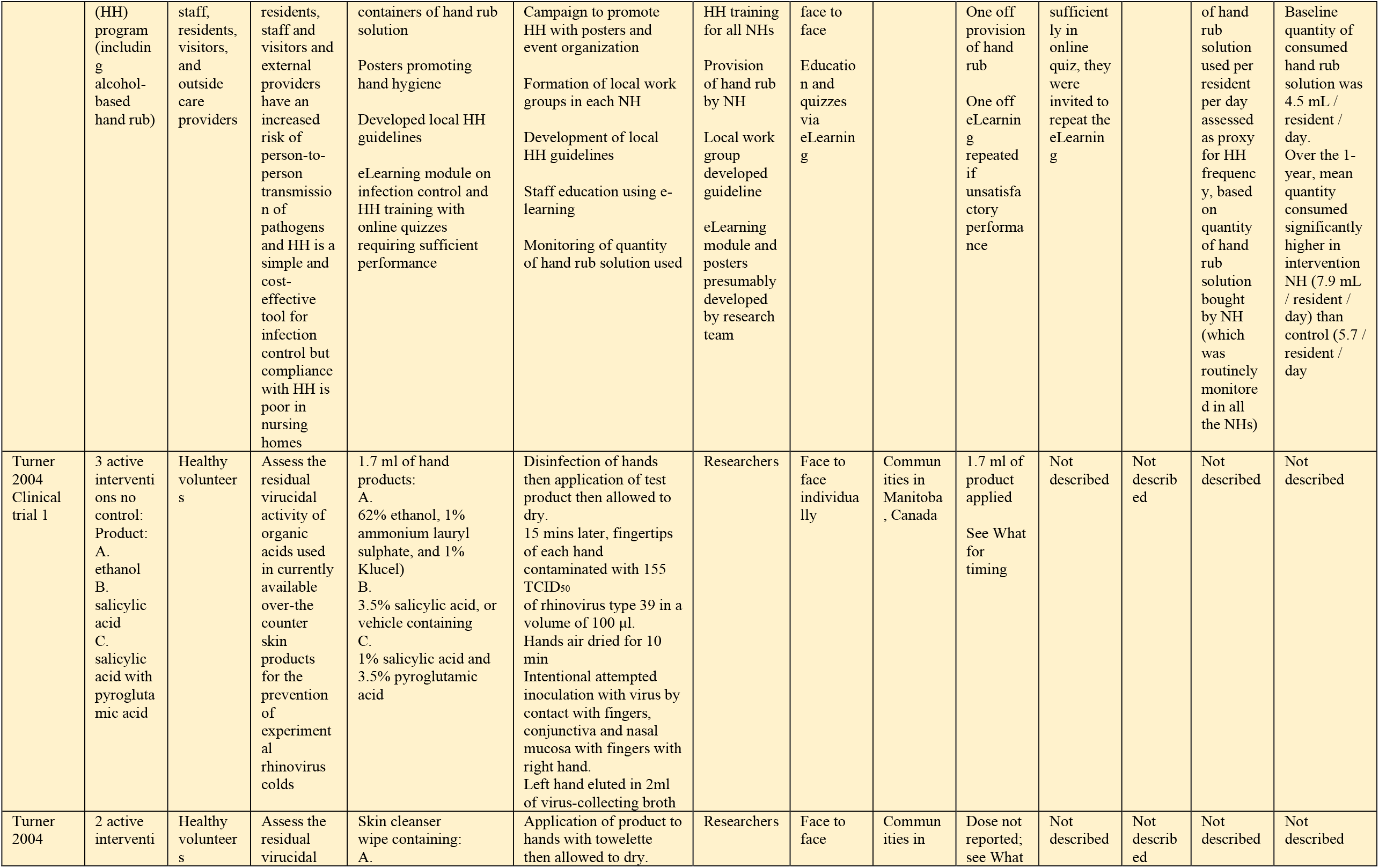
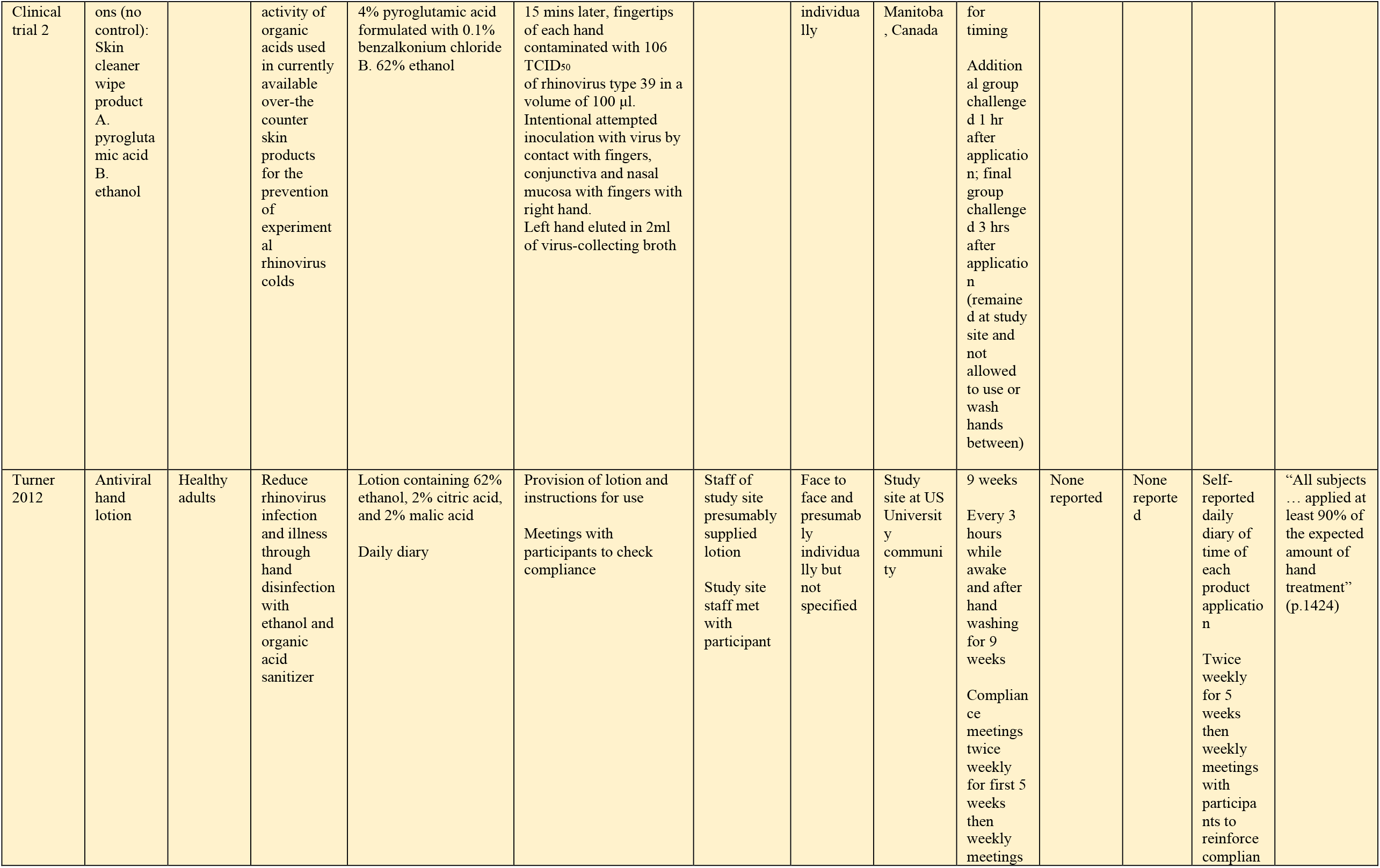
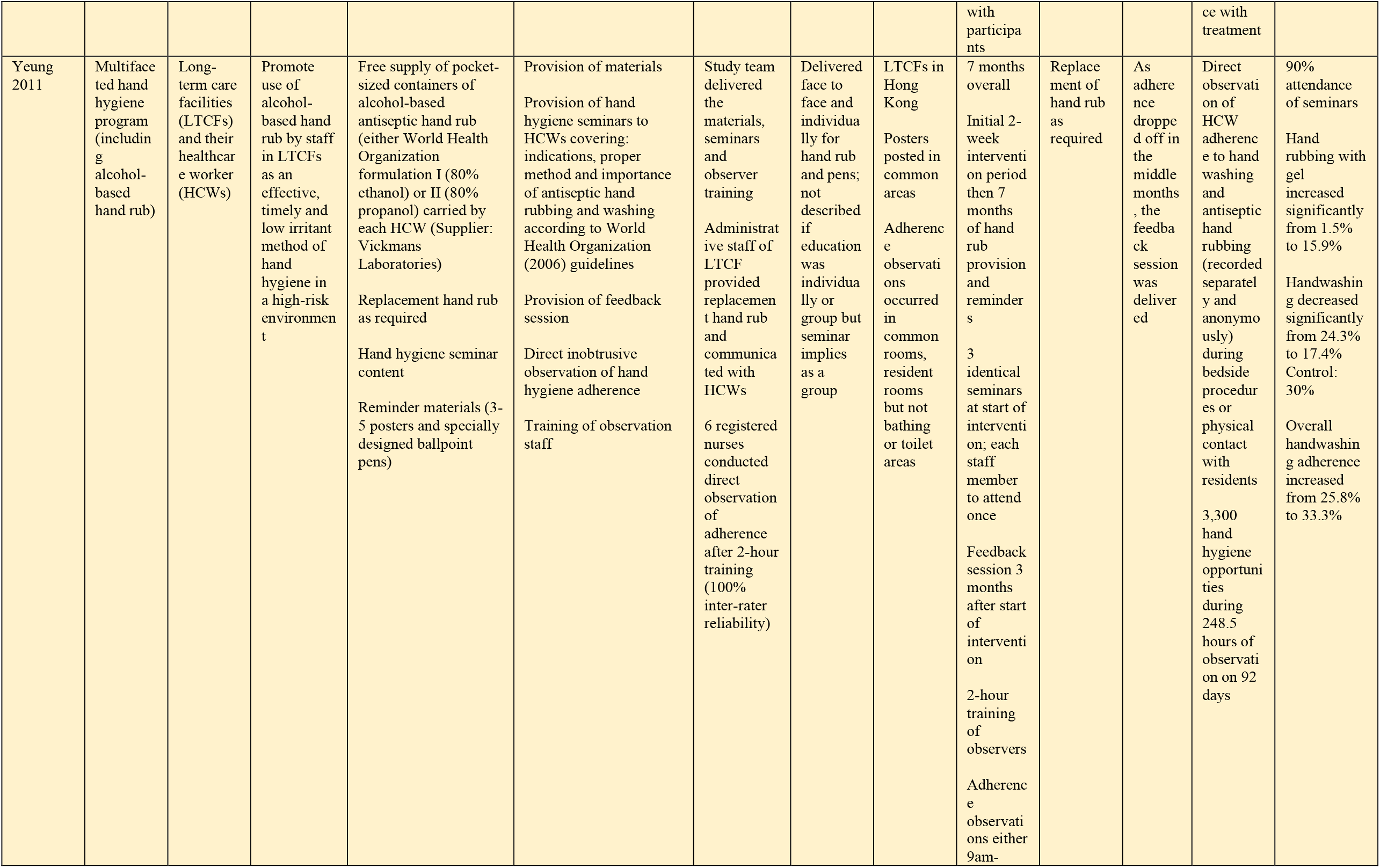
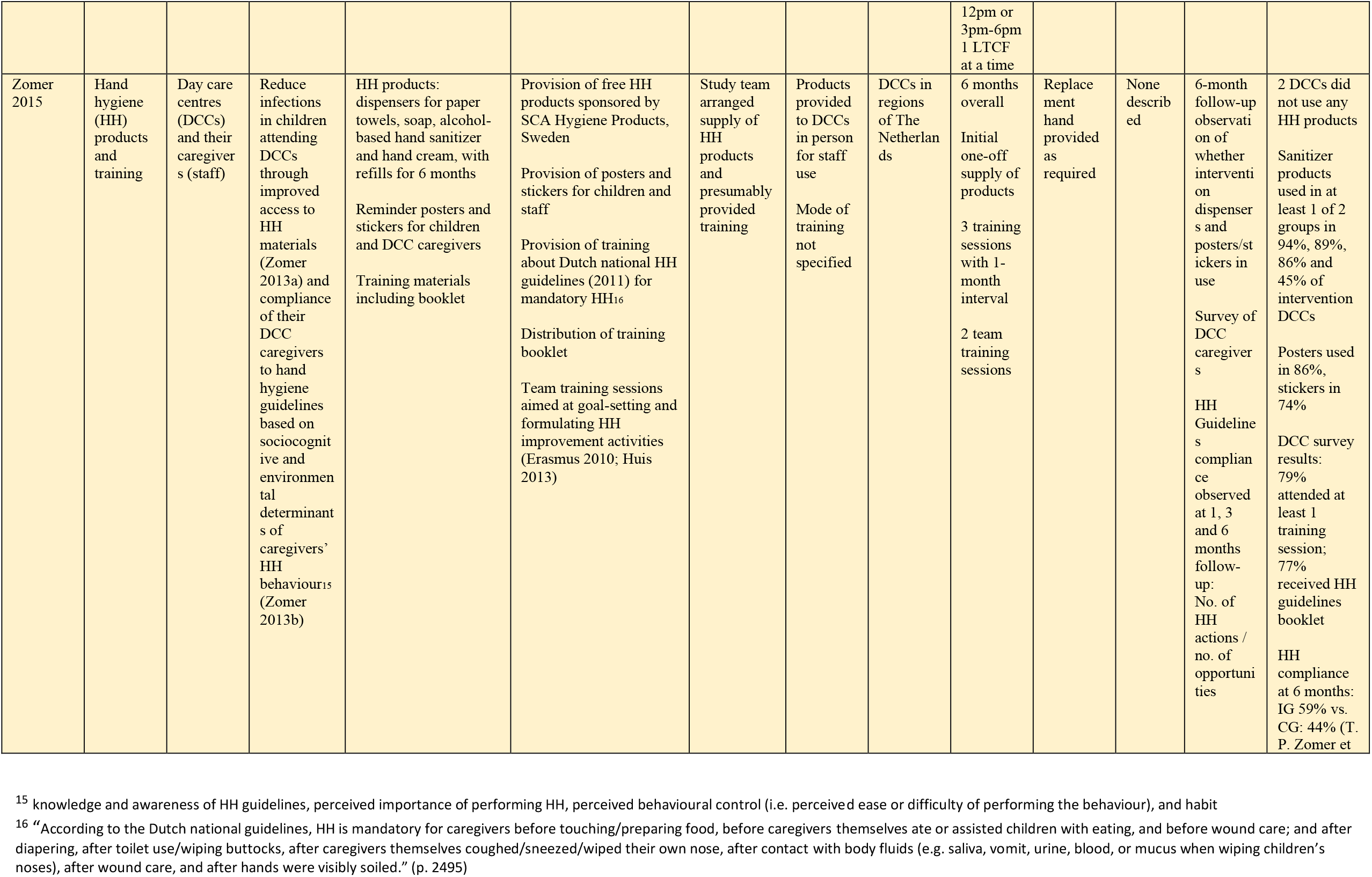
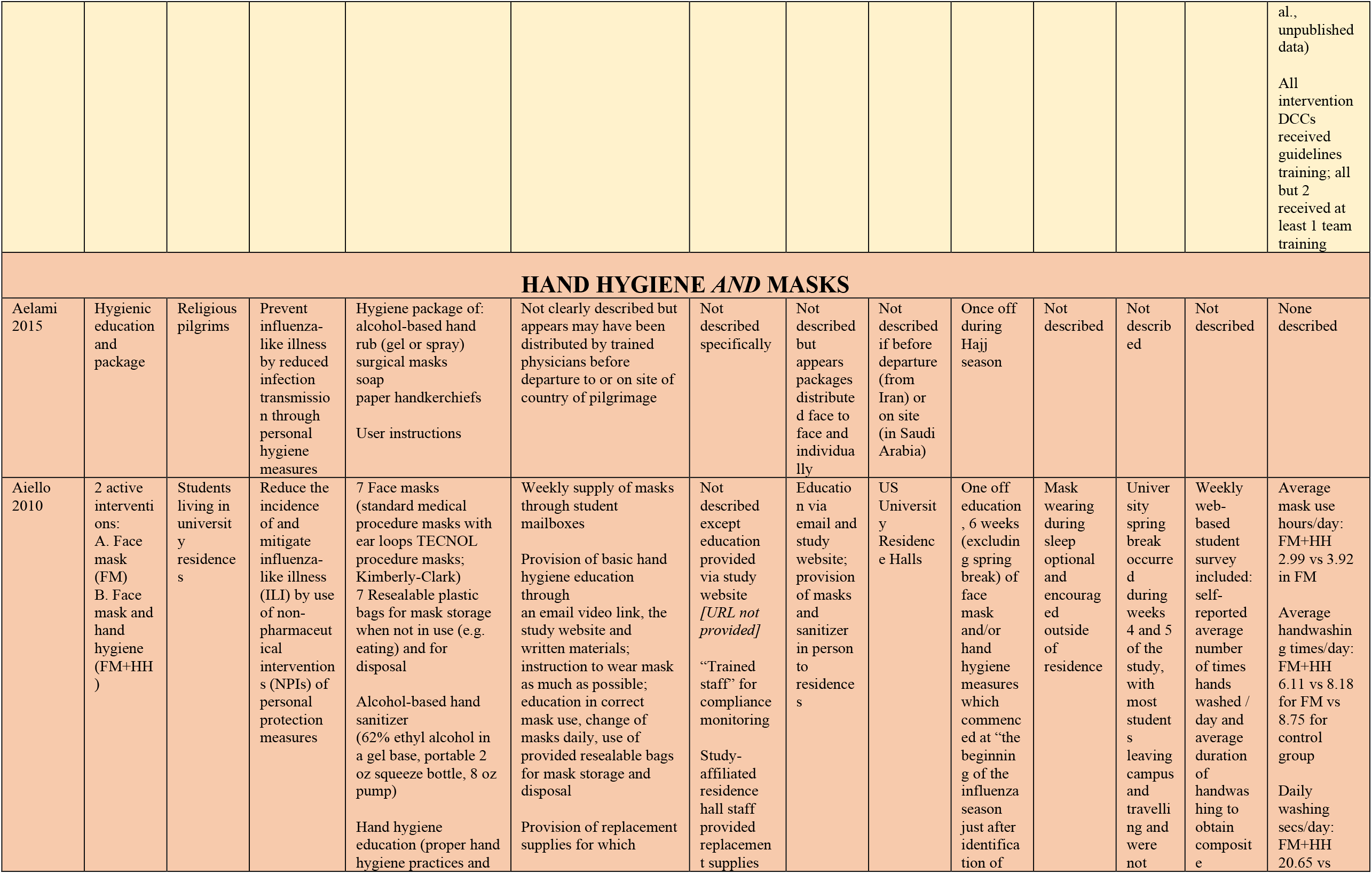
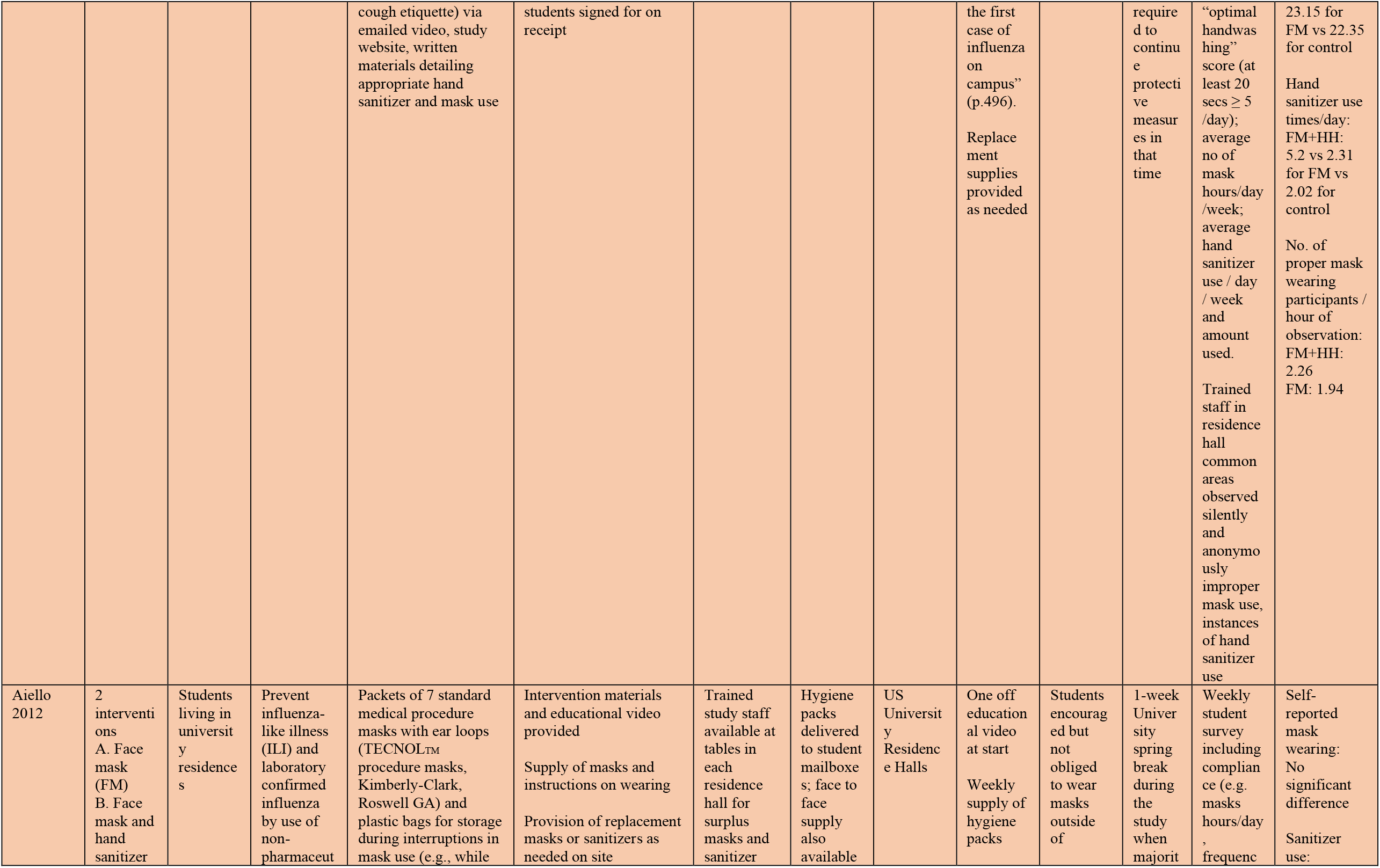
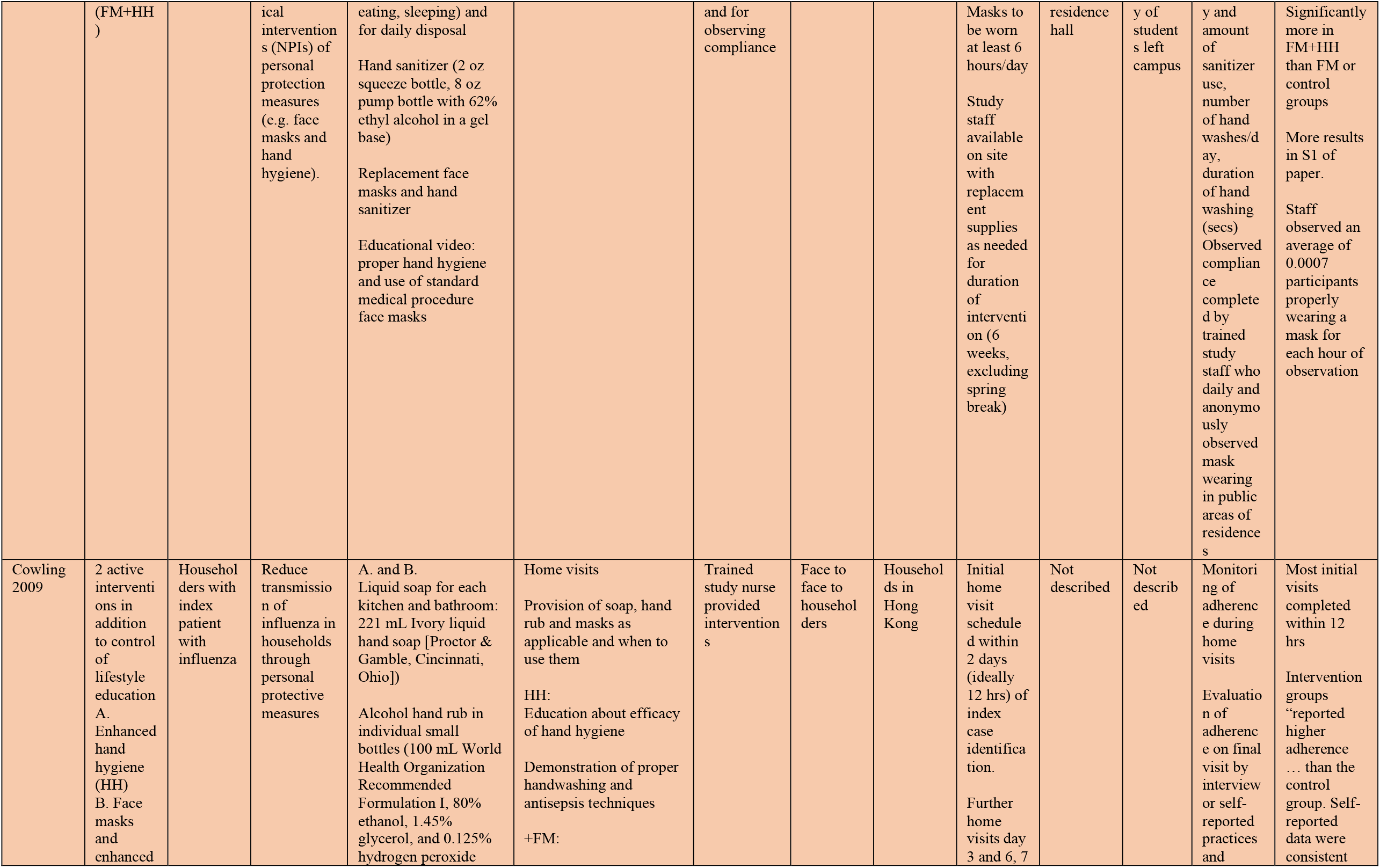
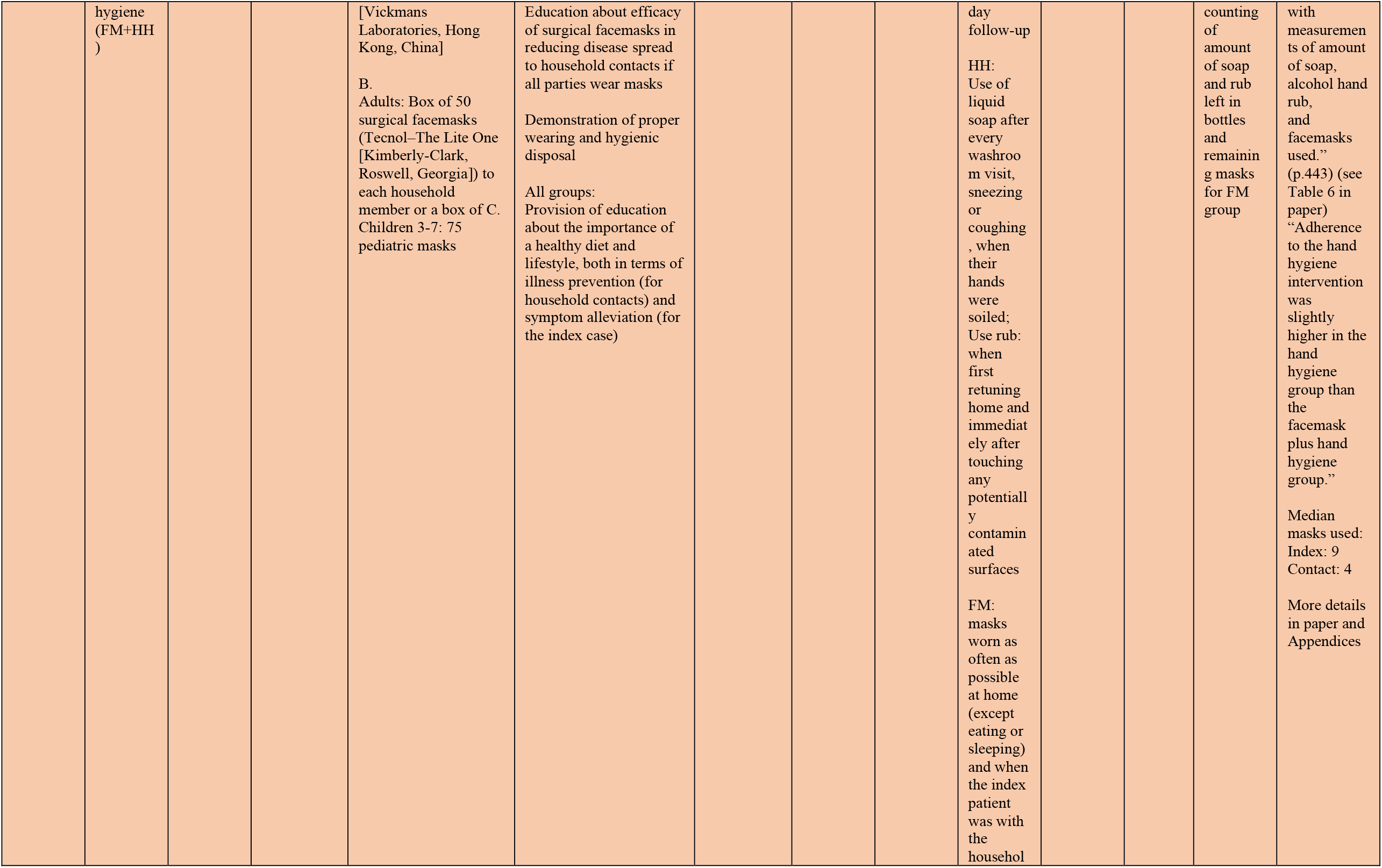
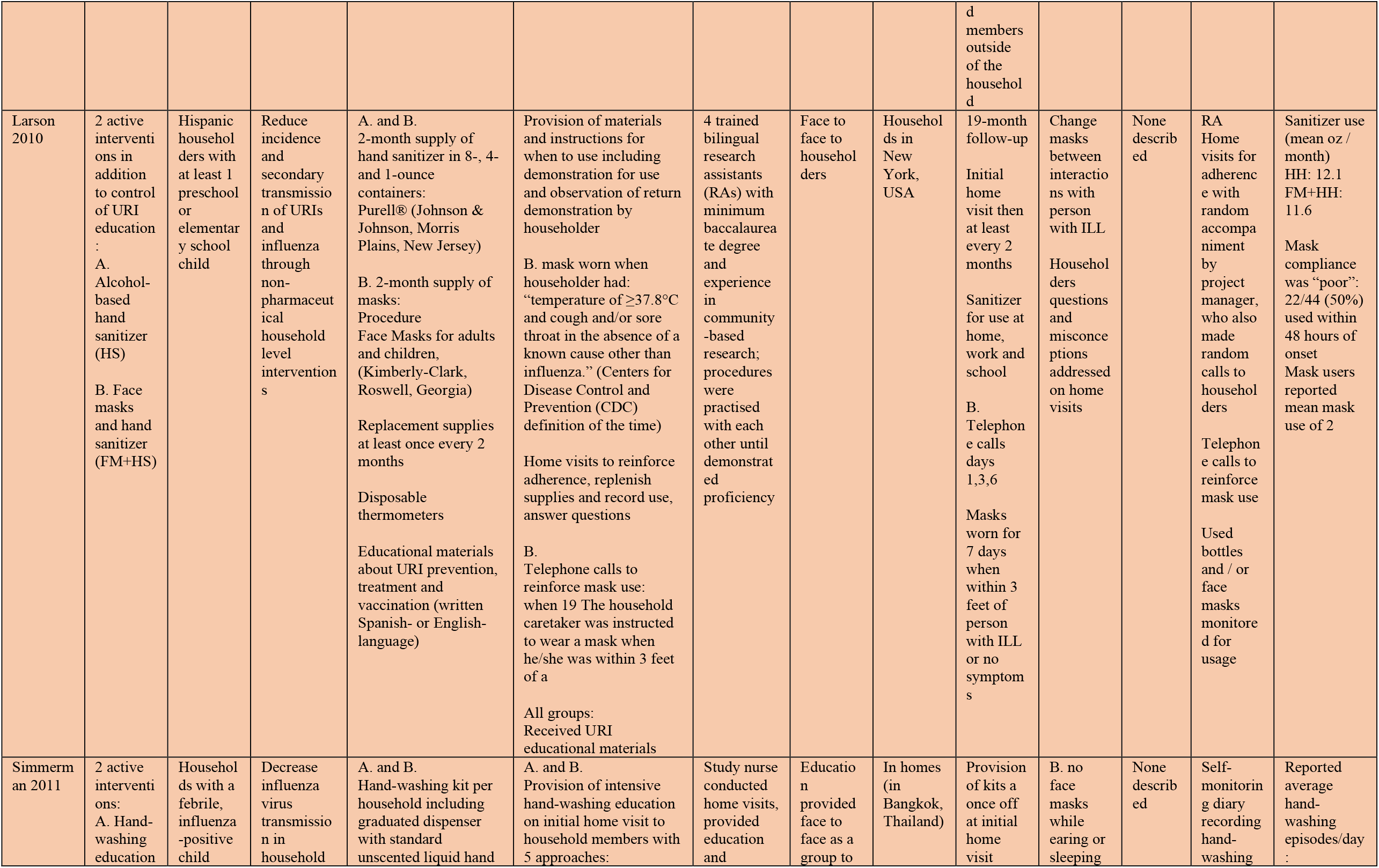
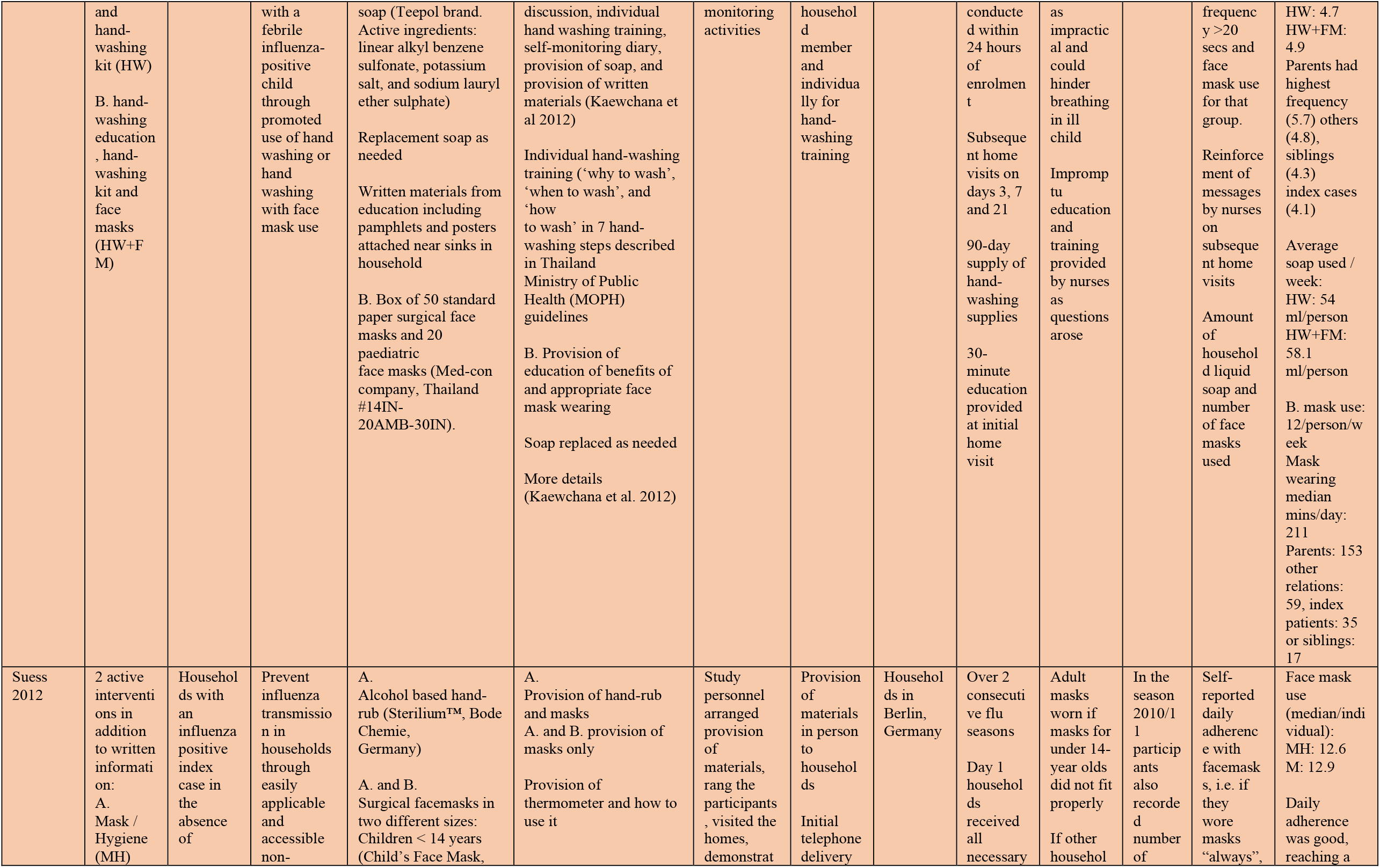
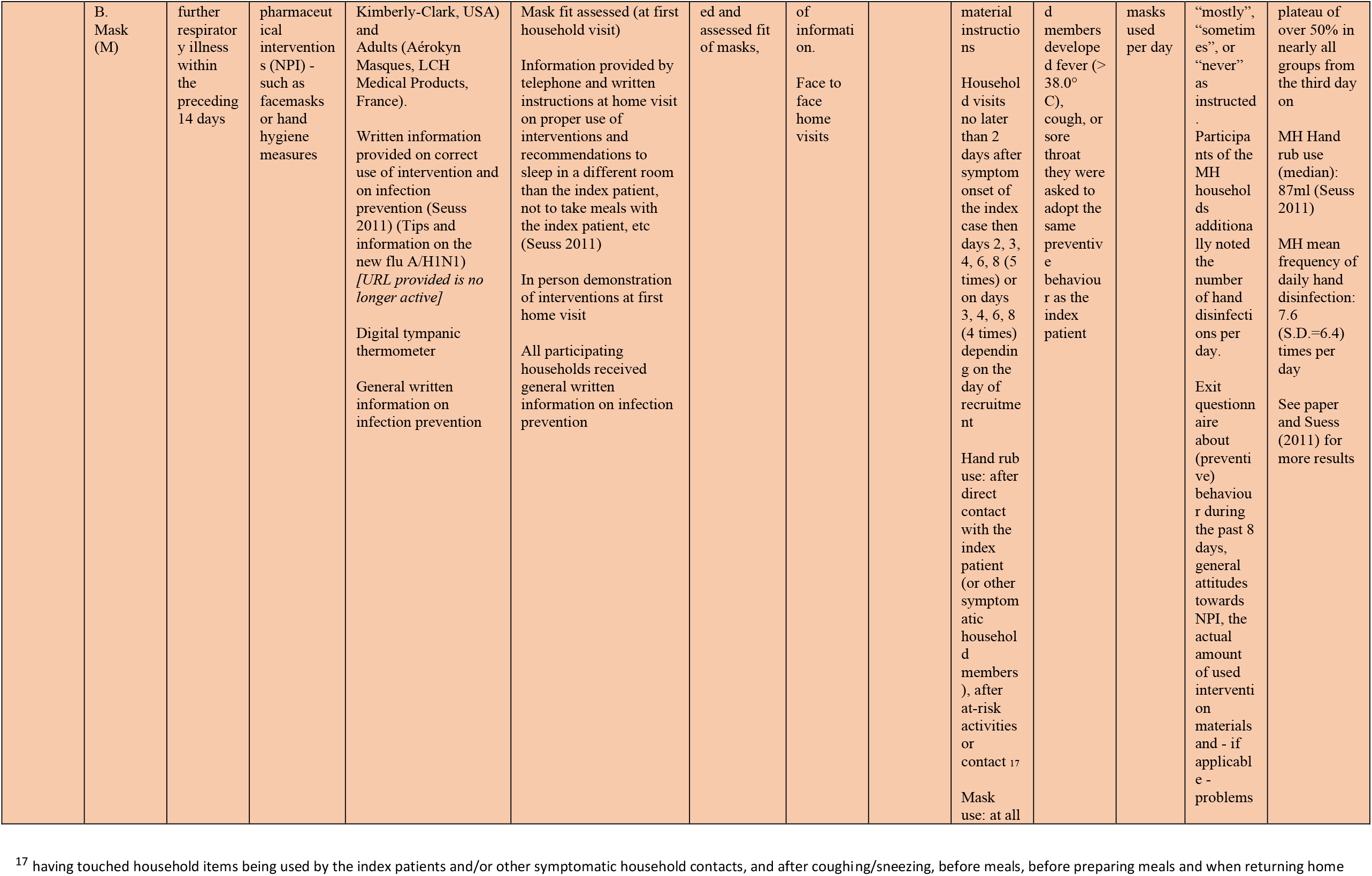
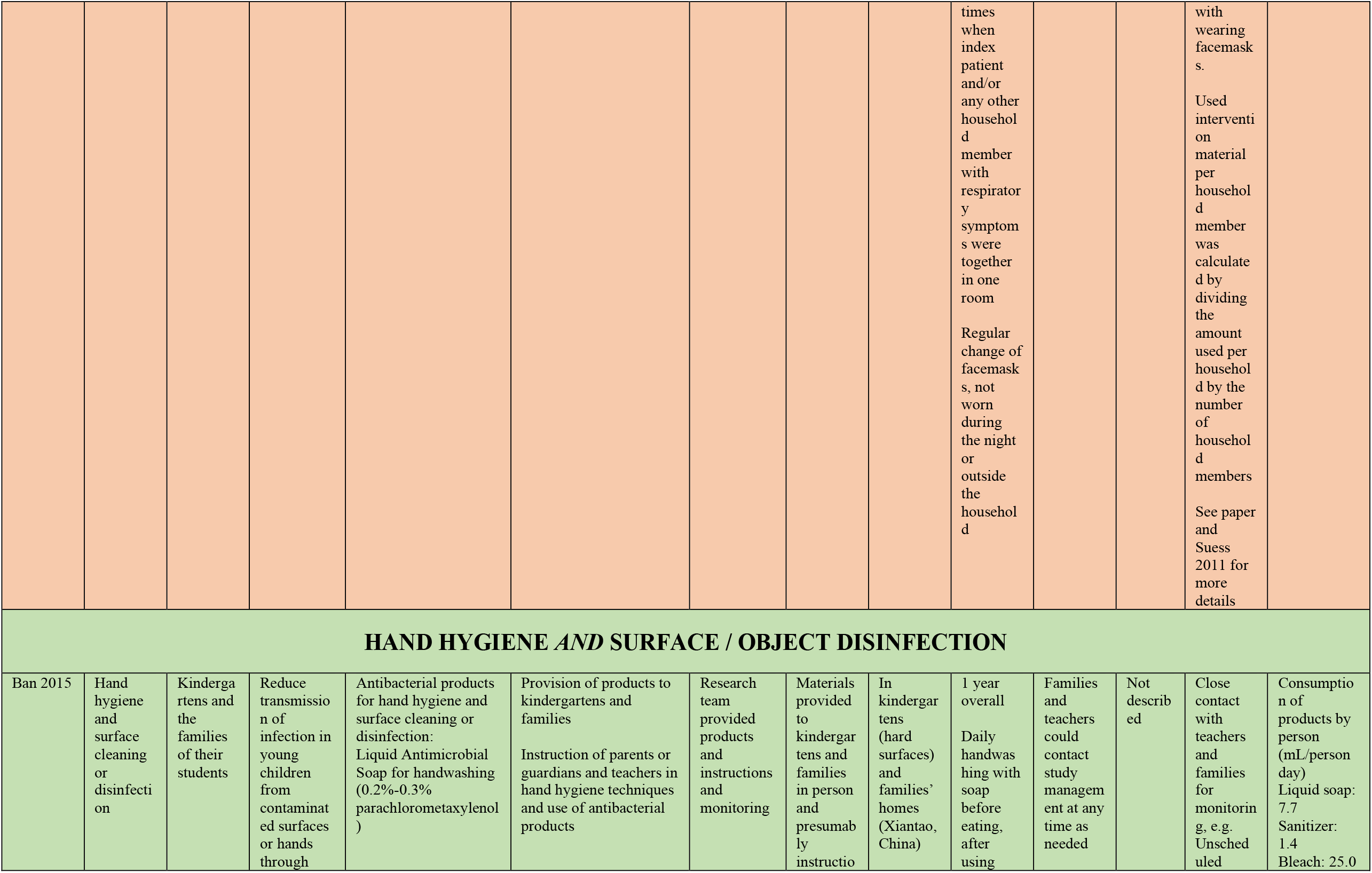
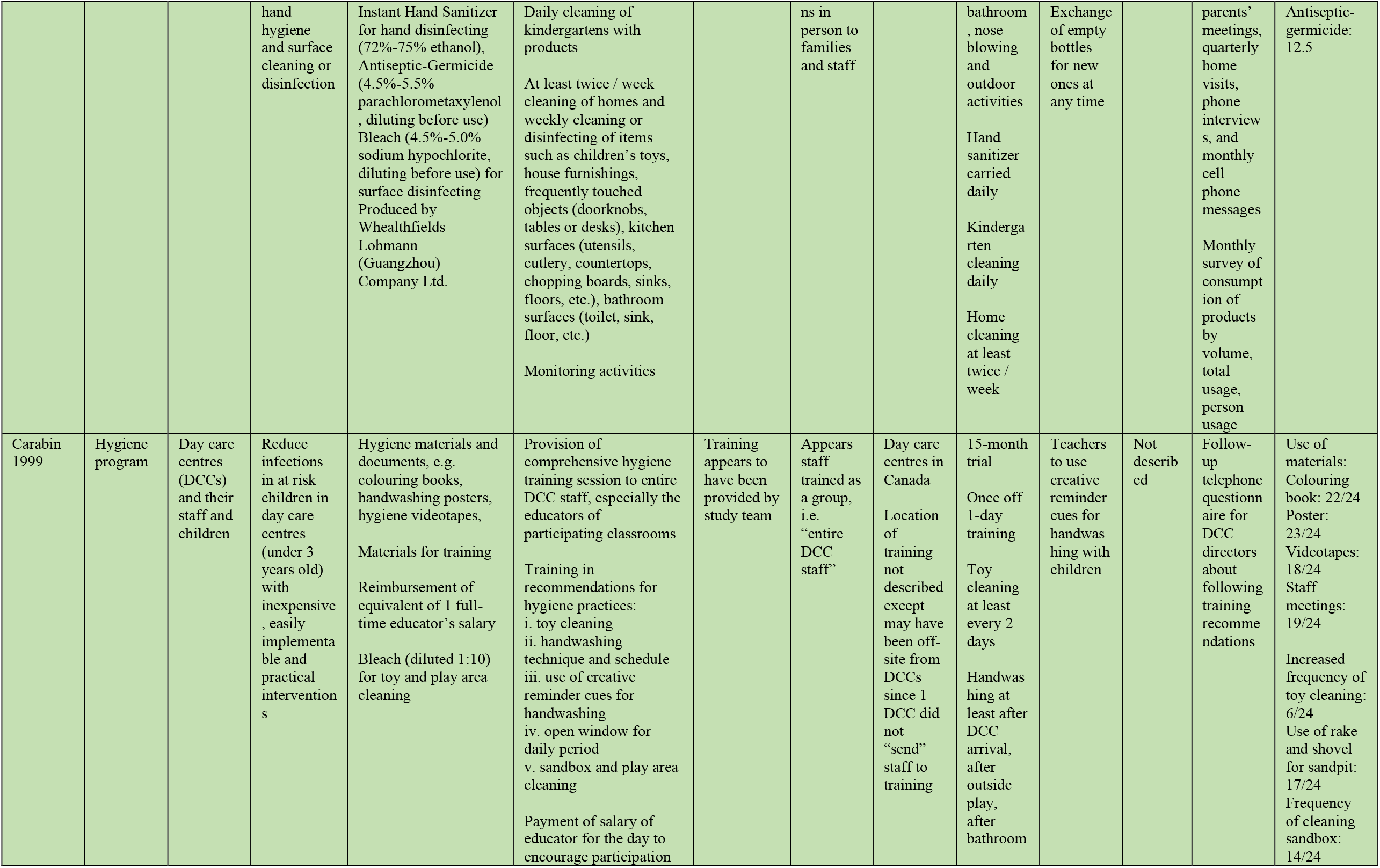
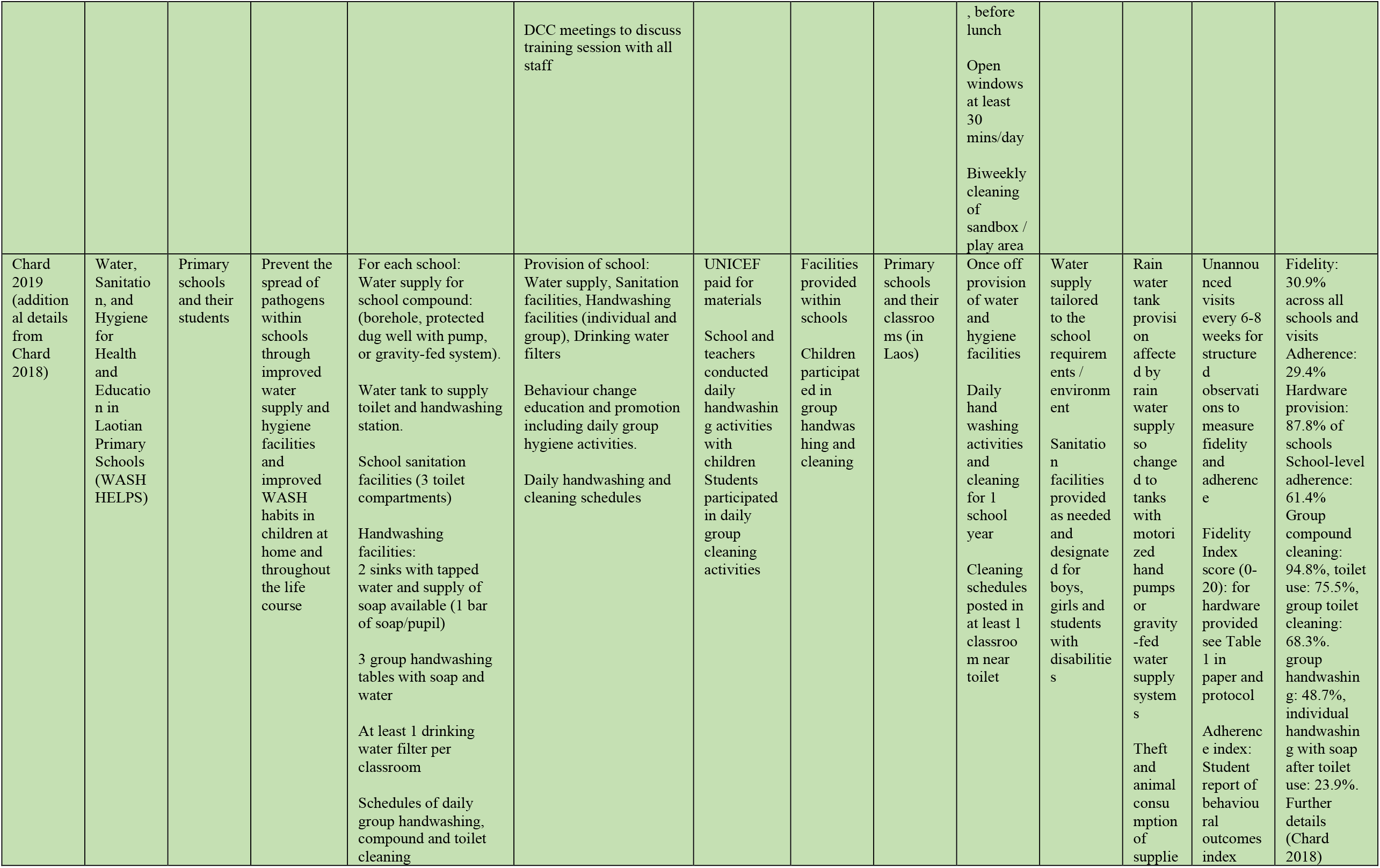
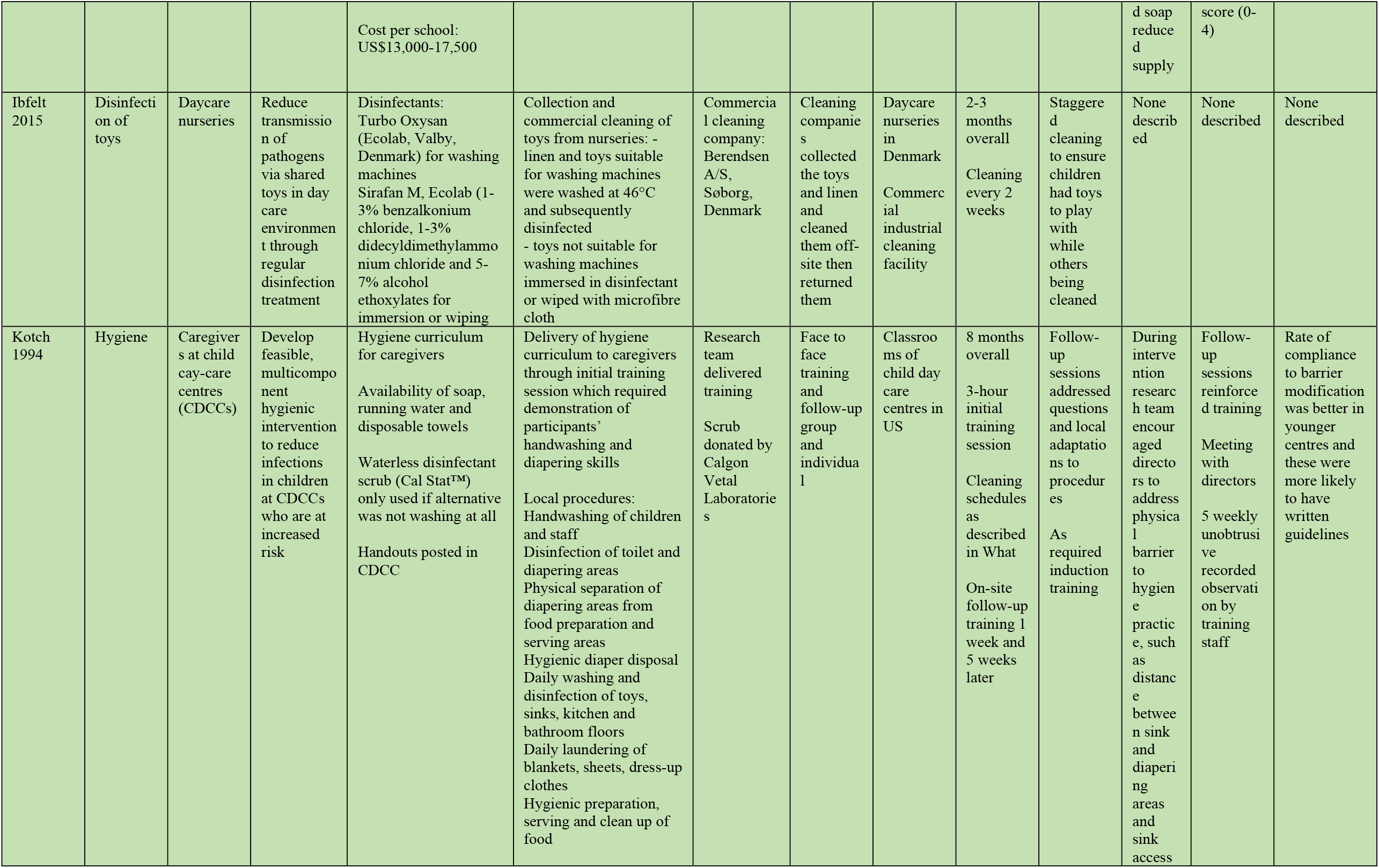
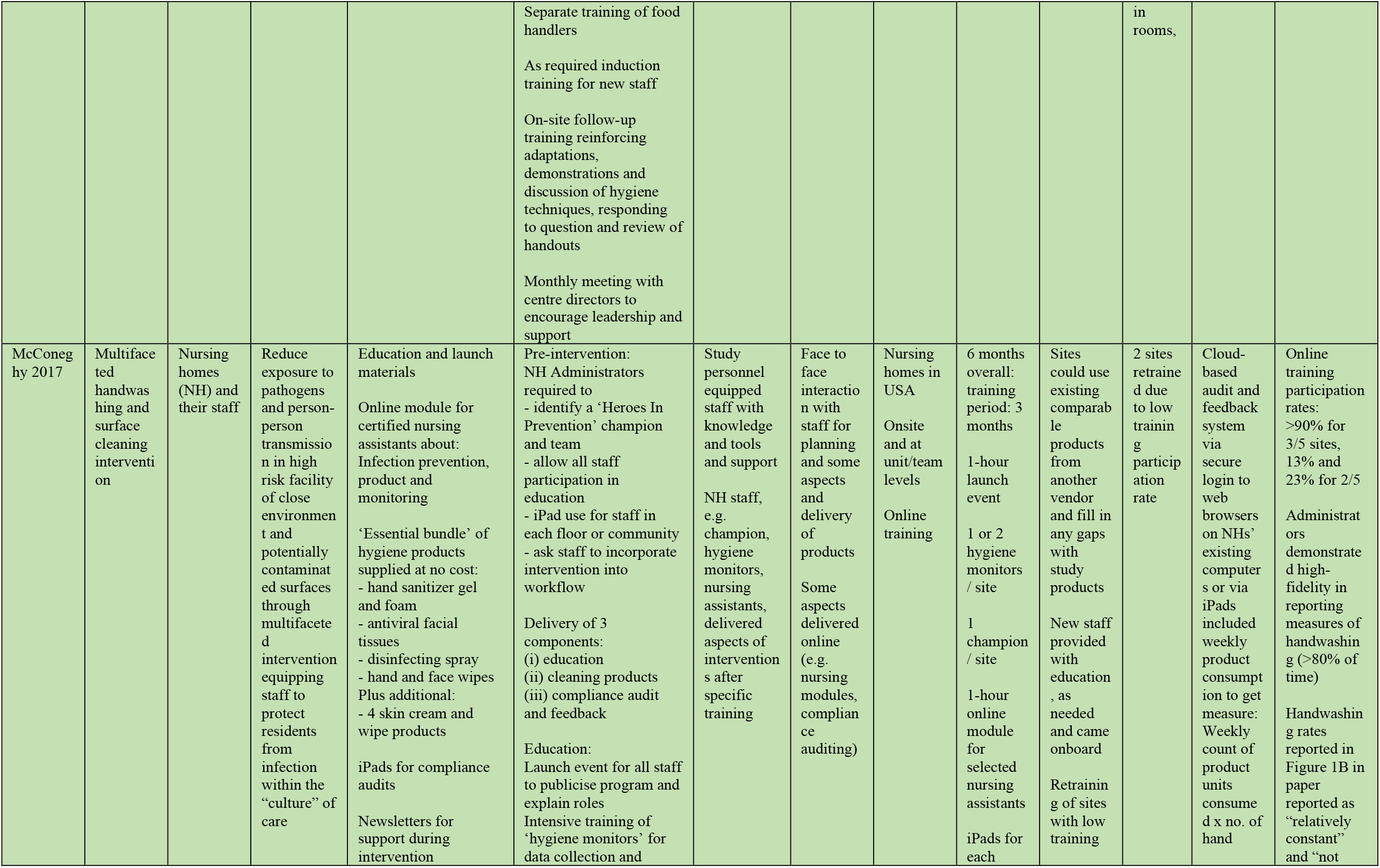
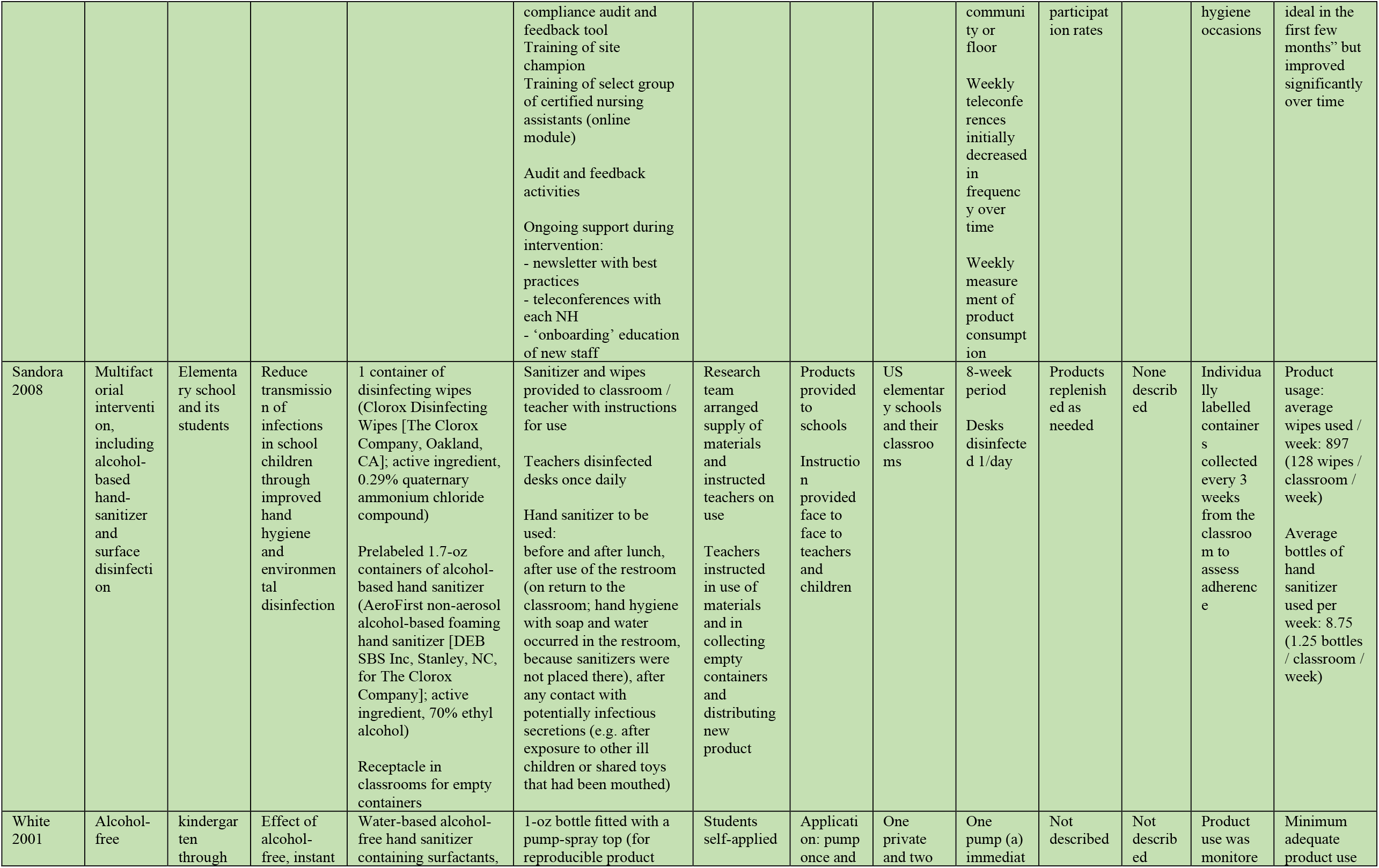
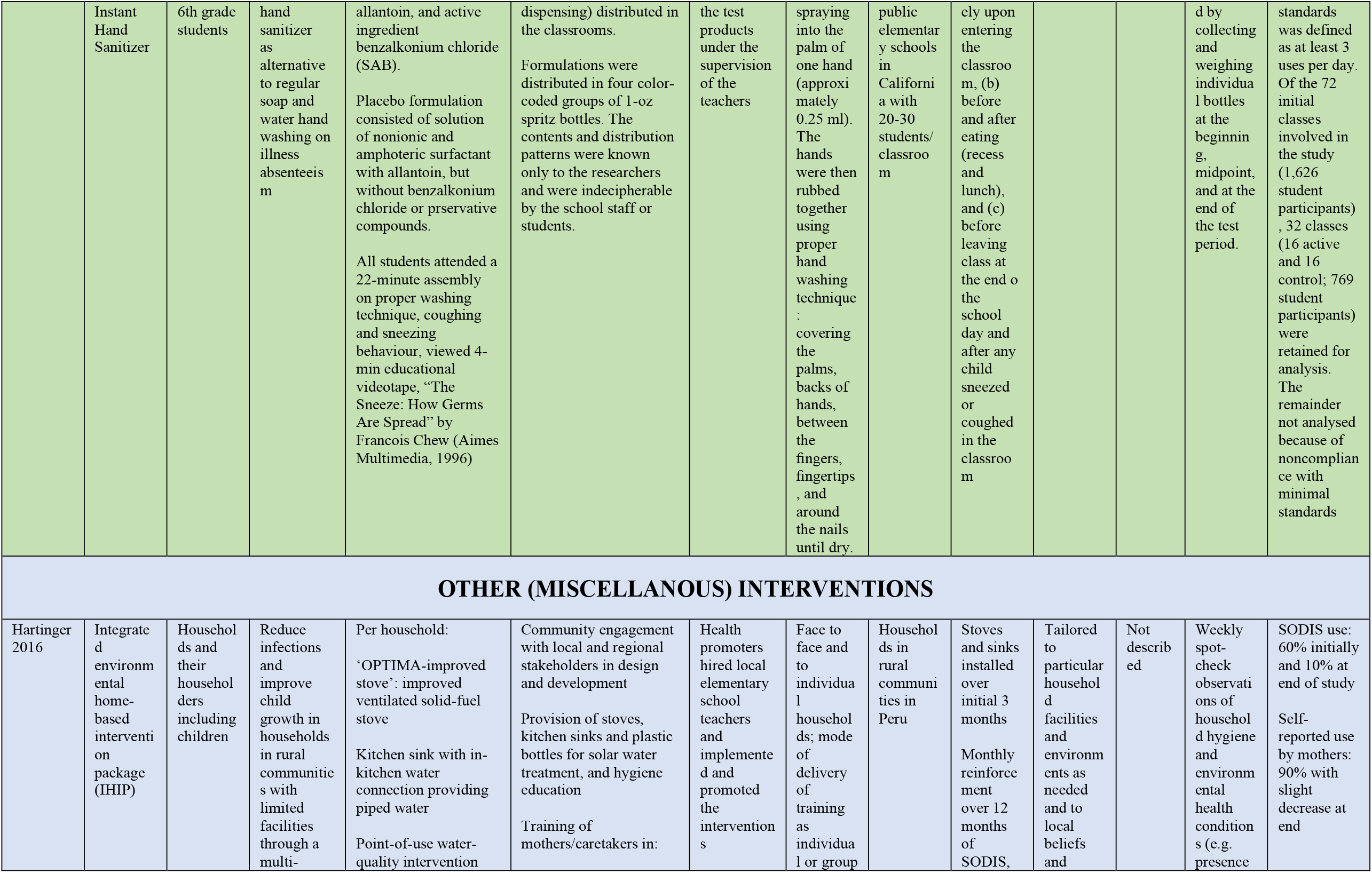
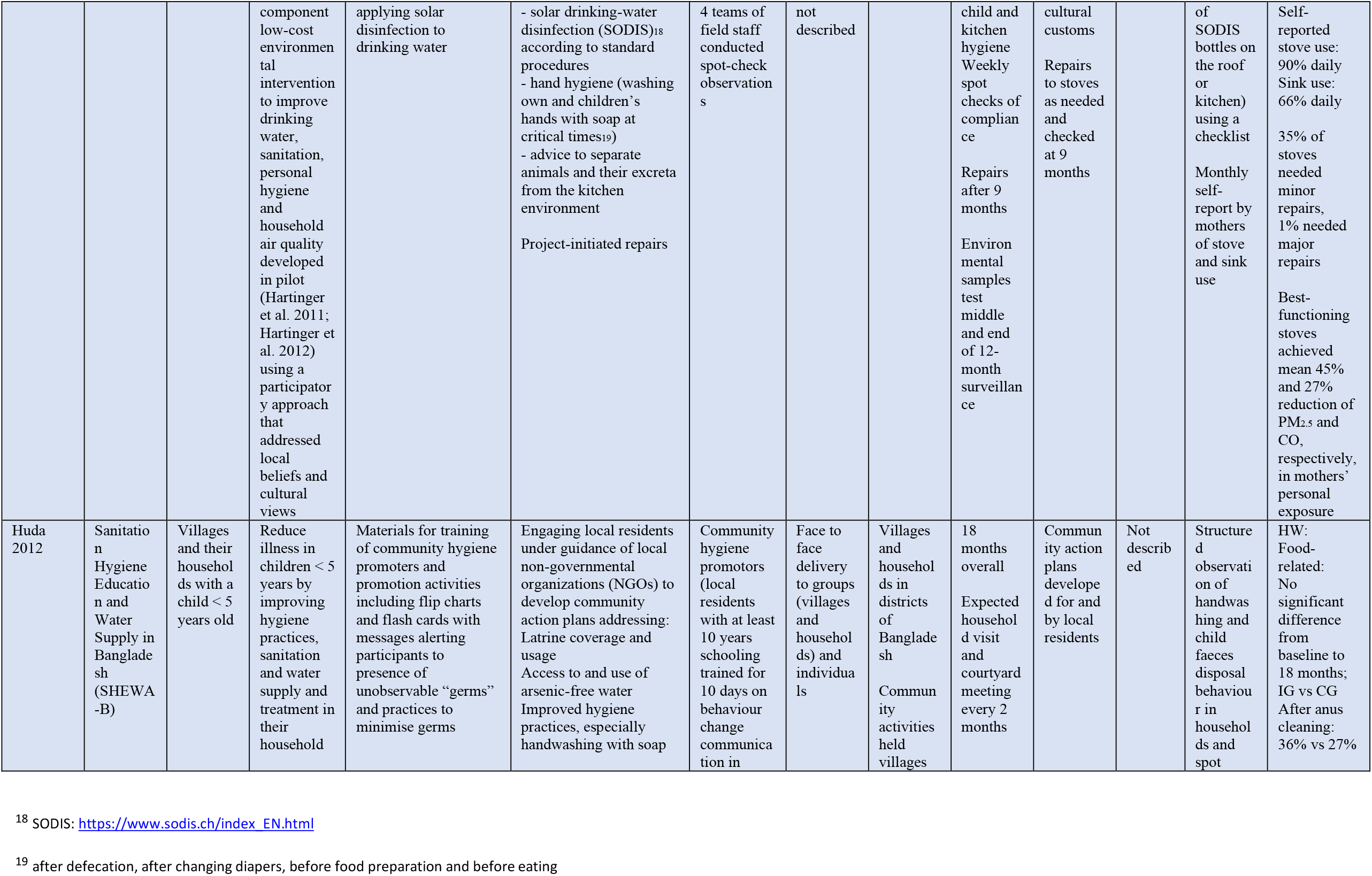
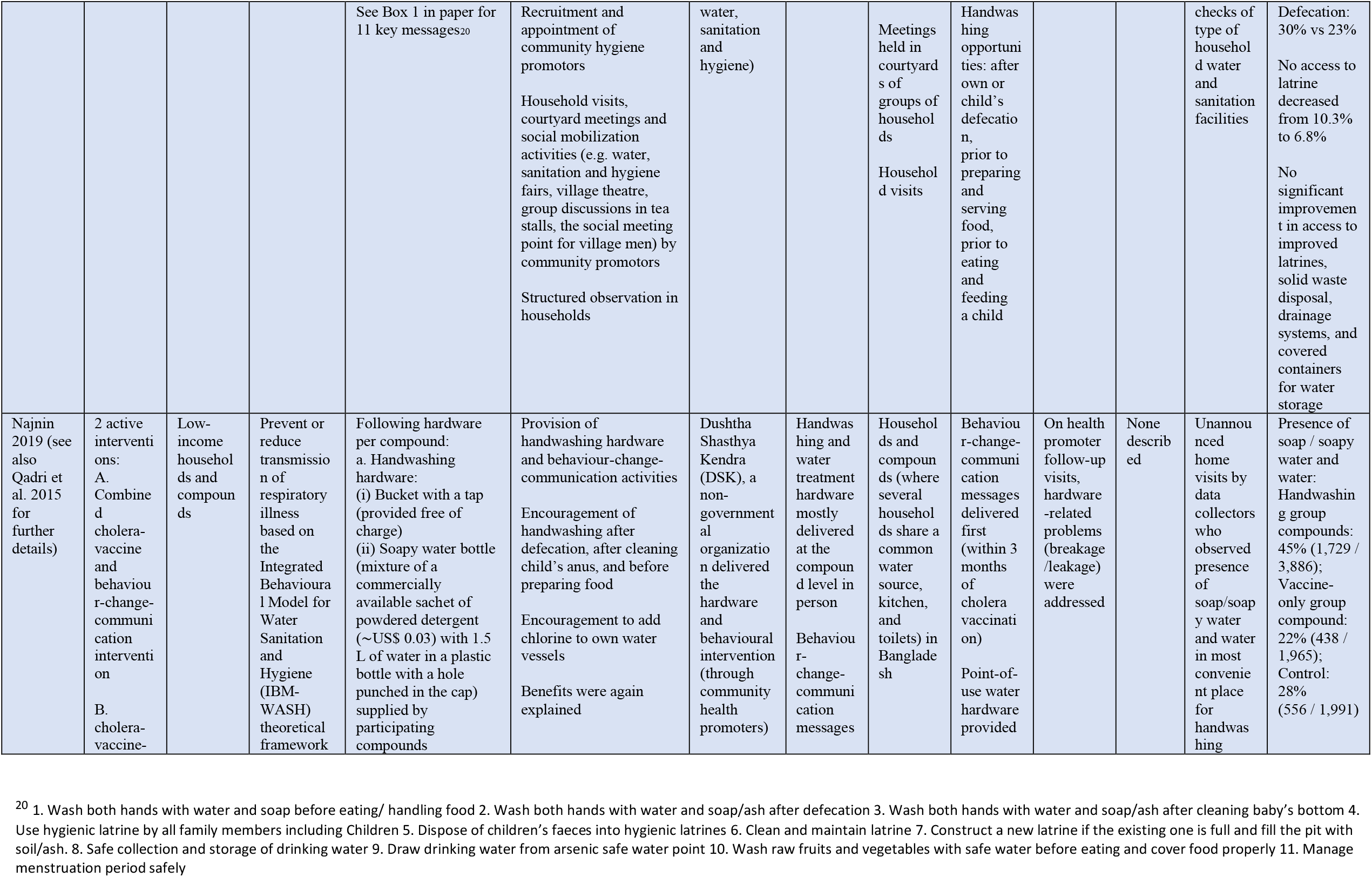
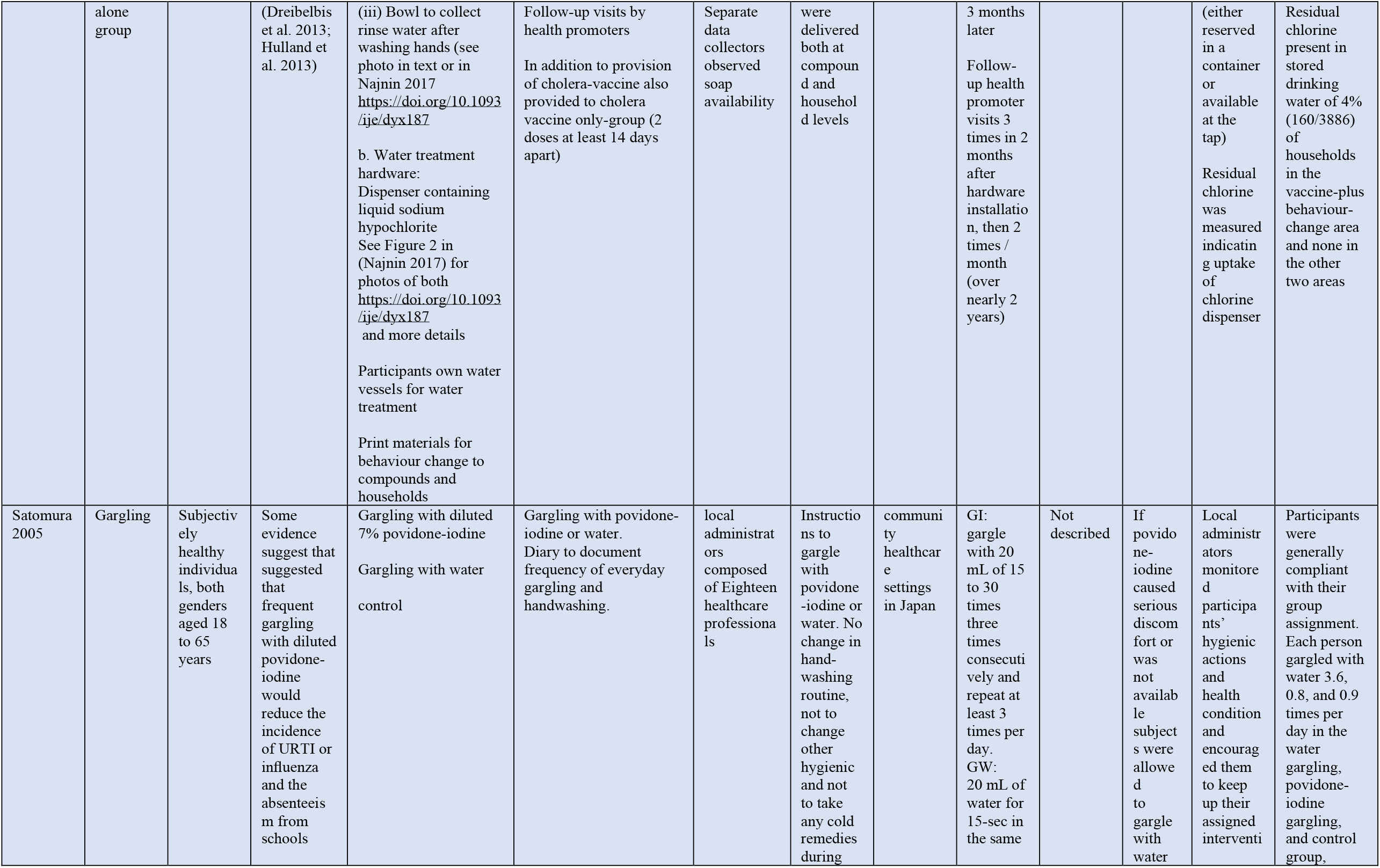
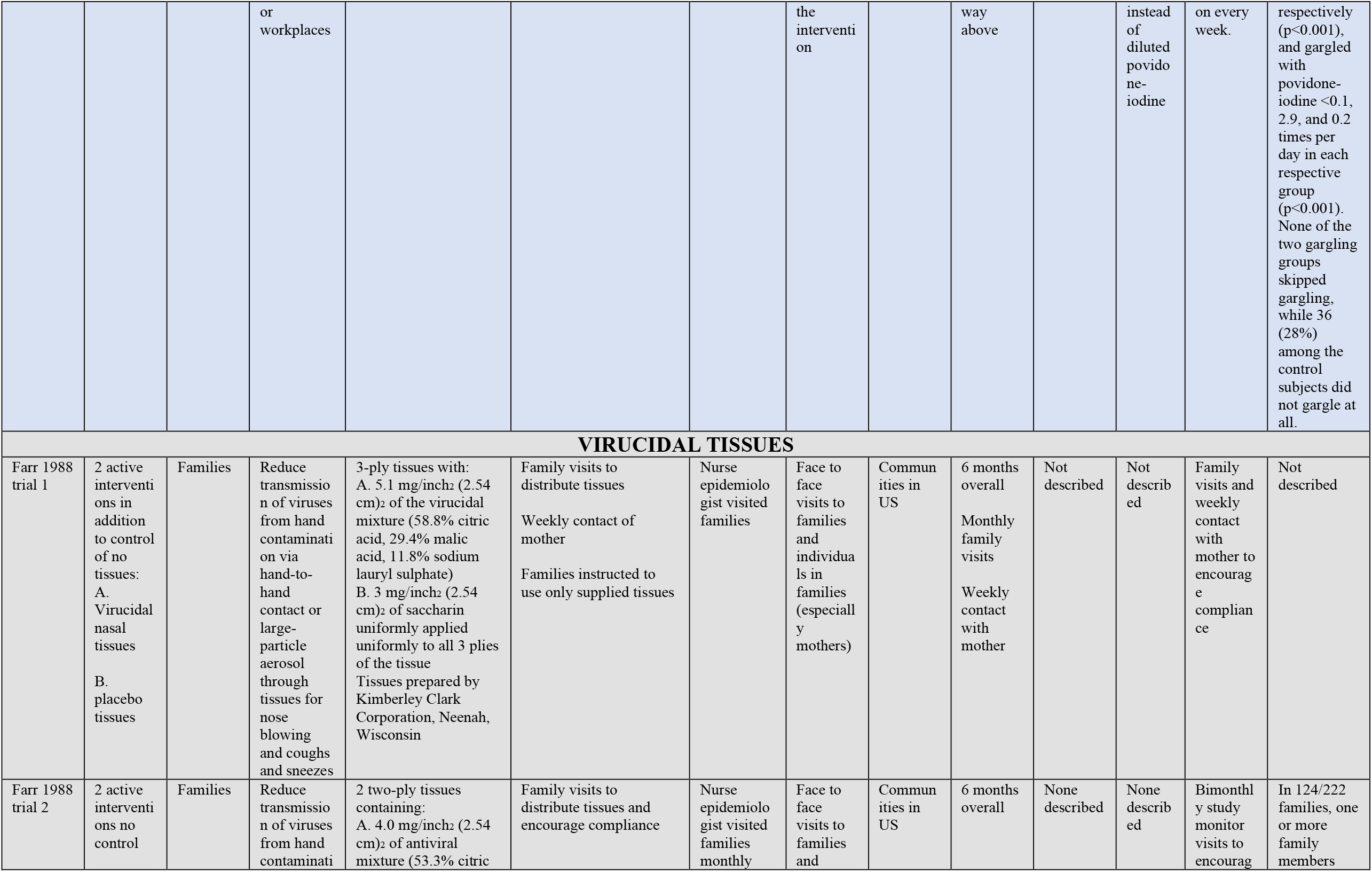
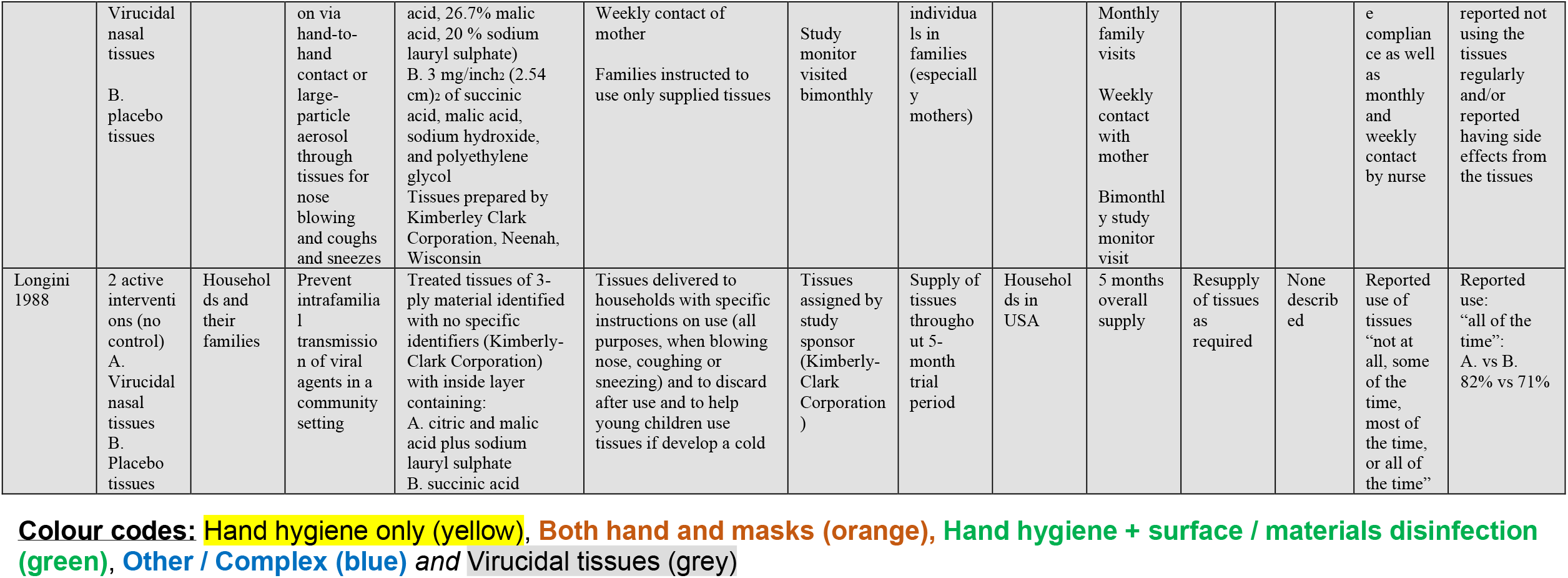
Description of interventions in included studies, using the items from the Template for Intervention Description and Replication (TIDieR) checklist.

Pooling of eight trials [14-19, 21] for the broad outcome of acute respiratory illness (ARI) showed a 16% relative reduction in the numbers of participants with ARI in the hand hygiene group (risk ratio (RR) 0.84, 95% CI 0.82 to 0.86) (Figure 2). However, when considering the more strictly-defined outcomes of influenza-like illness (ILI) and laboratory-confirmed influenza (influenza) [15, 16, 20, 22-28], the results were heterogeneous and not as strongly in favour of hand hygiene (RR 0.98, 95% CI 0.85 to 1.14 for ILI; RR 0.91, 95% CI 0.61 to 1.34 for influenza). Three trials quantified absenteeism from school or work [17, 23, 30] and demonstrated a 36% relative reduction in the absentee numbers in the hand hygiene group (RR 0.64, 95% CI 0.58 to 0.71). All 15 trials could be pooled for analysis of the composite outcome ‘ARI or ILI or influenza’, each study contributing the most comprehensive outcome reported only once. This showed a statistically significant 10% relative reduction of participants with a respiratory illness in favour of hand hygiene (95% CI 0.84 to 0.96) but with high heterogeneity (Figure 2).

We considered that studies in children might show a different effect from studies in adults or households, so we looked at the nine studies conducted in children [14, 17, 19-21, 25-28] (Figure S3). The result was consistent with the overall one, with RR for intervention to control being 0.92 (95%CI 0.84 to 1.01) with high heterogeneity.

A further 10 trials that compared hand hygiene to control [31-40] presented insufficient information to include in meta-analyses. The results of these trials were consistent with the findings from our meta-analysis. Characteristics and results from these studies are presented in Tables 1 and 2.

### Comparisons of different types of hand hygiene interventions

Five trials compared different hand hygiene interventions in a variety of settings, such as schools, low-income neighbourhoods and workplaces. There was considerable variation in interventions and insufficient information in trial reports to enable meta-analysis of these results (see Tables 1, 2 and 3 for details of the trials, interventions and outcomes). These trials looked at interventions such as soap and water, hand sanitiser, body wash and skin wipes, with or without additional hygiene education. One trial [41] compared different frequencies of handwashing in a kindergarten and found that compulsory handwashing every hour with an alcohol-based hand gel reduced absenteeism due to ILI more than handwashing every two hours or handwashing just before lunch. Morton et al [42] showed that using an alcohol gel as an adjunct to handwashing was more effective than handwashing alone in a primary school setting. However, this was not confirmed in another trial where both groups also received hygiene education [43]. Savolainen-Kopra et al [44] found no difference between soap and water and an alcohol-based hand rub. In low-income neighbourhoods in Pakistan, plain soap was equally effective as antibacterial soap in preventing ARIs in children aged under 5 years [45].

### Hand hygiene and masks versus control

Seven trials compared a combined intervention of hand hygiene and face masks with control (see Table 1, 2 and 3). These trials were carried out in households [15, 22, 26, 46] (4 trials), university student residences [47, 48] (2 trials), and a group of pilgrims at the annual Hajj [49] (1 trial). Pooling did not demonstrate a statistically significant difference between groups for the outcomes of ILI and influenza. However, the number of trials and events was lower than for the comparison of hand hygiene alone versus control, therefore CIs were wide. For ILI, the RR for intervention to control was 0.97 (95% CI 0.80 to 1.19) with high heterogeneity, and for influenza it was 0.97 (95% CI 0.69 to 1.36) (Figure 3)

### Hand hygiene and masks versus hand hygiene alone

Three trials studied the incremental benefit of masks in addition to hand hygiene on the outcomes of ILI and influenza in households [15, 22, 26]. When pooled there was no statistically significant difference between groups for either outcome (RR for intervention to control for ILI was 1.03, 95% CI 0.69 to 1.53; and for influenza the RR was 0.99, 95% CI 0.69 to 1.44) (Figure 4)

### Hand hygiene and disinfection of surfaces, objects, or environment versus control

Seven trials compared an intervention consisting of hand hygiene and disinfection of surfaces, toys, linen or other components of the environment with a control (see Table 1). Variation in scope and type of interventions and insufficient data in trial reports precluded meta-analysis. Three trials in young children [50-52] found a reduction of respiratory infections or absences, but the other three trials found no difference between intervention and control groups with children [53] [54], or in rates of respiratory infections in nursing home residents [55].

One study [56] compared disinfection of toys and linen in childcare centres (without a hand hygiene intervention) with a control. This trial found a statistically significant reduction in some respiratory viruses on surfaces, but not in coronaviruses.

### Virucidal tissues versus placebo

Three trials included in 2 papers [57] [58] investigated the role of virucidal tissues (see Table 1, and supplementary Table S1 with authors’ outcome definitions) in interrupting transmission of naturally occurring respiratory infections in households. The results were inconsistent and suggest that virucidal tissues do not play a major role in stopping people from passing on respiratory viruses within their household.

### Complex hygiene interventions versus control

Four c-RCTs implemented complex, multi-modal sanitation, education, cooking and hygiene interventions (see Tables 1, 2 and S1). All four trials were conducted in low-income countries in settings with minimal to no access to basic sanitation. In one trial in 100 primary schools in Laos [59] clean water supply, sanitation facilities, handwashing equipment and drinking water filters were provided to the intervention schools as part of a Water, Sanitation and Hygiene (WASH) programme with hygiene education. The control schools received the intervention after the trial period. They did not find an effect on infections or absenteeism amongst the students and conclude that such interventions alone are insufficient. The study by Huda et al [60] in villages in Bangladesh used local ‘promotors’ to visit households with young children regularly to deliver hygiene education including handwashing, latrine use, faeces and waste disposal, and water storage. Although the intervention households showed more ‘desired behaviour’ this did not result in a measurable reduction of respiratory illness. In another study in urban Bangladesh [61], handwashing equipment (bucket, tap, soap) and point-of-use water treatment were provided to intervention households and community health promotors delivered a behavioural intervention in addition to an oral cholera vaccine. There was no impact of this large-scale intervention on respiratory illness. However, the prevalence of respiratory illness was lower in those who had access to soap and water, irrespective of assignment to intervention or control group. A comprehensive intervention in rural Peru delivered stoves, kitchen sinks, plastic bottles for solar water treatment and hygiene education to households [62]. They did not find an effect on respiratory infections in children and suggest persistent poor indoor air quality may play a role. Another study [63] conducted among healthy adults compared throat gargling with water or povodine-iodine versus control. It was found that simple water gargling was effective in preventing and reducing the severity of upper respiratory tract infections.

### Safety of hand hygiene and other hygiene measures

Of all the included randomised trials, only six reported any outcomes related to the safety of these measures. [21, 39, 41, 43, 44, 46]. Skin and hand irritation were the most commonly reported adverse outcome in three out of four trials [21, 39, 43]. Two trials stated that no adverse outcomes were encountered among the participants. [41, 44]

## DISCUSSION

### Statement of MAIN findings

The composite outcome of ‘ARI or ILI or influenza”, the outcome of ARI alone and the outcome of absenteeism showed a significant benefit from hand hygiene. A non-significant benefit in favour of hand hygiene for laboratory-confirmed influenza may reflect less effect for this specific virus or sample size issues for a more rigourous outcome measure. We did not find an effect of combined hand hygiene and masks interventions, although there were few trials, mostly small in size, so CIs are wide. Similarly, we did not find a benefit for the addition of masks to hand hygiene, again CIs are wide, and it is difficult to draw a conclusion from these data. There were too few trials comparing different types of hand hygiene interventions to be certain of any differences between soap and water, alcohol-based hand sanitisers, or other types of interventions.

The findings in this review with respect to hand hygiene should be considered generally relevant to viral respiratory infections, given that diverse population categories of adults, children and families were studied and in multiple congregate settings including schools, childcare centres, homes and offices. These represent real world settings where transmission of viral respiratory infections occurs. Most respiratory viruses including SARS-CoV-2 are considered to be predominantly spread via respiratory droplets and/or contact routes [64]. Data from studies of SARS-CoV-2 contamination of the environment based on the presence of viral RNA, suggests significant fomite contamination from the virus. [65] [66] Hand hygiene would be expected to be beneficial of benefit in reducing the spread of this virus given the Joint Mission on COVID-19 of 75,465 cases suggested the majority of transmissions occurred within families in close contact with each other [64].

Studies on the major coronavirus pathogens including SARS-CoV-2, indicate that they are relatively persistent on surfaces such as wood, glass, metal, and especially plastic, with viability maintained from 4 hours to 9 days [*67*] *and* [*68*]. This family of viruses is very susceptible to the concentrations of alcohol commonly found in most hand sanitising preparations used for hand hygiene, suggesting biologic plausibility for their inactivation [67]. Similarly, the SARS-CoV-2 would be readily inactivated by most commonly used disinfectants.

A recent study from a designated COVID-19 hospital in Wuhan, China, suggested that contact was one of the major routes of transmission of SARS-CoV-2 [69]. They found that poor hand hygiene, despite the use of full PPE, was independently associated with a risk of SARS-CoV-2 HCW transmission using multivariate logistic regression in a retrospective cohort study with a relative risk (RR) of 3.07 (95% CI 1.14-5.15) and 2.45 (95% CI1.45-4.03), respectively, in a high risk and low risk clinical unit, which supports the overall findings of our review with specific reference to the COVID-19 virus.[69]

### Strengths and weaknesses of the study

This update of our review focused on the evidence from RCTs, providing a higher level of certainty, compared with the previous version, which also meta-analysed observational studies. However, many of the trials were small and hence underpowered, and at high or unclear risk of bias due to poor reporting of methods, and lack of blinding. The populations, outcomes, comparators and interventions tested were heterogeneous. Interventions were often complex in their implementation (e.g. handwashing education aimed at different stakeholders with specific instructions) and adherence was often modest or not reported. Studies were either situated in households or more closely monitored and regulated environments such as day care centres, primary schools, or army training centres. Only three of the trials in this review were conducted in a healthcare setting: all in nursing homes [55] [37] [40], and included healthcare workers, limiting generalisability of the results to those settings. Although two trials took place during the years of the previous SARS pandemic (2002 to 2003), the interventions were not tailored to this situation, and reducing transmission of SARS was not mentioned in the trials [19, 45]. Questions about effectiveness during an outbreak with the magnitude of the current SARS CoV-2 pandemic therefore remain unanswered.

### Relationship to other studies

There are several previous systematic reviews on hand hygiene and respiratory infections. Five reviewed the evidence in a community setting [70] [71] [72] [73] [74], and three focussed specifically on children [75] [76] [77]. The earliest review by Rabie et al in 2006 [70] included 8 studies and only 3 were randomised trials. Although each individual study was suggestive of an impact of handwashing on ARI, the pooled estimate of 7 studies was described as “indicative”, as studies were few, of poor quality, and limited in geographical scope. The review by Warren-Gash, 2013 [71] included 16 studies (10 were randomised trials), showed mixed results with inconclusive. Wong et al [72] identified 10 RCTs and reported that the combination of hand hygiene with facemasks in the developed countries only (5 trials) had statistically significant effect on reducing laboratory-confirmed influenza (RR = 0.73; 95% CI = 0.53 to 0.99; I2 = 0%; p = 0.05) and ILI outcomes (RR = 0.78; 95% CI = 0.68 to 0.90; I2 = 0%; p = 0.0008), while hand hygiene alone did not show significant reduction in respiratory outcomes. This significant reduction in ILI and influenza for hand hygiene and facemasks was possibly based on the raw numbers without considering any cluster effects in the included cluster trials which produced narrow confidence intervals and possibly biased treatment effect estimates. Moreover, trials from the less developed countries were not included in the review and this significant effect was not sustained when all the trials identified in the review were combined. Saudners-Hastings et al, [73] reviewed all the studies on the effectiveness of personal protective measures (PPM) in interrupting pandemic influenza transmission but identified only 2 randomised trials [46] [78] which reported a significant effect of hand hygiene. In a recent review by Moncion et al [74], 7 randomised trials of hand hygiene compared to control were identified but the majority did not find statistically significant differences in SARs for laboratory-confirmed or possible influenza between hand hygiene and control groups.

Systematic reviews of RCTs on hand hygiene interventions among children [75] [76] or at a non-clinical workplace [77], identified heterogeneous trials with quality issues including small numbers of clusters and participants, inadequate randomisation and self-reported outcomes. Evidence of impact on respiratory infections was equivocal.

### Unanswered questions and future research

Though the studies reviewed make it clear that hand hygiene has a modest effect, several questions remain unanswered. First, the high heterogeneity means there may be substantial differences in the effect of different interventions. The poor reporting limited our ability to extract the information needed to assess any “dose response” relationship. Second, the sustainability of hand washing is unclear: some programmes achieved 5 to 10 hand washings per day, but compliance may diminish with time, as motivation decreases or because of adverse effects from frequent hand washing. Third, there is still little information on combinations of hand washing with other interventions, and how those are best introduced and sustained. Finally, and perhaps most importantly, most interventions were intensive within small organisations, and the ability to scale these up to national-level interventions is unclear.

## CONCLUSIONS

The use of hand hygiene is an essential component of the WHO recommendations for epidemic and pandemic respiratory virus infections transmitted predominantly by the droplet and contact route, along with strict adherence with the use of personal protective equipment [79]. The benefits of hand hygiene found in our study have important implications for policymakers and support the recommendations for hand hygiene in the current WHO recommendations for COVID-19 [64].

The combined effect of the trials is small, but highly statistically significant. There is also considerable variation across trials, suggesting a different impact across implementation methods and settings. More research is urgently needed into the sources of variability of these effects. However, given the low cost and minimal disruption from good hand washing behaviour, we believe this small effect warrants continued promotion as part of a combined strategy to reduce the spread of respiratory viruses. No single strategy - other than prolonged isolation of the entire population - can block an epidemic spread. The alternative is combining multiple, partially effective interventions, such as hand washing, crowd reduction, or self-isolation of symptomatic patients, etc. This model is used in medical error reduction, where a common analogy is the “Swiss cheese model” whereby enough slices of cheese are needed to prevent the holes lining up, and an error occurring. Hand washing is clearly not a complete solution but adds one important layer of a combined strategy to reduce respiratory viral transmission.

## Data Availability

This is a systematic review with meta-analysis which used the data of the included studies. All the data used in the forest plots are available. Dr Jones (the manuscript’s guarantor) affirms that the manuscript is an honest, accurate, and transparent account of the study being reported; that no important aspects of the study have been omitted; and that any discrepancies from the study as planned (and, if relevant, registered) have been explained.

## Contributorship

All authors contributed equally to the design of the update, screening, extraction, interpretation and writing the manuscript which is approved by all authors. JC designed and carried out the searches and MJ and EB carried out the analysis.

## Acknowledgements

The authors thank Dr Elizabeth Gibson for her assistance with data extraction.

## APPENDIX

### Search strings for data bases run on 01/04/2020

#### PubMed

(“Influenza, Human”[Mesh] OR “Influenzavirus A”[Mesh] OR “Influenzavirus B”[Mesh] OR “Influenzavirus C”[Mesh] OR Influenza[tiab] OR “Respiratory Tract Diseases”[Mesh] OR “Bacterial Infections/transmission”[Mesh] OR Influenzas[tiab] OR “Influenza-like”[tiab] OR ILI[tiab] OR Flu[tiab] OR Flus[tiab] OR “Common Cold”[Mesh:NoExp] OR “common cold”[tiab] OR colds[tiab] OR coryza[tiab] OR coronavirus[Mesh] OR “sars virus”[Mesh] OR coronavirus[tiab] OR Coronaviruses[tiab] OR “coronavirus infections”[Mesh] OR “severe acute respiratory syndrome”[Mesh] OR “severe acute respiratory syndrome”[tiab] OR “severe acute respiratory syndromes”[tiab] OR sars[tiab] OR “respiratory syncytial viruses”[Mesh] OR “respiratory syncytial virus, human”[Mesh] OR “Respiratory Syncytial Virus Infections”[Mesh] OR “respiratory syncytial virus”[tiab] OR “respiratory syncytial viruses”[tiab] OR rsv[tiab] OR parainfluenza[tiab] OR ((Transmission[tiab]) AND (Coughing[tiab] OR Sneezing[tiab])) OR ((respiratory[tiab] AND Tract[tiab]) AND (infection[tiab] OR Infections[tiab] OR illness[tiab])))

AND

(“Hand Hygiene”[Mesh] OR handwashing[tiab] OR hand-washing[tiab] OR ((Hand[tiab] OR Alcohol[tiab]) AND (wash[tiab] OR Washing[tiab] OR Cleansing[tiab] OR Rinses[tiab] OR hygiene[tiab] OR rub[tiab] OR Rubbing[tiab] OR sanitiser[tiab] OR sanitizer[tiab] OR cleanser[tiab] OR disinfected[tiab] OR Disinfectant[tiab] OR Disinfect[tiab] OR antiseptic[tiab] OR virucid[tiab])) OR “gloves, protective”[Mesh] OR Glove[tiab] OR Gloves[tiab] OR Masks[Mesh] OR “respiratory protective devices”[Mesh] OR facemask[tiab] OR Facemasks[tiab] OR mask[tiab] OR Masks[tiab] OR respirator[tiab] OR respirators[tiab] OR “Protective Clothing”[Mesh:NoExp] OR “Protective Devices”[Mesh] OR “patient isolation”[tiab] OR ((school[tiab] OR Schools[tiab]) AND (Closure[tiab] OR Closures[tiab] OR Closed[tiab])) OR Quarantine[Mesh] OR quarantine[tiab] OR “Hygiene intervention”[tiab] OR “Mouthwashes”[Mesh] OR gargling[tiab] OR “nasal tissues”[tiab] OR “Eye Protective Devices”[Mesh] OR Glasses[tiab] OR Goggle[tiab] OR “Eye protection”[tiab] OR Faceshield[tiab] OR Faceshields[tiab] OR Goggles[tiab] OR “Face shield”[tiab] OR “Face shields”[tiab] OR Visors[tiab])

AND

(“Communicable Disease Control”[Mesh] OR “Disease Outbreaks”[Mesh] OR “Disease Transmission, Infectious”[Mesh] OR “Infection Control”[Mesh] OR Transmission[sh] OR “Prevention and control”[sh] OR “Communicable Disease Control”[tiab] OR “Secondary transmission”[tiab] OR ((Reduced[tiab] OR Reduce[tiab] OR Reduction[tiab] OR Reducing[tiab] OR Lower[tiab]) AND (Incidence[tiab] OR Occurrence[tiab] OR Transmission[tiab] OR Secondary[tiab])))

AND

(Randomized controlled trial[pt] OR controlled clinical trial[pt] OR randomized[tiab] OR randomised[tiab] OR placebo[tiab] OR “drug therapy”[sh] OR randomly[tiab] OR trial[tiab] OR groups[tiab])

NOT

(Animals[Mesh] not (Animals[Mesh] and Humans[Mesh]))

NOT

(“Case Reports”[pt] OR Editorial[pt] OR Letter[pt] OR Meta-Analysis[pt] OR “Observational Study”[pt] OR “Systematic Review”[pt] OR “Case Report”[ti] OR “Case series”[ti] OR Meta-Analysis[ti] OR “Meta Analysis”[ti] OR “Systematic Review”[ti])

### CENTRAL

([mh “Influenza, Human”] OR [mh “Influenzavirus A”] OR [mh “Influenzavirus B”] OR [mh “Influenzavirus C”] OR Influenza:ti,ab OR [mh “Respiratory Tract Diseases”] OR Influenzas:ti,ab OR “Influenza-like”:ti,ab OR ILI:ti,ab OR Flu:ti,ab OR Flus:ti,ab OR [mh ^”Common Cold”] OR “common cold”:ti,ab OR colds:ti,ab OR coryza:ti,ab OR [mh coronavirus] OR [mh “sars virus”] OR coronavirus:ti,ab OR Coronaviruses:ti,ab OR [mh “coronavirus infections”] OR [mh “severe acute respiratory syndrome”] OR “severe acute respiratory syndrome”:ti,ab OR “severe acute respiratory syndromes”:ti,ab OR sars:ti,ab OR [mh “respiratory syncytial viruses”] OR [mh “respiratory syncytial virus, human”] OR [mh “Respiratory Syncytial Virus Infections”] OR “respiratory syncytial virus”:ti,ab OR “respiratory syncytial viruses”:ti,ab OR rsv:ti,ab OR parainfluenza:ti,ab OR ((Transmission) AND (Coughing OR Sneezing)) OR ((respiratory:ti,ab AND Tract) AND (infection:ti,ab OR Infections:ti,ab OR illness:ti,ab)))

AND

([mh “Hand Hygiene”] OR handwashing:ti,ab OR “hand-washing”:ti,ab OR ((Hand:ti,ab OR Alcohol:ti,ab) AND (wash:ti,ab OR Washing:ti,ab OR Cleansing:ti,ab OR Rinses:ti,ab OR hygiene:ti,ab OR rub:ti,ab OR Rubbing:ti,ab OR sanitiser:ti,ab OR sanitizer:ti,ab OR cleanser:ti,ab OR disinfected:ti,ab OR Disinfectant:ti,ab OR Disinfect:ti,ab OR antiseptic:ti,ab OR virucid:ti,ab)) OR [mh “gloves, protective”] OR Glove:ti,ab OR Gloves:ti,ab OR [mh Masks] OR [mh “respiratory protective devices”] OR facemask:ti,ab OR Facemasks:ti,ab OR mask:ti,ab OR Masks:ti,ab OR respirator:ti,ab OR respirators:ti,ab OR [mh ^”Protective Clothing”] OR [mh “Protective Devices”] OR “patient isolation”:ti,ab OR ((school:ti,ab OR Schools:ti,ab) AND (Closure:ti,ab OR Closures:ti,ab OR Closed:ti,ab)) OR [mh Quarantine] OR quarantine:ti,ab OR “Hygiene intervention”:ti,ab OR [mh Mouthwashes] OR gargling:ti,ab OR “nasal tissues”:ti,ab OR [mh “Eye Protective Devices”] OR Glasses:ti,ab OR Goggle:ti,ab OR “Eye protection”:ti,ab OR Faceshield:ti,ab OR Faceshields:ti,ab OR Goggles:ti,ab OR “Face shield”:ti,ab OR “Face shields”:ti,ab OR Visors:ti,ab)

AND

([mh “Communicable Disease Control”] OR [mh “Disease Outbreaks”] OR [mh “Disease Transmission, Infectious”] OR [mh “Infection Control”] OR “Communicable Disease Control”:ti,ab OR “Secondary transmission”:ti,ab OR ((Reduced:ti,ab OR Reduce:ti,ab OR Reduction:ti,ab OR Reducing:ti,ab OR Lower:ti,ab) AND (Incidence:ti,ab OR Occurrence:ti,ab OR Transmission:ti,ab OR Secondary:ti,ab)))

### Embase

(‘influenza’/exp OR Influenza:ti,ab OR ‘Respiratory Tract Disease’/exp OR Influenzas:ti,ab OR Influenza-like:ti,ab OR ILI:ti,ab OR Flu:ti,ab OR Flus:ti,ab OR ‘Common Cold’/de OR “common cold”:ti,ab OR colds:ti,ab OR coryza:ti,ab OR ‘coronavirus’/exp OR ‘SARS coronavirus’/exp OR coronavirus:ti,ab OR Coronaviruses:ti,ab OR ‘coronavirus infection’/exp OR ‘severe acute respiratory syndrome’/exp OR “severe acute respiratory syndrome”:ti,ab OR “severe acute respiratory syndromes”:ti,ab OR sars:ti,ab OR ‘Pneumovirus’/exp OR ‘Human respiratory syncytial virus’/exp OR “respiratory syncytial virus”:ti,ab OR “respiratory syncytial viruses”:ti,ab OR rsv:ti,ab OR parainfluenza:ti,ab OR ((Transmission) AND (Coughing OR Sneezing)) OR ((respiratory:ti,ab AND Tract) AND (infection:ti,ab OR Infections:ti,ab OR illness:ti,ab)))

AND

(‘hand washing’/exp OR handwashing:ti,ab OR hand-washing:ti,ab OR ((Hand:ti,ab OR Alcohol:ti,ab) AND (wash:ti,ab OR Washing:ti,ab OR Cleansing:ti,ab OR Rinses:ti,ab OR hygiene:ti,ab OR rub:ti,ab OR Rubbing:ti,ab OR sanitiser:ti,ab OR sanitizer:ti,ab OR cleanser:ti,ab OR disinfected:ti,ab OR Disinfectant:ti,ab OR Disinfect:ti,ab OR antiseptic:ti,ab OR virucid:ti,ab)) OR ‘protective glove’/exp OR Glove:ti,ab OR Gloves:ti,ab OR ‘mask’/exp OR ‘gas mask’/exp OR facemask:ti,ab OR Facemasks:ti,ab OR mask:ti,ab OR Masks:ti,ab OR respirator:ti,ab OR respirators:ti,ab OR ‘protective clothing’/de OR ‘protective equipment’/exp OR “patient isolation”:ti,ab OR ((school:ti,ab OR Schools:ti,ab) AND (Closure:ti,ab OR Closures:ti,ab OR Closed:ti,ab)) OR ‘Quarantine’/exp OR quarantine:ti,ab OR “Hygiene intervention”:ti,ab OR ‘mouthwash’/exp OR gargling:ti,ab OR “nasal tissues”:ti,ab OR ‘eye protective device’/exp OR Glasses:ti,ab OR Goggle:ti,ab OR “Eye protection”:ti,ab OR Faceshield:ti,ab OR Faceshields:ti,ab OR Goggles:ti,ab OR “Face shield”:ti,ab OR “Face shields”:ti,ab OR Visors:ti,ab)

AND

(‘Communicable Disease Control’/exp OR ‘epidemic’/exp OR ‘disease transmission’/exp OR ‘Infection Control’/exp OR “Communicable Disease Control”:ti,ab OR “Secondary transmission”:ti,ab OR ((Reduced:ti,ab OR Reduce:ti,ab OR Reduction:ti,ab OR Reducing:ti,ab OR Lower:ti,ab) AND (Incidence:ti,ab OR Occurrence:ti,ab OR Transmission:ti,ab OR Secondary:ti,ab)))

AND

(random* OR factorial OR crossover OR placebo OR blind OR blinded OR assign OR assigned OR allocate OR allocated OR ‘crossover procedure’/exp OR ‘double-blind procedure’/exp OR ‘randomized controlled trial’/exp OR ‘single-blind procedure’/exp NOT (‘animal’/exp NOT (‘animal’/exp AND ‘human’/exp)))

### CINAHL

((MH “Influenza, Human+”) OR (MH “Orthomyxoviridae+”) OR TI Influenza OR AB Influenza OR (MH “Respiratory Tract Diseases+”) OR TI Influenzas OR AB Influenzas OR TI Influenza-like OR AB Influenza-like OR TI ILI OR AB ILI OR TI Flu OR AB Flu OR TI Flus OR AB Flus OR (MH “Common Cold+”) OR TI “common cold” OR AB “common cold” OR TI colds OR AB colds OR TI coryza OR AB coryza OR (MH “coronavirus+”) OR (MH “sars virus+”) OR TI coronavirus OR AB coronavirus OR TI Coronaviruses OR AB Coronaviruses OR (MH “coronavirus infections+”) OR (MH “severe acute respiratory syndrome+”) OR TI “severe acute respiratory syndrome” OR AB “severe acute respiratory syndrome” OR TI “severe acute respiratory syndromes” OR AB “severe acute respiratory syndromes” OR TI sars OR AB sars OR (MH “respiratory syncytial viruses+”) OR TI “respiratory syncytial virus” OR AB “respiratory syncytial virus” OR TI “respiratory syncytial viruses” OR AB “respiratory syncytial viruses” OR TI rsv OR AB rsv OR TI parainfluenza OR AB parainfluenza OR ((Transmission) AND (Coughing OR Sneezing)) OR ((TI respiratory OR AB respiratory AND Tract) AND (TI infection OR AB infection OR TI Infections OR AB Infections OR TI illness OR AB illness)))

AND

((MH “Handwashing+”) OR TI handwashing OR AB handwashing OR TI hand-washing OR AB hand-washing OR ((TI Hand OR AB Hand OR TI Alcohol OR AB Alcohol) AND (TI wash OR AB wash OR TI Washing OR AB Washing OR TI Cleansing OR AB Cleansing OR TI Rinses OR AB Rinses OR TI hygiene OR AB hygiene OR TI rub OR AB rub OR TI Rubbing OR AB Rubbing OR TI sanitiser OR AB sanitiser OR TI sanitizer OR AB sanitizer OR TI cleanser OR AB cleanser OR TI disinfected OR AB disinfected OR TI Disinfectant OR AB Disinfectant OR TI Disinfect OR AB Disinfect OR TI antiseptic OR AB antiseptic OR TI virucid OR AB virucid)) OR (MH “gloves+”) OR TI Glove OR AB Glove OR Gloves OR (MH “Masks+”) OR (MH “respiratory protective devices+”) OR TI facemask OR AB facemask OR TI Facemasks OR AB Facemasks OR TI mask OR AB mask OR TI Masks OR AB Masks OR TI respirator OR AB respirator OR TI respirators OR AB respirators OR (MH “Protective Clothing”) OR (MH “Protective Devices+”) OR TI “patient isolation” OR AB “patient isolation” OR ((TI school OR AB school OR TI Schools OR AB Schools) AND (TI Closure OR AB Closure OR TI Closures OR AB Closures OR TI Closed OR AB Closed)) OR (MH “Quarantine+”) OR TI quarantine OR AB quarantine OR TI “Hygiene intervention” OR AB “Hygiene intervention” OR (MH “Mouthwashes+”) OR TI gargling OR AB gargling OR TI “nasal tissues” OR AB “nasal tissues” OR (MH “Eye Protective Devices+”) OR TI Glasses OR AB Glasses OR TI Goggle OR AB Goggle OR TI “Eye protection” OR AB “Eye protection” OR TI Faceshield OR AB Faceshield OR TI Faceshields OR AB Faceshields OR TI Goggles OR AB Goggles OR TI “Face shield” OR AB “Face shield” OR TI “Face shields” OR AB “Face shields” OR TI Visors OR AB Visors)

AND

((MH “Infection Control+”) OR (MH “Disease Outbreaks+”) OR (MH “Infection Control+”) OR TI “Communicable Disease Control” OR AB “Communicable Disease Control” OR TI “Secondary transmission” OR AB “Secondary transmission” OR ((TI Reduced OR AB Reduced OR TI Reduce OR AB Reduce OR TI Reduction OR AB Reduction OR TI Reducing OR AB Reducing OR TI Lower OR AB Lower) AND (TI Incidence OR AB Incidence OR TI Occurrence OR AB Occurrence OR TI Transmission OR AB Transmission OR TI Secondary OR AB Secondary)))

AND

((MH “Clinical Trials+”) OR (MH “Quantitative Studies”) OR TI placebo* OR AB placebo* OR (MH “Placebos”) OR (MH “Random Assignment”) OR TI random* OR AB random* OR TI ((singl* or doubl* or tripl* or trebl*) W1 (blind* or mask*)) OR AB ((singl* or doubl* or tripl* or trebl*) W1 (blind* or mask*)) OR TI clinic* trial* OR AB clinic* trial* OR PT clinical trial)

